# Genome-wide meta-analysis conducted in three large biobanks expands the genetic landscape of lumbar disc herniations

**DOI:** 10.1101/2023.10.15.23296916

**Authors:** Ville Salo, Juhani Määttä, Eeva Sliz, FinnGen, Ene Reimann, Reedik Mägi, Estonian Biobank Research Team, Kadri Reis, Abdelrahman G. Elhanas, Anu Reigo, Priit Palta, Tõnu Esko, Jaro Karppinen, Johannes Kettunen

## Abstract

**Introductory paragraph:** Given that lumbar disc herniation (LDH) is a prevalent spinal condition that causes significant individual suffering and societal costs^1^, the genetic basis of LDH has received relatively little research. Our aim was to increase understanding of the genetic factors influencing LDH. We performed a genome-wide association analysis (GWAS) of LDH in the FinnGen project and in Estonian and UK biobanks, followed by a genome-wide meta-analysis to combine the results. In the meta-analysis, we identified 41 loci that have not been associated with LDH in prior studies on top of the 23 known risk loci. We detected LDH-associated loci in the vicinity of genes related to inflammation, disc-related structures, and synaptic transmission. Overall, our research contributes to a deeper understanding of the genetic factors behind LDH, potentially paving the way for the development of new therapeutics, prevention methods, and treatments for symptomatic LDH in the future.

## Main text

Lumbar disc herniation (LDH) is one of the most frequent structural findings in the lumbar spine to cause specific symptoms. Disc herniation is a general term which includes different types of disc displacements such as disc protrusion, extrusion, and sequestration^2^. Disc protrusions, for instance, are rarely symptomatic, and they are prevalent even among asymptomatic subjects. The overall prevalence of LDH in the whole population has been estimated to be 14% in a large cross-sectional study, and herniations are found most frequently at the L4/5 and L5/S1 segments of the spine^3^.

LDH has a clinical relevance if it causes radicular symptoms in the lower extremity. LDH is, indeed, the most frequent condition to cause lumbar radicular pain, i.e., sciatica, or radiculopathy^4^. The mechanisms deriving LDH to cause radicular pain are manifold. Radicular pain is partly evoked by mechanical compression of the nerve root, but inflammatory mediators and autoimmune responses play a considerable role there too^5^. When the nerve root is perturbed, typical symptoms include radicular pain, paresthesia, or numbness in the area of the nerve, and possible weakness in the muscles innervated by the affected nerve root^6^.

Typically, patients with symptomatic LDH are treated conservatively, but surgery is required if there is a sudden or progressive neurological deficit or unmanageable pain despite appropriate conservative treatment^6^. Overall, surgery has not been proven to have superior outcomes in the long term, even though surgery can have better pain relief in the short term^7,8^. Most of the patients will recover from symptoms rather quickly, however some studies show conflicting evidence. Psychosocial factors, such as the patient’s own beliefs of recovery, can also have a role in prognosis^9^.

Genetic influence on LDH and sciatica has been established through studies that have shown certain loci to be associated with these conditions using genome-wide association analysis (GWAS)^10,11^. Symptomatic LDH could be caused by factors affecting either disc-related structures, such as collagen^12^, or other morphologies, such as nerve-related, inflammatory, or autoimmune structures^5^. Previous studies have associated several pathways with LDH pathogenesis, encompassing inflammation, chondroitin sulfation, collagen synthesis, and chondrogenic differentiation^10,11^. Some of these genes have also been associated with back pain and the regulation of pain sensations^13–15^. The etiological factors behind LDH are quite well understood^16^. However, the genetic factors behind these distinct features warrant more research. The aim of this study was to explore different genetic features behind LDH by conducting a GWAS using data from three large biobanks: FinnGen, Estonian Biobank, and UK Biobank.

In the meta-analysis, we identified 41 novel (Fig. S1.1-41, Table 1, Table S1) and replicated 23 known loci (Fig. S2, Table S2), each containing at least one genome-wide significantly associated variant associated with LDH as defined by International Classification of Diseases (ICD)-10 codes M51 (M51.1-51.9, Fig, 1, Table S3). In addition, secondary signals at 5 of the loci were observed in the conditional analysis (Table S2).We estimated LD score regression-derived SNP-based heritability to be 0.08 (standard error [SE] = 0.003), suggesting that genetic factors account for 8% of the common variation in LDH risk. The genomic inflation factor lambda (1.47) suggested inflation in the test statistics. Given that the intercept value was 1.12, inflation might be caused by a polygenic signal. In FinnGen data, SNP based heritability was estimated to be 14.3% [SE]=0.0083, and lambda 1.39 with intercept of 1.13. Currently, there are no published SNP-based heritability estimates for LDH and, therefore it would be important to replicate these results.

**Table 1.**
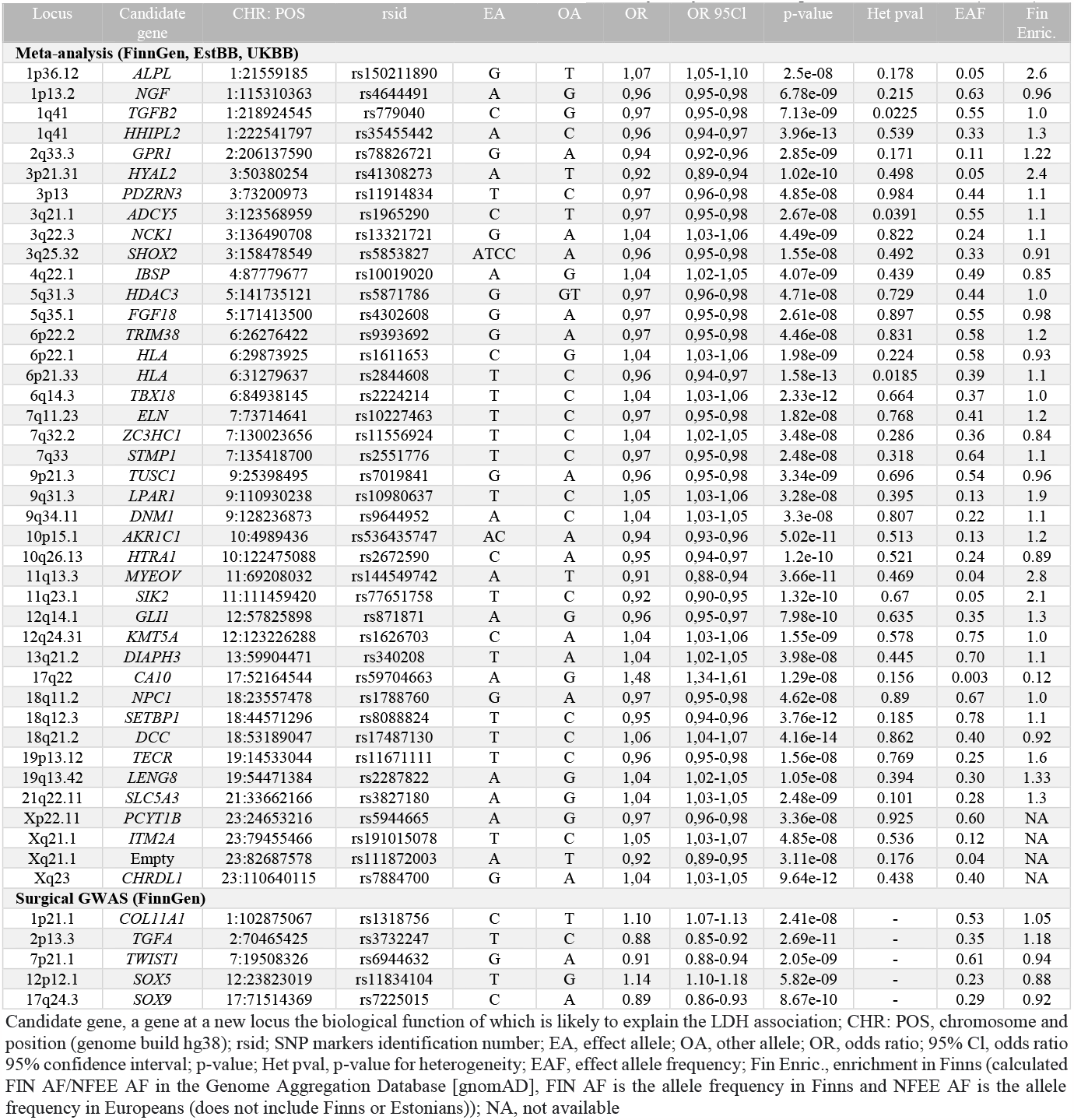
The list of novel lead variants at the 41 genome-wide significant (p<5x10^-8^) loci that were associated with LDH in the meta-analysis (top) and five novel loci associated with LDH related surgical operations in sensitivity analysis that were performed in FinnGen (bottom).

In two sets of sensitivity analyses conducted in FinnGen, more strict case definitions were used by limiting LDH cases to LDH cases with radiculopathy (M51.1) and to those who have undergone surgery (Table S1). No statistically significant differences in the effect sizes of the lead variants were observed between the original meta-analysis and the GWAS specific to the M51.1 endpoint. On the other hand, when comparing the original meta-analysis with the GWAS on LDH patients who underwent surgical treatment, differences in the effect sizes were observed for 9 variants (Fig. 1, Fig. S3, Table S4). In this analysis, we also identified five novel loci associated with LDH-related surgical operations were also found (Fig. S4, Table 1, Table S5).

**Fig. 1.**
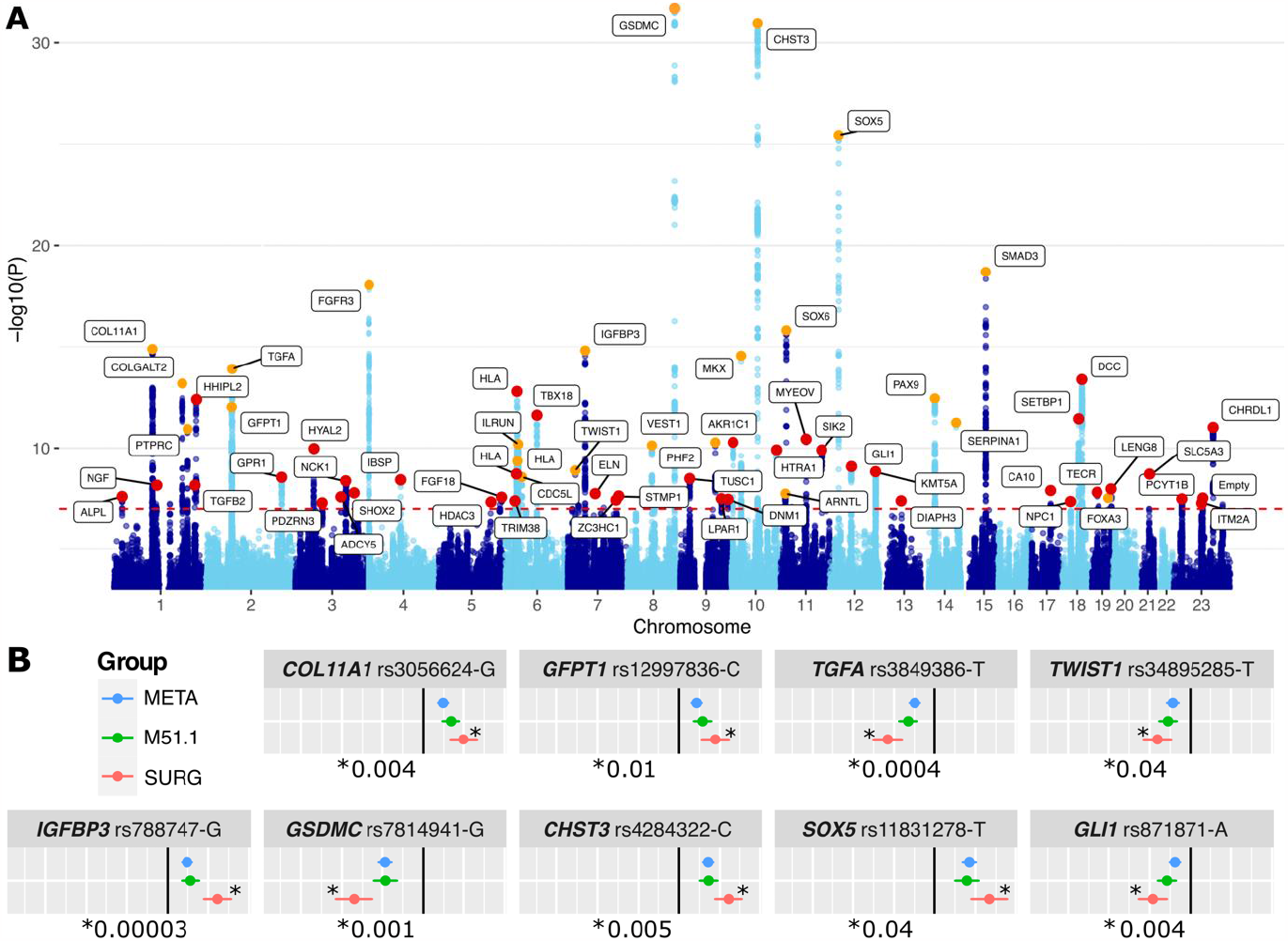
Above, the Manhattan plot of the associations detected to be associated with LDH in the meta-analysis of 80 724 cases and 748 975 controls. Previously reported loci are indicated with orange color, while 41 novel loci that we observed are highlighted in red. Candidate genes possibly explaining the LDH associations were used as loci identifiers. The red dashed line depicts the genome-wide significance limit (p<5x10^-8^). Below, a comparison of the effect sizes of the lead variants discovered in the original meta-analysis (ICD-10:M51 [blue]) and in sensitivity analyses with more strict case definitions (ICD-10:M51.1 [green] or a surgery [red]). Dots indicate effect size and vertical lines are the corresponding 95% confidence intervals. For effect differences statistical comparison, we used a two-tailed test, using group-specific effect estimates of the variants and the corresponding standard errors *((Effect_Meta-Effect_M51*.*1)/ sqrt(standarderror_Meta*^*2*^*+standarderror_M51*.*1*^*2*^)). A P-value <0.05 was considered the limit of a significant effect difference. * observed significant effect size differences and p-values of differences between the meta-analysis and surgery patients. No statistically significant effect differences were found between the meta-analysis and M51.1. Other variants can be seen in Fig. S3.

As reported previously, numerous LDH-associated loci were in the vicinity of genes related to inflammation or disc-related structures (Table S2), indicating that these pathways play a central role in LDH pathogenesis. In addition, we detected novel loci near genes related to the Wnt/β-catenin pathway, such as *PDZRN3* (*PDZ domain containing ring finger 1*, locus 3p13), activation of the Wnt/β-catenin pathway has been found to be associated with endplate degeneration, increased intervertebral disc (IVD) cell senescence, and extracellular matrix degradation^17,18^. LDH associations were also observed in loci near *NGF* (*nerve growth factor*), *DCC* (*DCC netrin-1 receptor*), and *NCK1* (*NCK adaptor protein 1*). These genes are involved in the growth and regulation of nerve axons^19,20^, suggesting a potential connection between genes affecting nerves and the nervous system and LDH pathogenesis. The dysfunction of these genes could lead to IVD innervation^19^, and increase sensation of pain. Notably, some of these genes have already been associated with pain sensation in previous studies^13,19^.

In addition, we observed novel LDH associations near *CA10* (*carbonic anhydrase 10*) and *DNM1* (*Dynamin 1*) that are involved in synaptic transmission. *CA10* blocks the binding of heparan sulfate to neurexin, which possibly affects the function of neurexins^21^. Neurexins are pre-synaptic cell adhesion molecules that play a role in connecting neurons at synapses. Heparan sulfate has been found to potentially expand the interactome of neurexins, and they also play a role in fine-tuning synaptic transmission^22^. *CA10* is expressed especially in the central nervous system, and it has been associated with chronic pain in previous studies^13,23^. *DNM1* plays a central role in the transmitting nociceptive messages within the nociceptive circuits in the dorsal horn of the spinal cord, where *DNM1*-mediated endocytosis of synaptic vesicles enables sustained neurotransmission^24^. While further studies are needed, the association of genes involved in synaptic transmission with LDH suggests that differences in synaptic transmission may influence differences in pain perception in patients with morphologically similar LDHs. Moreover, these genes could also contribute to the chronification of pain in patients with symptomatic LDH. The findings above underline the relevance of the physiological factors of the nervous system in addition to the disc-related structures behind symptomatic LDH.

The MAGMA gene-based test highlighted multiple genes that we had deemed as potential candidate genes at the novel LDH-associated loci (Table 1, Fig. 2A), thus providing supportive evidence for our findings. In the MAGMA gene-set analysis (Fig. 2B), we observed the most significant enrichments for pathways responsible for chondrocyte differentiation. Significant gene-set enrichments were also observed for cartilage and connective tissue development pathways, as well as for pathways related to chromatin organization and modification. Among the gene-sets related to the nervous system, we also observed significant enrichments in gene-sets related to the structure and active zone organization of presynapses. In the MAGMA tissue expression analysis (Fig. S5), we found no tissues showing a positive correlation between tissue-specific gene expression profiles and LDH associations. The likely reason for the null result is the absence of the relevant tissues, namely cartilage and bone, in the Genotype-Tissue Expression (GTEx) dataset used in these analyses.

**Fig. 2.**
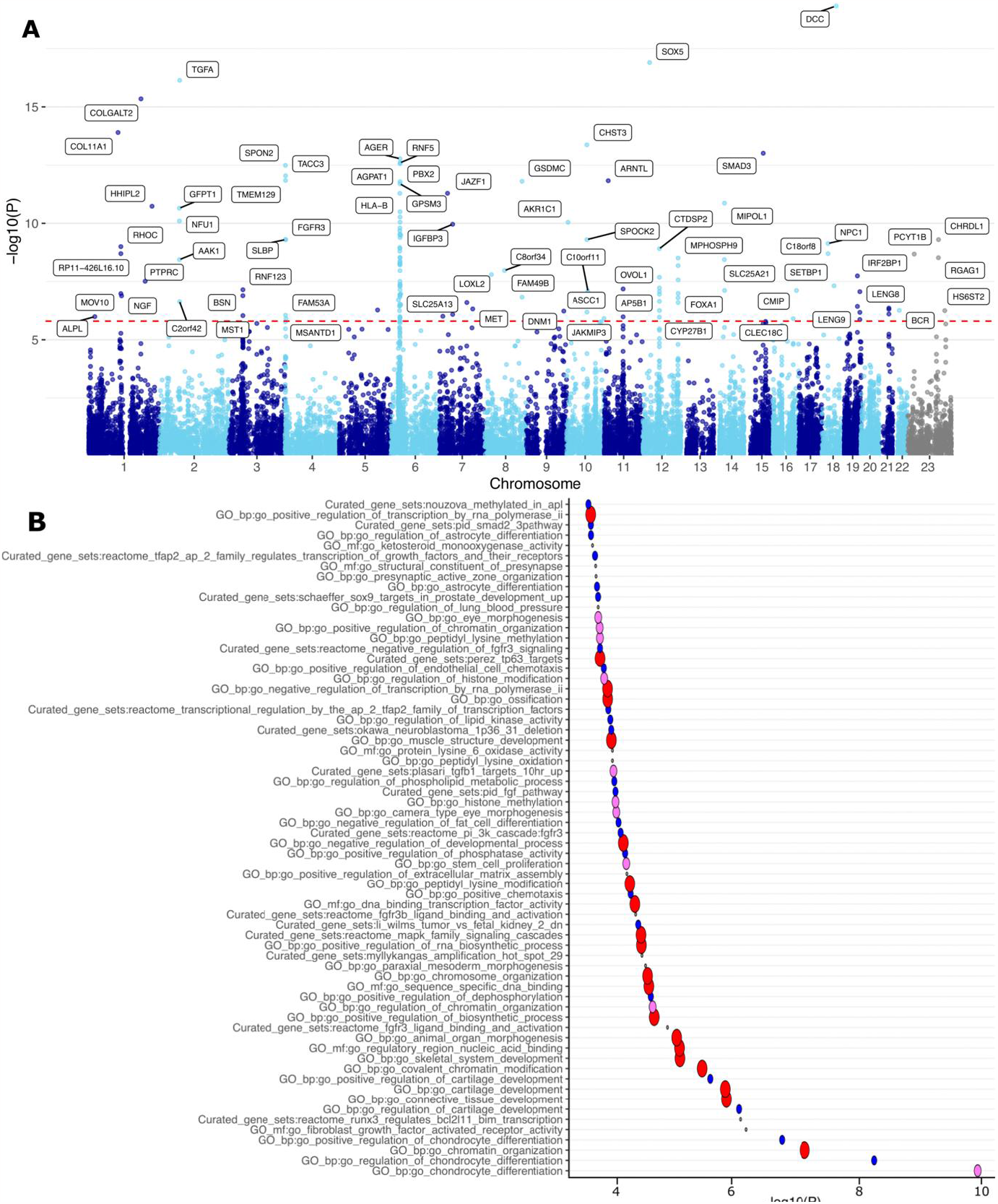
Above, results of the MAGMA^25^ gene-based test in a Manhattan plot. X-axis chromosomes, y-axis -log10(p-value). Below, MAGMA gene-set enrichment analysis. Plot shows significantly enriched pathways (pFDR < 0.05), curated gene sets, and GO-annotations ranked by p-value -log10(P). The size of the circles refers to the size of the gene set. Small gray <15, blue 15–100, violet 100-200, and red >200 genes. The analysis was done using FUMA^26^, and the gene sets and GO annotations included in the analysis are from MSigDB^27^.

For LDH-associated loci, we investigated the effect of variants on the accumulation of LDH diagnoses and, in addition, their effect on having to undergo surgery. As observed previously^28,29^, LDH diagnoses accumulated greatly between the ages of 40 and 50 until circa 70 years old, after which the accumulation of diagnoses was very low (Fig. 3). The differences between the variants were very small on average, with a few exceptions. For some variants, the LDH risk-increasing effects of different alleles were more noticeable. For two variants, a statistically significant difference between the genotypes was observed even before the age of 30. *GSDMC* (*gasdermin C*, Fig. 3A) differed from the variant’s other genotypes at the age of 26 (p=0.0005). *CHST3* (*carbohydrate sulfotransferase* 3, Fig. 3B) homozygotes became significantly different at the age of 25 (p=2.22e^-5^). For other variants, the differences became statistically significant at a later age, *IGFBP3* (*insulin like growth factor binding protein 3*, Fig 3C) at the 35 years of age (p=0.001) and *SOX6* (*SRY-box transcription factor 6*, Fig 3D) at the 36 years of age (p=0.03).These genes have already been associated with LDH, but this study is the first to observe the age at which diagnoses begin to accumulate for the variants in question. Otherwise, we found that the effect of the most loci was modest and followed the sample prevalence values (12.2% for LDH diagnoses and 2.6% for surgical patients, Table S6). We also repeated the analysis with only M51.1 cases, where all variants behaved identically in the analyses (Fig. S7). These accumulation results are only based on the analyses of the Finnish population (FinnGen), and these results would benefit from replication in other populations as well.

**Fig. 3.**
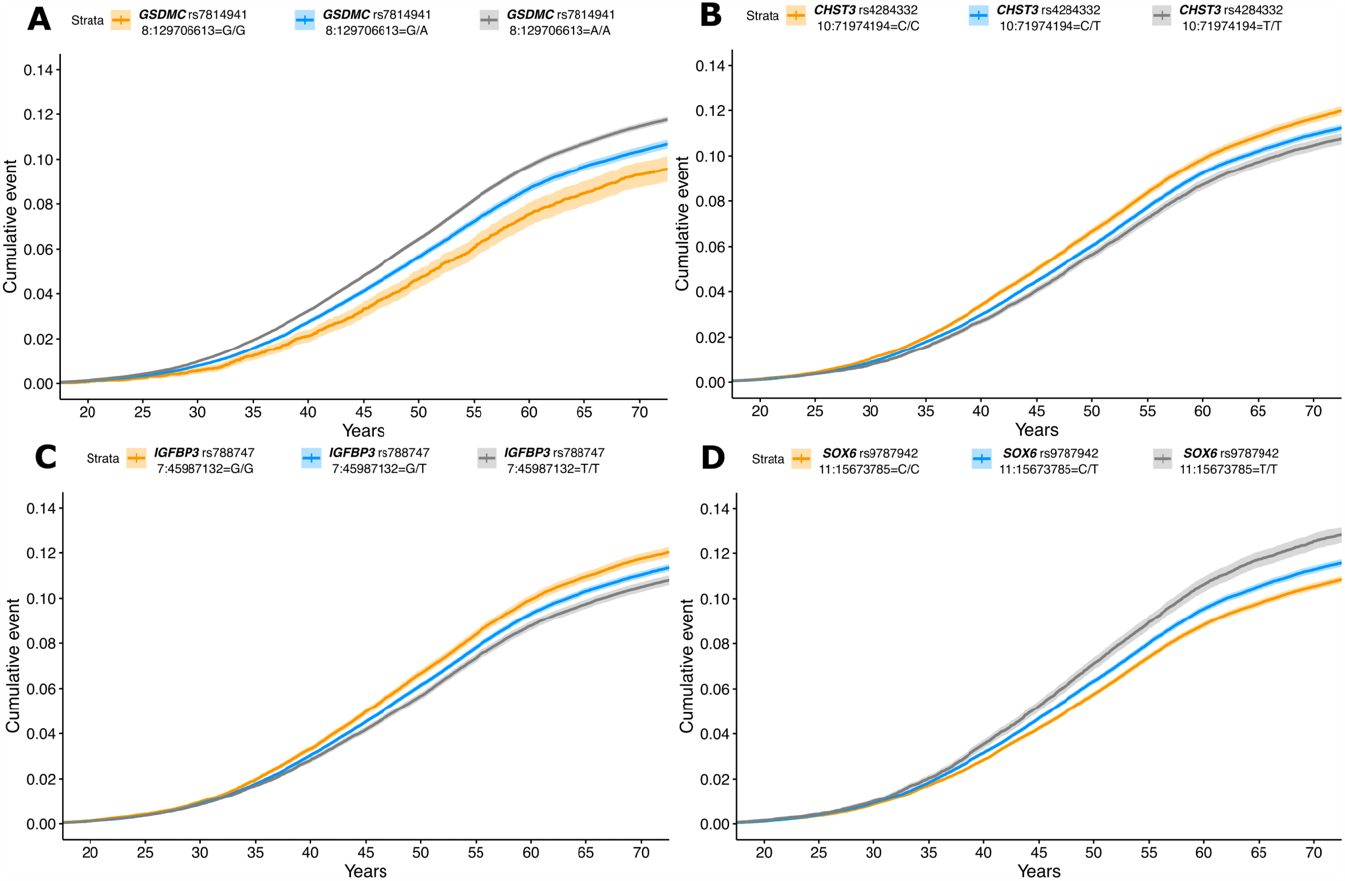
Kaplan-Meier plots for *GSDMC (rs7814941, 8:129706613:G:A),CHST3 (rs4284332, 10:719741194:C:T), IGFBP3 (rs788747, 7:45987132:G:T), SOX6 (rs9787942, 11:15673785:C:T)*. Age in years is on the x-axis of the graphs, and cumulative disease severity on the y-axis. The orange line depicts homozygotes for the effect allele, gray homozygotes for the other allele, and blue correspondingly heterozygotes who have one of each allele. The accumulation of diagnoses for *GSDMC (rs7814941, 8:129706613:G:A)* and *CHST3 (rs4284332, 10:719741194:C:T)* variants before the age of 30 can be seen in more detail in Fig. S6.

We found specific significant genetic correlations between LDH and 438 traits. The most significant positive genetic correlation in terms of smallest p-value was observed with the number of treatments/medications taken (Fig. 4: rg=0.52, pFDR=2.27^-100^), while the largest significant genetic correlation was observed with the diagnosis of dorsalgia (Fig. 4: rg=0.83, pFDR=1.23^-45^). LDH was also positively genetically correlated with other pain-related endpoints, such as neck and shoulder pain (Fig. 4: rg=0.58, pFDR=8.60^-95^) and knee pain (Fig. 4: rg=0.46, pFDR=2.07^-51^). The most significant negative genetic correlation of LDH was with a higher level of education (Fig. 4: rg=-0.41, pFDR=2.10^-88^).

**Fig. 4.**
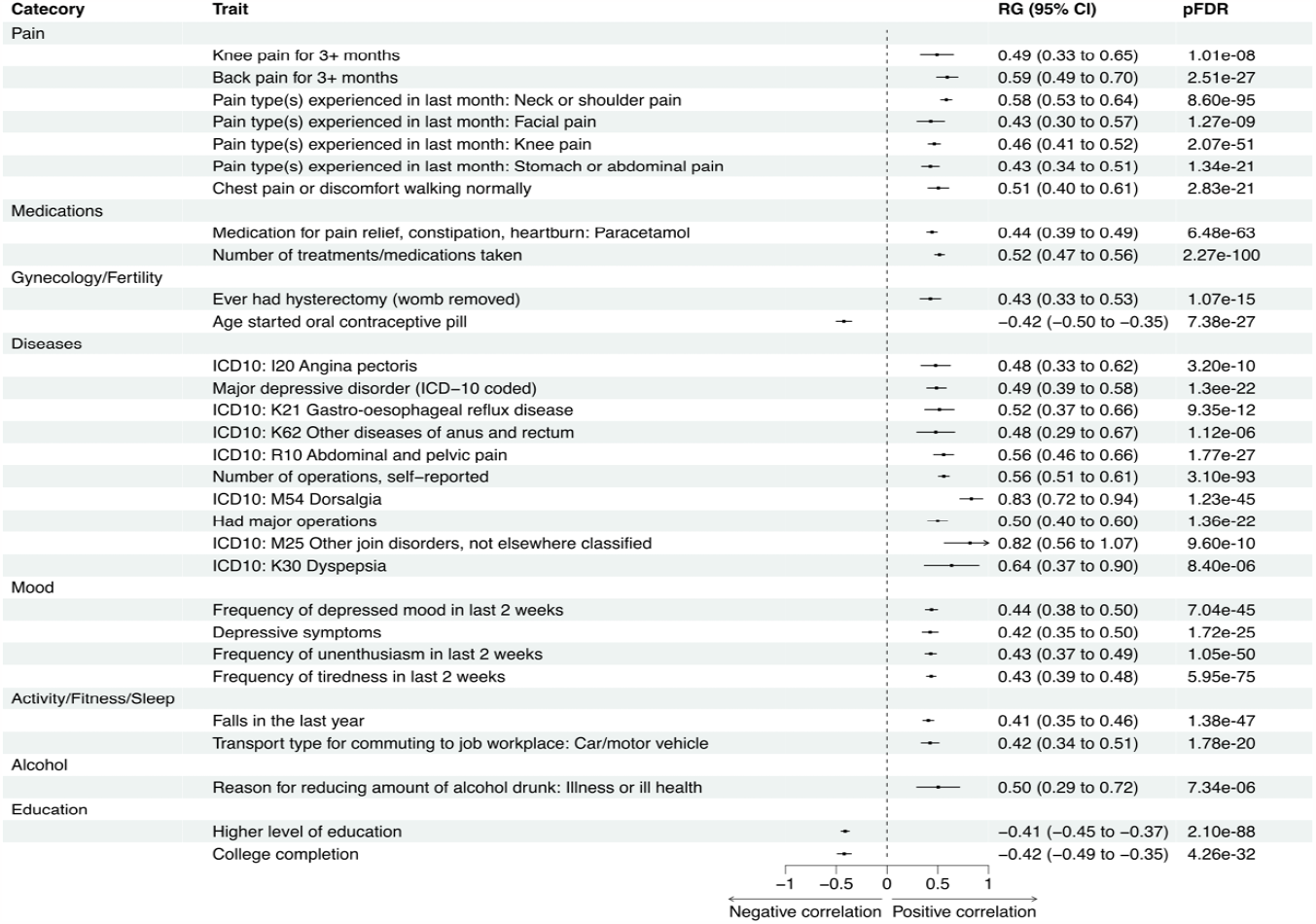
Genetic correlations were calculated using LDSC-software. All traits were extracted from the GWAS database provided by the MRC Integrative Epidemiology Unit (IEU). Only the strongest observed (rg< -0.4 & rg> 0.45) correlations with a significant false discovery corrected p-value than(p_FDR_ < 0.05) are shown in the figure. RG, genetic correlation coefficient value; pFDR, false discovery rate-corrected p-value. Genetic correlations for all 438 phenotypes can be seen in Table S7.

In Mendelian randomization, we uncovered potential causal relationship between several factors and LDH (Fig. 5, Table S8, Table S9). Results suggested a causal relationship between being overweight and higher LDH risk (OR=1.15, pFDR=0.002, Fig. 5, Fig. S8.1). Similarly, a potential causal relationship was observed between lumbar spine bone mineral density (LSBMD) and higher LDH risk (OR=1.15, pFDR=2.29^-5^, Fig. 5, Fig. S8.2). Both overweight and LSBMD are well-known risk factors for LDH, and both have been found to cause increased mechanical loading on the lumbar discs and vertebral endplates, affecting the pathogenesis of LDH^30–34^. A possible causal relationship was also observed between a higher level of education and lower LDH risk (OR=0.34, pFDR=4.35^-23^, Fig. 5, Fig. S8.3). In the previous studies, patients with lower socioeconomic status have been found to be more symptomatic, with worse pain outcomes, and more depression^35^. People with higher education usually have a better income level, they tend to seek treatment at an earlier stage after the onset of LDH, and they often have better pain management methods and generally healthier lifestyles^35,36^. Additionally, we noted a potential causal relationship between LDH and the frequency of tiredness in last two weeks (beta=0.02, pFDR=0.046, Fig. 5, Fig. S9.1) and back pain (beta=0.05, pFDR=6.20^-13^, Fig. 5, Fig. S9.2). The role of LDH to cause back pain is well known^4^, and the possible causal relationship we observed between LDH, and increased tiredness likely arises from sleeping problems, which are commonly reported by patients suffering from radicular pain^9^. These findings should be interpreted with caution, as even though we did not observe pleiotropy, some causal estimates were heterogeneous. In the leave-out analyses, all causal estimates were consistently in the same direction, so individual variants do not seem to drive the observed causal relationships (Fig. S10-11).

**Fig. 5.**
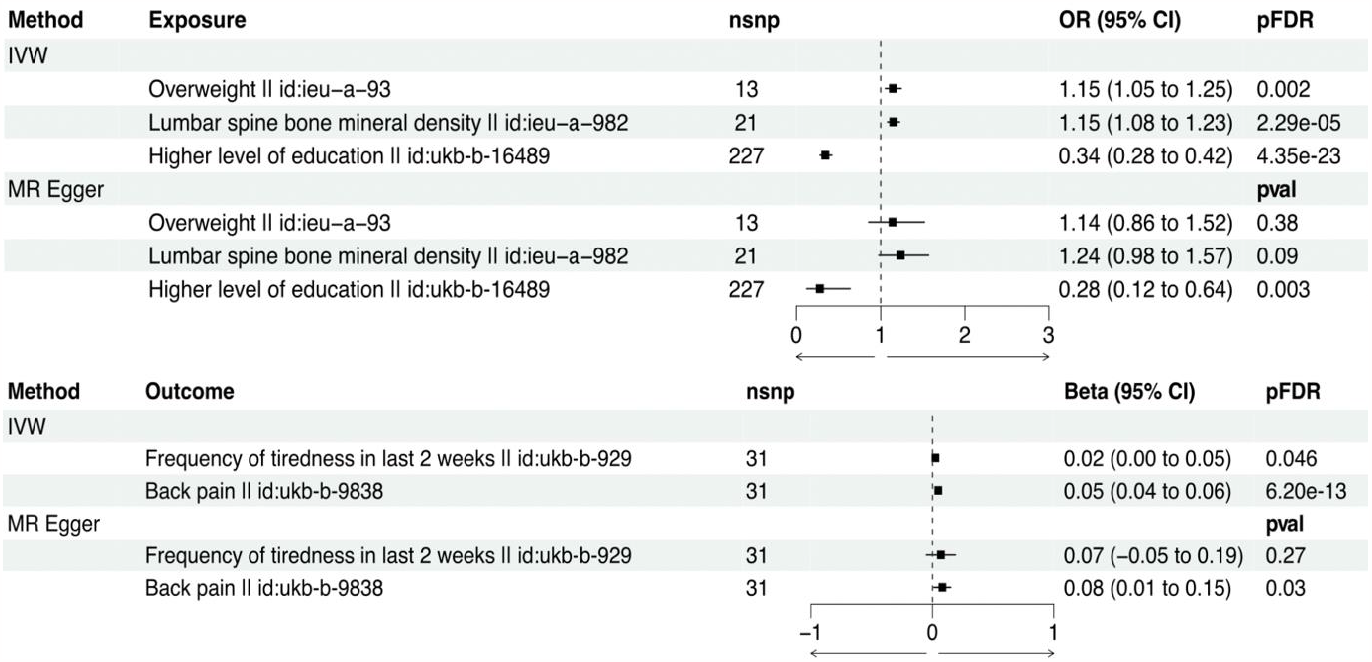
Exposures potentially causal for LDH (above), and outcomes that LDH was potentially found to be causal for (below). The analysis was performed using the TwoSampleMR R library and data from the present study and MRC-IEU database. Inverse variance weighted model was our primary analysis, for which statistical significance was considered at false discovery rate corrected p-value (pFDR < 0.05). As a sensitivity analysis, we also performed analysis by using MR Egger. nsnp, number of SNP’s; OR (95% CI), odds ratio and its 95% confidence interval; Beta (95% CI), beta estimate and it’s 95% confidence interval; pFDR, false discovery rate-corrected p-value.

The incorporation of data from three extensive biobanks enabled large sample size and facilitated discoveries of multiple genome-wide significant associations with LDH. Of note, our sample is limited to European ancestry only. Variations in the relative prevalence of LDH cases across the sample populations included in the meta-analysis, suggest potential discrepancies in how biobanks can identify LDH patients.

In conclusion, the novel LDH risk loci that we found expand the understanding of the hereditary causes of LDH. While changes in disc-related structures and inflammation-related factors play a major role in the etiology of LDH, our results suggest that nervous system-related mechanisms may also be implicated.

## Data Availability

Summary statistics will be made available through the NHGRI-EBI GWAS Catalog with GCSTxxxxxxx upon publication. The Finnish biobank data can be accessed through the Fingenious services (https://site.fingenious.fi/en/) managed by FINBB.

## Methods

### Study populations

#### FinnGen

The main goal of FinnGen (www.finngen.fi/en) is a better understanding of disease mechanisms by combining genomic and health data from up to over 500,000 Finns, with the aim of making healthcare and medical care more efficient. The aim of the studies is to find connections between individual genetic differences and diseases. The FinnGen project has the necessary ethical and prior permits for biobank research (Supplementary Note), and all persons who have provided a research sample are aware of the intended use of the samples and have given their written consent to biobank research either in connection with sample donation or when participating in older research projects, the materials of which have been transferred to Finnish biobanks with the written consent of Fimea.

*Tenth Revision* (ICD-10) codes were used to characterize phenotype M51 (M51.0-M51.9: Thoracic, thoracolumbar, and lumbosacral intervertebral disc disorders, Table S1). Patients without a record of these ICD codes were categorized as controls. The Hospital Discharge Registry and the Cause of Death Registry served as patient data sources; the analysis did not include patients whose registration data was only available in the primary care registry. The FinnGen (R9-version) data used in the study contains 37 636 LDH cases and 270 964 controls.

We also performed two sensitivity GWAS analyses in FinnGen using stricter case definitions. The first of the two additional case definitions included only LDH patients with the M51.1 code. There were 18 857 cases and 270 964 controls in the GWAS; LDH patients who also had other LDH codes were excluded from the analysis (Table S1). The second additional case definition included LDH patients who had undergone an LDH-related operation (NOMESCO version 1.15, ABC07, ABC16 & ABC26); this analysis included 7347 cases and controls of 270 964. LDH cases that had not been operated were excluded from the analysis. The same was done for operated cases that didn’t have a LDH diagnosis, as these patients were probably operated as a result of acute injury.

### Estonian biobank (EstBB)

The Estonian Biobank (www.genomics.ut.ee/en) cohort is a volunteer-based sample of the Estonian resident adult population (aged ≥18 years)^37^. Estonians represent 83%, Russians 14%, and other nationalities 3% of all participants. The current number of participants is > 205,000 and represents a large proportion, > 15 % of the Estonian adult population, making it ideally suited to population-based studies. General practitioners (GPs) and medical personnel in the special recruitment offices have recruited participants throughout the country. At baseline, the GPs performed a standardized health examination of the participants, who also donated blood samples for DNA, white blood cells and plasma tests and filled out a 16-module questionnaire on health-related topics such as lifestyle, diet and clinical diagnoses described in WHO ICD-10. A significant part of the cohort has whole genome sequencing (3000), whole exome sequencing (2500), genome-wide single nucleotide polymorphism (SNP) array data (200 000) and/or NMR metabolome data (200 000) available. In the meta-analysis, there were 34 035 LDH cases and 66 533 controls from the Estonian Biobank.

### UK biobank

The UK biobank (https://pan.ukbb.broadinstitute.org/) material consists of samples collected during the years 2006-2010. Samples were collected from hundreds of thousands of people aged 40–69 from all over Great Britain. We utilized the summary statistics from the PanUKBB project and used subset of European ancestry in the analysis that contained 9053 LDH patients and 411 478 controls from the UK biobank.

### Genotyping, imputation & quality control

#### FinnGen

Illumina and Affymetrix DNA microarrays were used to determine genotypes. Genotype data were quality controlled to exclude variants with a low Hardy-Weinberg equilibrium (HWE) p-value (<1x10^-6^), minor allele count (MAC) below three, and high missingness (cut-off 2%), as well as individuals with high genotype missingness (cut-off 5%), high levels of heterozygosity (±4 SD), non-Finnish ancestry, and individuals whose sex did not match the genotype data. Samples were pre phased using Eagle 2.3.5, with the number of conditioning haplotypes set to 20 000. Beagle 4.1 was used for genotype imputation. The reference panel was Finnish SISu v3, and the imputation protocol has been described at (dx.doi.org/10.17504/protocols.io.nmndc5e). Finally, post-imputation quality control was carried out to exclude variants with imputation information less than 0.6.

### EstBB

For genotyping, Illumina Human CoreExome, OmniExpress, 370CNV BeadChip and GSA arrays were used. Quality control included filtering on the basis of sample call rate (< 98%), heterozygosity (> mean ± 3SD), genotype and phenotype sex discordance, cryptic relatedness (IBD > 20%) and outliers from the European descent based on the MDS plot in comparison with HapMap reference samples. SNP quality filtering included call rate (<99%), MAF (<1%) and extreme deviation from Hardy–Weinberg equilibrium (P < 1 × 10−4). Imputation was performed using SHAPEIT2 for prephasing, the Estonian-specific reference panel^38^ and IMPUTE2^39^ with default parameters. Association testing was carried out with snptest-2.5.2, adjusting for 4 PCs, arrays, current age, and sex(when relevant). Individuals were excluded from the analysis if their call-rate was < 95% or sex defined using X chromosome heterozygosity estimates didn’t match phenotypic data. Variants with call-rate < 95%, MAF < 1% or HWE p-value < 1e-4 (autosomal variants only) and indels were excluded.

### GWAS

In FinnGen and in EstBB, GWAS using an additive genetic model was performed using the Regenie program^40^, adjusting each phenotype for age, sex, and the first 10 genetic principal components. The sensitivity analyses with stricter case definitions conducted in FinnGen were performed using Regenie and the same covariates as above. The goal of the sensitivity analyses was to evaluate whether general degeneration of IVD has an effect on the effect estimates of the lead variants observed in the meta-analysis, and if the effect estimates obtained in the original meta-analysis differ from the ones obtained using stricter case definitions. The aim was also to identify variants that could underlie LDH cases requiring surgery.

### Meta-analysis

A Python-based software was used for inverse-variance weighted meta-analysis (https://github.com/FINNGEN/META_ANALYSIS/). Variant data from the Estonian and UK biobanks were converted with from hg19 to hg38 prior to meta-analysis using the Picard liftover (http://broadinstitute.io/picard). In case of no exact match, matching was tried by flipping strand and or switching (EA->OA/OA->EA) for the EstBB and UKBB variants. If there were multiple variants in the same position, the exact match was favored. In total, there were 829 699 participants in the meta-analysis, of which there were 80 724 cases and 748 975 controls.

### Candidate gene characterization

We defined a locus as a window of 2MB (± 1,000,000 bases) containing at least one variant associated with LDH at P<5x10^-8^. We also performed conditional analyzes for the loci to identify possible secondary signals. The analyzes were performed with the GCTA software package^41^, and the lead variants detected from the loci were used as a covariate. For those loci where secondary signals were detected, the analysis was repeated based on the results of the first conditional analysis. In the analysis, the secondary signal detected in the first round was used as a covariate; however, no secondary signals were found in these analyses. For the loci that had not been reported in association with LDH in prior studies, we determined a potential candidate gene with a relevant biological function with the help of literature and databases (Genbank^42^, Uniprot^43^, GTEx-Portal^44^) and identified variants affecting gene regulation (eQTL).

### Heritability

The LDSC software^45^ was used to calculate the SNP-based heritability estimate. Heritability estimation was performed using the liability scale, with a sample prevalence of 0.097 and a population prevalence of 0.14 as estimated by Zhang et al. (2016)^3^. In FinnGen’s data, the sample prevalence was 0.122.

### Functional annotations

Functional annotations of the results of the meta-analysis were completed using FUMA^26^. Functional settings were selected for the analysis, in which case the program uses functional information for mapping. Positional mapping was also performed, and for that, SNP markers were selected for the region of exons or introns affecting post-transcriptional modifications and involved in gene regulation. Optional options included filtering SNP markers based on CADD results, which provided additional information on the possible harmful effects of SNP markers. In addition, filtering of SNP markers was performed based on the RegulomeDB results, and in turn, information was obtained based on gene expression data and epigenomics about the possible functions of SNP markers affecting gene regulation. In the mapping, gene expression data were also utilized, and eQTL mapping was performed. For this, we used whole blood (GTEx v8) as a tissue, and the focus was only on genes involved in protein-coding. A MAGMA analysis^25^, a functional association test, was also performed in the run, which focuses on gene-level information, unlike GWAS, where associations are reported at the variant level. MAGMA uses curated gene sets and GO annotations from MSigDB^27^ in the analyses. A 10kb gene window and selected GTEx v8 tissue variants were put into the analysis; the HLA region was also left out of the annotations.

### Survival analysis

With Kaplan-Meier’s, our aim was to observe how the LDH diagnoses accumulate for different variants according to age, and to evaluate whether there are differences in the accumulation between variants. Of the variants where differences in the accumulation of diagnoses were observed on the basis of the plots, we determined the exact age when the curve of the homozygote increasing the LDH risk statistically differed from the curves of other two genotypes of the same variant. The calculation was performed with the two-tailed test by using the survival rates and survival rate standard errors, which were obtained with the ‘survfit’ function, which is part of the ‘survival’ R library. P< 0.05 was used as the statistical cut-off value for a significant difference. Additionally, we calculated cumulative morbidity for every variant to further clarify whether some variants accumulate more diagnoses. Only FinnGen data was used for these analyses.

### Genetic correlations

Genetic correlations were calculated between LDH, and 438 other phenotypes extracted from the GWAS database provided by the MRC Integrative Epidemiology Unit (IEU) (https://gwas.mrcieu.ac.uk/). The LDSC software^45^ was used for these calculations. We used a false discovery rate (FDR)-corrected p-value (pFDR) < 0.05 as the limit for significant correlations.

### Mendelian randomization

For Mendelian randomization, we used the Two-Sample MR R library to conduct a bi-directional Mendelian randomization to examine the causal relationships between LDH and its associated risk factors. Risk factors related to lifestyle, pain, medication, and mood were included in the analysis (Table S10). Due to a bi-directional study approach, we were able to evaluate whether risk factors are causal for LDH and, consequently, if LDH is causal for risk factors. We obtained the LDH instruments from the FinnGen GWAS results since many of the GWAS data provided by the MRC-IEU are UKBB-based. This ensured that there was no overlap between the study populations. Variants correlated with each other were removed from the data so that only independent variants would be included in the analysis. For this, we used the default clumping settings (clumbing window 10 000kb, r^2^ 0.001). We run our primary analysis using the Inverse Variance Weighted (IVW) model. In the sensitivity analyses, we obtained MR Egger estimates. We also performed Cochran’s Q-test and the MR Egger intercept test to evaluate the heterogeneity and pleiotropy of the instruments. A leave-one-out analysis was also performed to see if there is a specific SNP driving a potentially observable causal relationship.

## Acknowledgements

E.S. was funded by Academy of Finland (grant number: 338229) and Orion Research Foundation sr. J.K. was funded by Sigrid Juselius foundation. We want to acknowledge the participants and investigators of FinnGen study. The FinnGen project is funded by two grants from Business Finland (HUS 4685/31/2016 and UH 4386/31/2016) and the following industry partners: AbbVie Inc., AstraZeneca UK Ltd, Biogen MA Inc., Bristol Myers Squibb (and Celgene Corporation & Celgene International II Sàrl), Genentech Inc., Merck Sharp & Dohme LCC, Pfizer Inc., GlaxoSmithKline Intellectual Property Development Ltd., Sanofi US Services Inc., Maze Therapeutics Inc., Janssen Biotech Inc, Novartis Pharma AG, and Boehringer Ingelheim International GmbH. Following biobanks are acknowledged for delivering biobank samples to FinnGen: Auria Biobank (www.auria.fi/biopankki), THL Biobank (www.thl.fi/biobank), Helsinki Biobank (www.helsinginbiopankki.fi), Biobank Borealis of Northern Finland (https://www.ppshp.fi/Tutkimus-ja-opetus/Biopankki/Pages/Biobank-Borealis-briefly-in-English.aspx), Finnish Clinical Biobank Tampere (www.tays.fi/en-US/Research_and_development/Finnish_Clinical_Biobank_Tampere), Biobank of Eastern Finland (www.ita-suomenbiopankki.fi/en), Central Finland Biobank (www.ksshp.fi/fi-FI/Potilaalle/Biopankki), Finnish Red Cross Blood Service Biobank (www.veripalvelu.fi/verenluovutus/biopankkitoiminta), Terveystalo Biobank (www.terveystalo.com/fi/Yritystietoa/Terveystalo-Biopankki/Biopankki/) and Arctic Biobank (https://www.oulu.fi/en/university/faculties-and-units/faculty-medicine/northern-finland-birth-cohorts-and-arctic-biobank). All Finnish Biobanks are members of BBMRI.fi infrastructure (www.bbmri.fi). Finnish Biobank Cooperative -FINBB (https://finbb.fi/) is the coordinator of BBMRI-ERIC operations in Finland. The Finnish biobank data can be accessed through the Fingenious^®^ services (https://site.fingenious.fi/en/) managed by FINBB.

This study was funded by European Union through the European Regional Development Fund Project No. 2014-2020.4.01.15-0012 GENTRANSMED and the Estonian Research Council Grant PUTs (PRG1911, PRG1291). Data analysis was carried out in part in the High-Performance Computing Center of University of Tartu. The activities of the EstBB are regulated by the Human Genes Research Act, which was adopted in 2000 specifically for the operations of the EstBB. Individual level data analysis in the EstBB was carried out under ethical approval [1.1-12/624] from the Estonian Committee on Bioethics and Human Research (Estonian Ministry of Social Affairs), using data according to release application [N04] from the Estonian Biobank.

