## Supplementary Materials for "Genome-wide meta-analysis conducted in three large biobanks expands the genetic landscape of lumbar disc herniations"

### LDH supplement

**Supplementary Note.** FinnGen DF9 Ethics statement
**Fig. S1.1** Regional plot of association on chr1 near *ALPL*
**Fig. S1.2** Regional plot of association on chr1 near *NGF*
**Fig. S1.3** Regional plot of association on chr1 near *TGFB2*
**Fig. S1.4** Regional plot of association on chr1 near *HHIPL2*
**Fig. S1.5** Regional plot of association on chr2 near *GPR1*
**Fig. S1.6** Regional plot of association on chr3 near *HYAL2*
**Fig. S1.7** Regional plot of association on chr3 near *PDZRN3*
**Fig. S1.8** Regional plot of association on chr3 near *ADCY5*
**Fig. S1.9** Regional plot of association on chr3 near *NCK1*
**Fig. S1.10** Regional plot of association on chr3 near *SHOX2*
**Fig. S1.11** Regional plot of association on chr4 near *IBSP*
**Fig. S1.12** Regional plot of association on chr5 near *HDAC3*
**Fig. S1.13** Regional plot of association on chr5 near *FGF18*
**Fig. S1.14** Regional plot of association on chr6 near *TRIM38*
**Fig. S1.15** Regional plot of association on chr6 near *HLA*
**Fig. S1.16** Regional plot of association on chr6 near *HLA*
**Fig. S1.17** Regional plot of association on chr6 near *TBX18*
**Fig. S1.18** Regional plot of association on chr7 near *ELN*
**Fig. S1.19** Regional plot of association on chr7 near *ZC3HC1*
**Fig. S1.20** Regional plot of association on chr7 near *STMP1*
**Fig. S1.21** Regional plot of association on chr9 near *TUSC1*
**Fig. S1.22** Regional plot of association on chr9 near *LPAR1*
**Fig. S1.23** Regional plot of association on chr9 near *DNM1*
**Fig. S1.24** Regional plot of association on chr10 near *AKR1C1*
**Fig. S1.25** Regional plot of association on chr10 near *HTRA1*
**Fig. S1.26** Regional plot of association on chr11 near *MYOEV*
**Fig. S1.27** Regional plot of association on chr11 near *SIK2*
**Fig. S1.28** Regional plot of association on chr12 near *GLI1*
**Fig. S1.29** Regional plot of association on chr12 near *KMT5A*
**Fig. S1.30** Regional plot of association on chr13 near *DIAPH3*
**Fig. S1.31** Regional plot of association on chr17 near *CA10*
**Fig. S1.32** Regional plot of association on chr18 near *NPC1*
**Fig. S1.33** Regional plot of association on chr18 near *SETBP1*
**Fig. S1.34** Regional plot of association on chr18 near *DCC*
**Fig. S1.35** Regional plot of association on chr19 near *TECR*
**Fig. S1.36** Regional plot of association on chr19 near *LENG8*
**Fig. S1.37** Regional plot of association on chr21 near *SLC5A3*
**Fig. S1.38** Regional plot of association on chrX near *PCYT1B*
**Fig. S1.39** Regional plot of association on chrX near *ITM2A*
**Fig. S1.40** Regional plot of association on chrX near *Empty*
**Fig. S1.41** Regional plot of association on chrX near *CHRDL1*
**Fig. S2** Beta estimates of lead variants in every dataset used in the study
**Fig. S3** Beta estimates of lead variants observed in meta-analysis and sensitivity analyses
**Fig. S4** Manhattan plot for meta-analysis and SURG GWAS
**Fig. S5** Results of MAGMA gene-based test and MAGMA tissue expression analysis
**Fig. S6** Kaplan-Meier plot for *GSDMC* and *CHST3* variants between ages 20-30
**Fig. S7** Kaplan-Meier plots with M51.1 diagnoses
**Fig. S8.1** Scatterplot for Overweight that could possibly be causal for LDH
**Fig. S8.2** Scatterplot for lumbar spine bone mineral density that could possibly be causal for LDH
**Fig. S8.3** Scatterplot for higher level of education that could possibly be causal for LDH
**Fig. S9.1** Scatterplot for frequency of tiredness in last 2 weeks for which LDH is potentially causal
**Fig. S9.2** Scatterplot for back pain for which LDH is potentially causal
**Fig. S10.1** Leave-one-out: Overweight ->LDH
**Fig. S10.2** Leave-one-out: Lumbar spine bone mineral density ->LDH
**Fig. S10.3** Leave-one-out: Higher level of education ->LDH
**Fig. S11.1** Leave-one-out: LDH -> Frequency of tiredness in last 2 weeks
**Fig. S11.2** Leave-one-out: LDH -> Back pain

**Table S1** The International Classification of Diseases codes used in phenotype characterization.
**Table S2** A list of lead variants at the 64 genome-wide significant loci that were associated with LDH in meta-analysis
**Table S3** Genomic locations and potential biological role of association signals
**Table S4** Effect differences between meta-analysis and SURG GWAS
**Table S5** Genome-wide significant ($p<5x10^{-8}$) lead variants associated with LDH related surgical operation
**Table S6** Cumulative morbidities and cumulative operations observed for every LDH associated variant
**Table S7** Results of genetic correlations for all 438 phenotypes
**Table S8** Potentially causal exposures for LDH
**Table S9** Outcomes that LDH is potentially causal
**Table S10** A list of risk factors studied in Mendelian randomization
**Table S11** A list of FinnGen authors and their affiliations
**Table S12** A list of Estonian Biobank Research team authors and their affiliations

**References**

**Supplementary Note.** FinnGen DF9 Ethics statement

Patients and control subjects in FinnGen provided informed consent for biobank research, based on the Finnish Biobank Act. Alternatively, separate research cohorts, collected prior the Finnish Biobank Act came into effect (in September 2013) and start of FinnGen (August 2017), were collected based on study-specific consents and later transferred to the Finnish biobanks after approval by Fimea (Finnish Medicines Agency), the National Supervisory Authority for Welfare and Health. Recruitment protocols followed the biobank protocols approved by Fimea. The Coordinating Ethics Committee of the Hospital District of Helsinki and Uusimaa (HUS) statement number for the FinnGen study is Nr HUS/990/2017.

The FinnGen study is approved by Finnish Institute for Health and Welfare (permit numbers: THL/2031/6.02.00/2017, THL/1101/5.05.00/2017, THL/341/6.02.00/2018, THL/2222/6.02.00/2018, THL/283/6.02.00/2019, THL/1721/5.05.00/2019 and THL/1524/5.05.00/2020), Digital and population data service agency (permit numbers: VRK43431/2017-3, VRK/6909/2018-3, VRK/4415/2019-3), the Social Insurance Institution (permit numbers: KELA 58/522/2017, KELA 131/522/2018, KELA 70/522/2019, KELA 98/522/2019, KELA 134/522/2019, KELA 138/522/2019, KELA 2/522/2020, KELA 16/522/2020), Findata permit numbers THL/2364/14.02/2020, THL/4055/14.06.00/2020,,THL/3433/14.06.00/2020, THL/4432/14.06/2020, THL/5189/14.06/2020, THL/5894/14.06.00/2020, THL/6619/14.06.00/2020, THL/209/14.06.00/2021, THL/688/14.06.00/2021, THL/1284/14.06.00/2021, THL/1965/14.06.00/2021, THL/5546/14.02.00/2020, THL/2658/14.06.00/2021, THL/4235/14.06.00/202, Statistics Finland (permit numbers: TK-53-1041-17 and TK/143/07.03.00/2020 (earlier TK-53-90-20) TK/1735/07.03.00/2021, TK/3112/07.03.00/2021) and Finnish Registry for Kidney Diseases permission/extract from the meeting minutes on 4$^{th}$ July 2019.

The Biobank Access Decisions for FinnGen samples and data utilized in FinnGen Data Freeze 9 include: THL Biobank BB2017_55, BB2017_111, BB2018_19, BB_2018_34, BB_2018_67, BB2018_71, BB2019_7, BB2019_8, BB2019_26, BB2020_1, Finnish Red Cross Blood Service Biobank 7.12.2017, Helsinki Biobank HUS/359/2017, HUS/248/2020, Auria Biobank AB17-5154 and amendment #1 (August 17 2020), AB20-5926 and amendment #1 (April 23 2020) and it´s modification (Sep 22 2021), Biobank Borealis of Northern Finland_2017_1013, Biobank of Eastern Finland 1186/2018 and amendment 22 § /2020, Finnish Clinical Biobank Tampere MH0004 and amendments (21.02.2020 & 06.10.2020), Central Finland Biobank 1-2017, and Terveystalo Biobank STB 2018001 and amendment 25$^{th}$ Aug 2020.

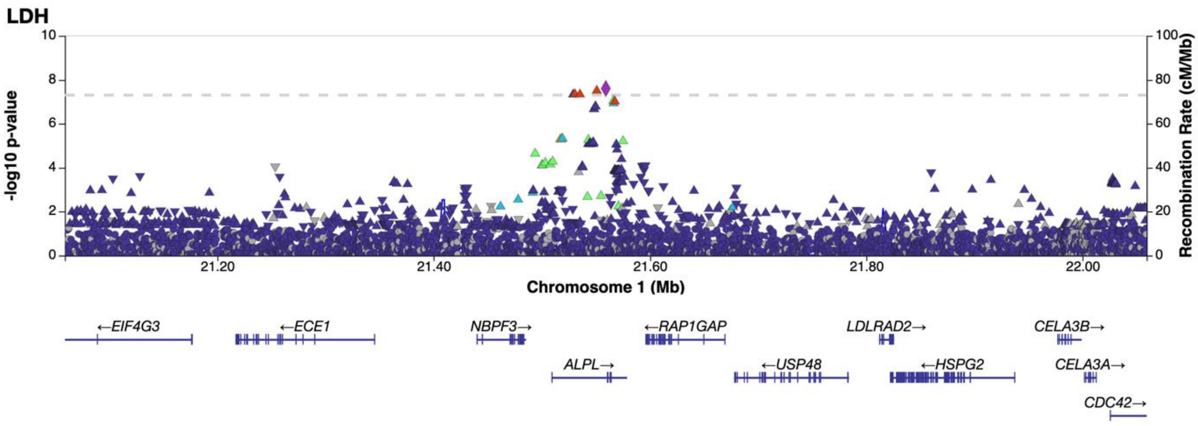


**Fig. S1.1** Regional association plot of novel LDH association on chromosome 1 limited to the area of 21059185-22059185. The association is based on the results of a meta-analysis. According to our research, the gene explaining the association of the locus is possibly *ALPL (alkaline phosphatase, biomineralization associated).*

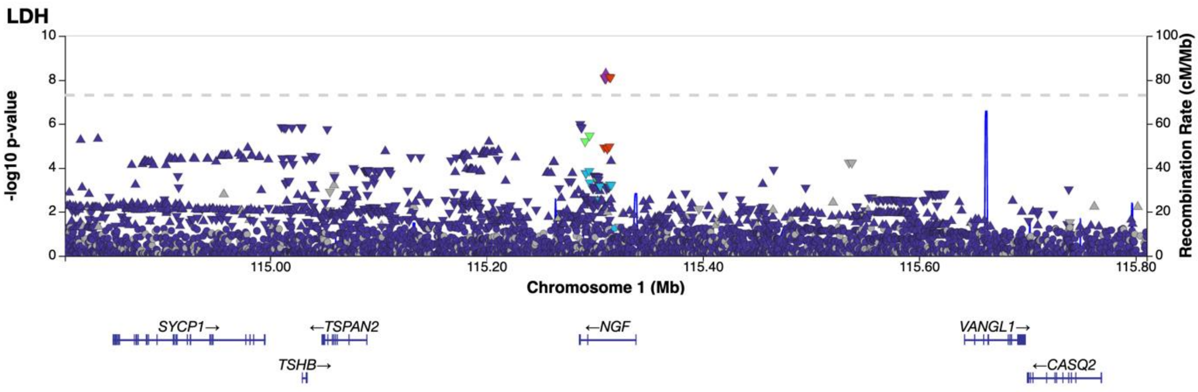


**Fig. S1.2** Regional association plot of novel LDH association on chromosome 1 limited to the area of 114810363-115810363. The association is based on the results of a meta-analysis. According to our research, the gene explaining the association of the locus is possibly *NGF (nerve growth factor).*

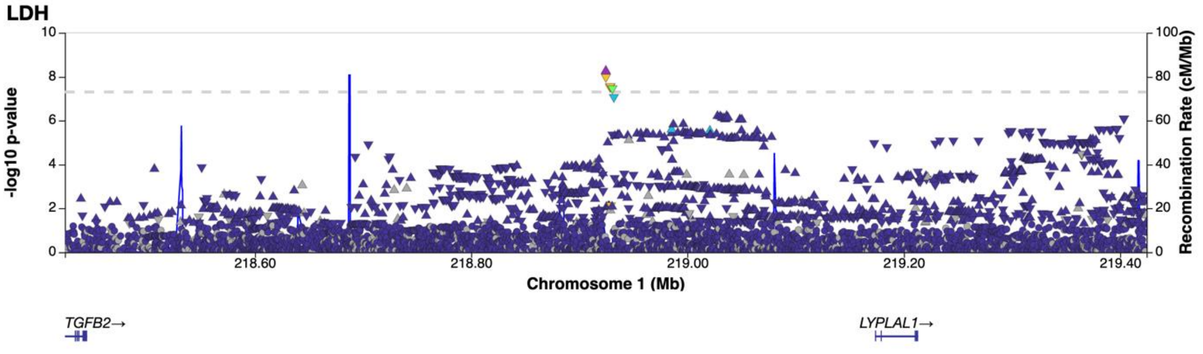


**Fig. S1.3** Regional association plot of novel LDH association on chromosome 1 limited to the area of 218424545-219424545. The association is based on the results of a meta-analysis. According to our research, the gene explaining the association of the locus is possibly *TGFB2 (transforming growth factor beta 2)*.

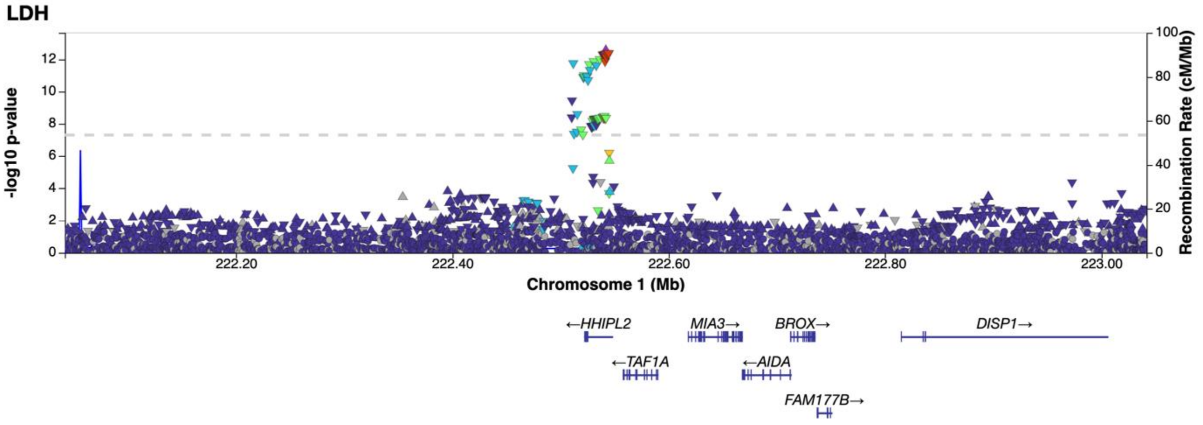


**Fig. S1.4** Regional association plot of novel LDH association on chromosome 1 limited to the area of 222041797-223041797. The association is based on the results of a meta-analysis. According to our research, the gene explaining the association of the locus is possibly *HHIPL2 (HHIP like 2)*.

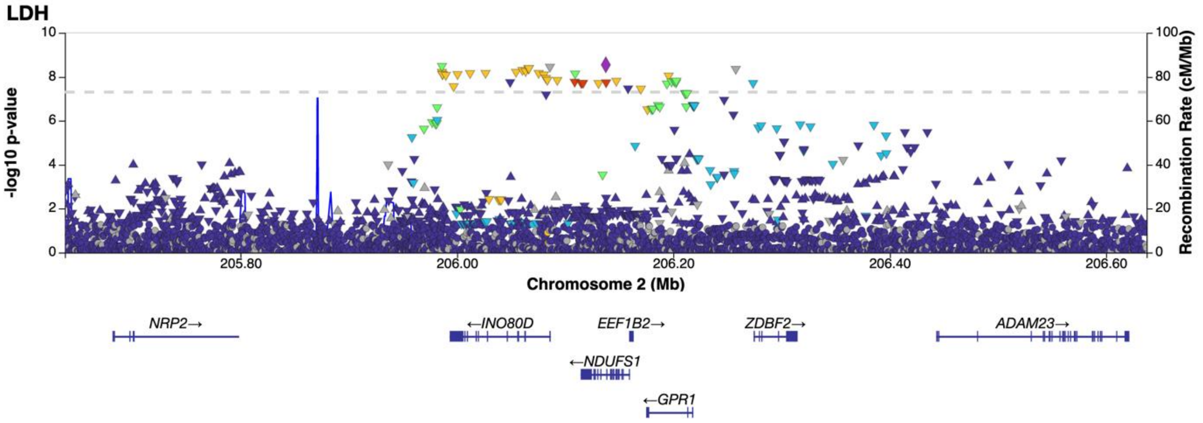


**Fig. S1.5** Regional association plot of novel LDH association on chromosome 2 limited to the area of 205637590-206637590. The association is based on the results of a meta-analysis. According to our research, the gene explaining the association of the locus is possibly *GPR1 (G-protein coupled receptor 1),* also known as *CMKFR2 (chemerin like receptor 2).*

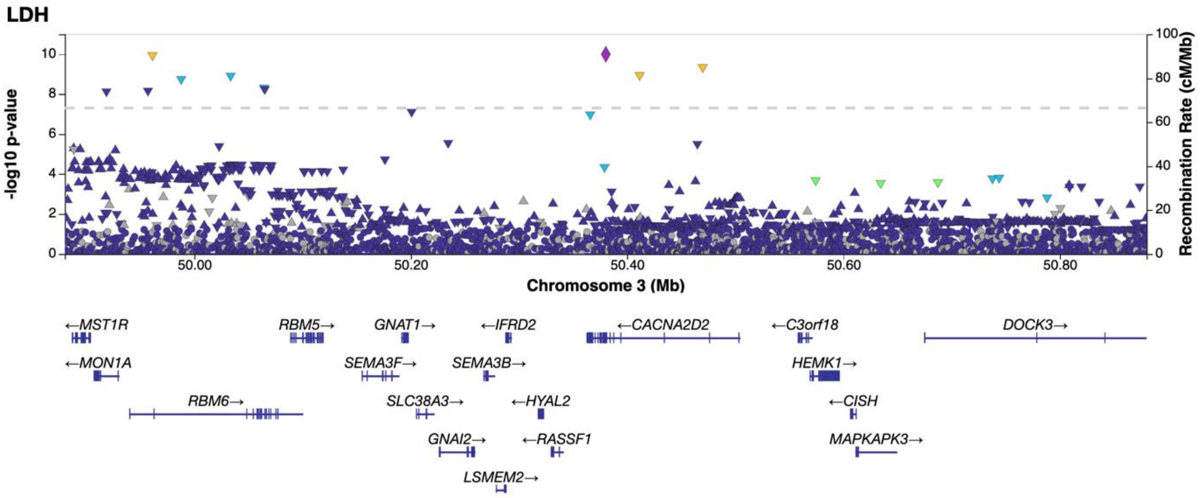


**Fig. S1.6** Regional association plot of novel LDH association on chromosome 3 limited to the area of 49880254-50880254. The association is based on the results of a meta-analysis. According to our research, the gene explaining the association of the locus is possibly *HYAL2 (hyaluronidase 2).*

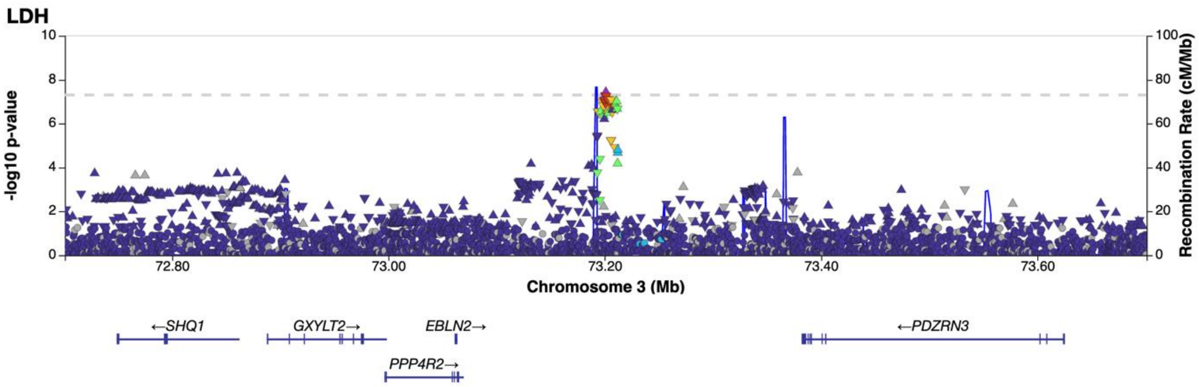


**Fig. S1.7** Regional association plot of novel LDH association on chromosome 3 limited to the area of 72700973-73700973. The association is based on the results of a meta-analysis. According to our research, the gene explaining the association of the locus is possibly *PDZRN3 (PDZ domain containing ring finger 3).*

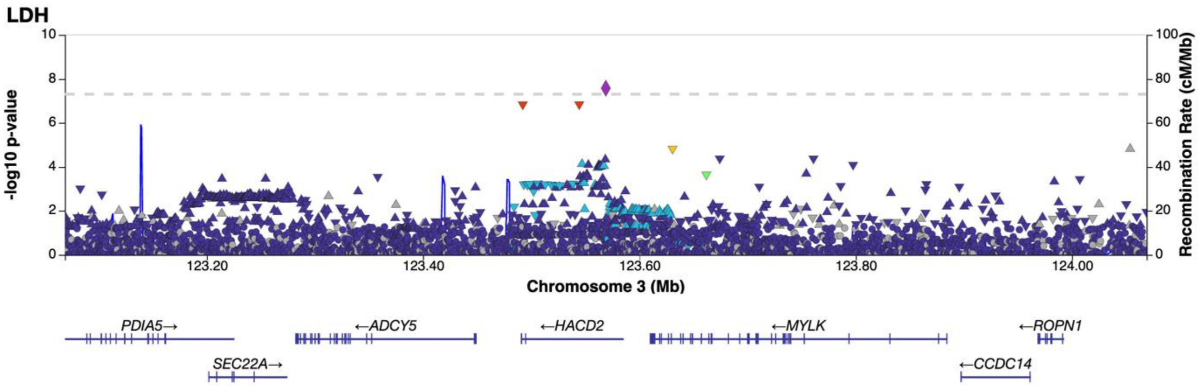


**Fig. S1.8** Regional association plot of novel LDH association on chromosome 3 limited to the area of 123068959-124068959. The association is based on the results of a meta-analysis. According to our research, the gene explaining the association of the locus is possibly *ADCY5 (adenylate cyclase 5).*

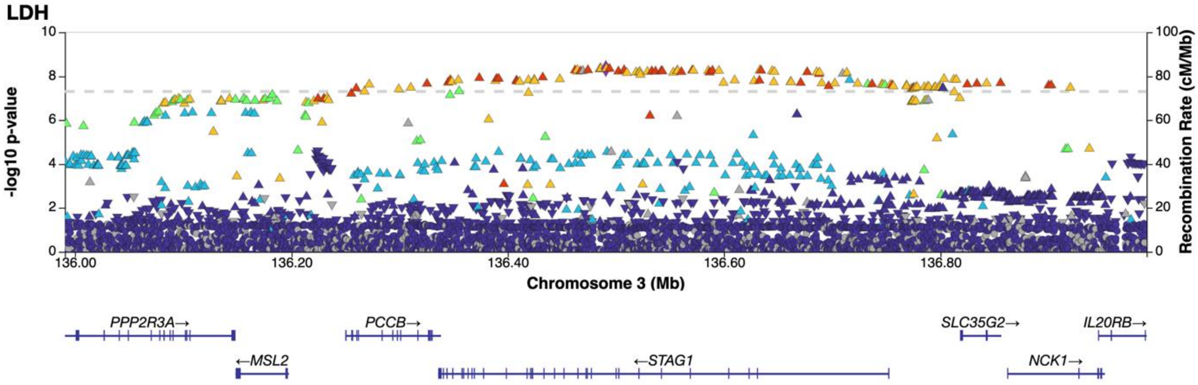


**Fig. S1.9** Regional association plot of novel LDH association on chromosome 3 limited to the area of 135990708-136990708. The association is based on the results of a meta-analysis. According to our research, the gene explaining the association of the locus is possibly *NCK1 (NCK adaptor protein 1).*

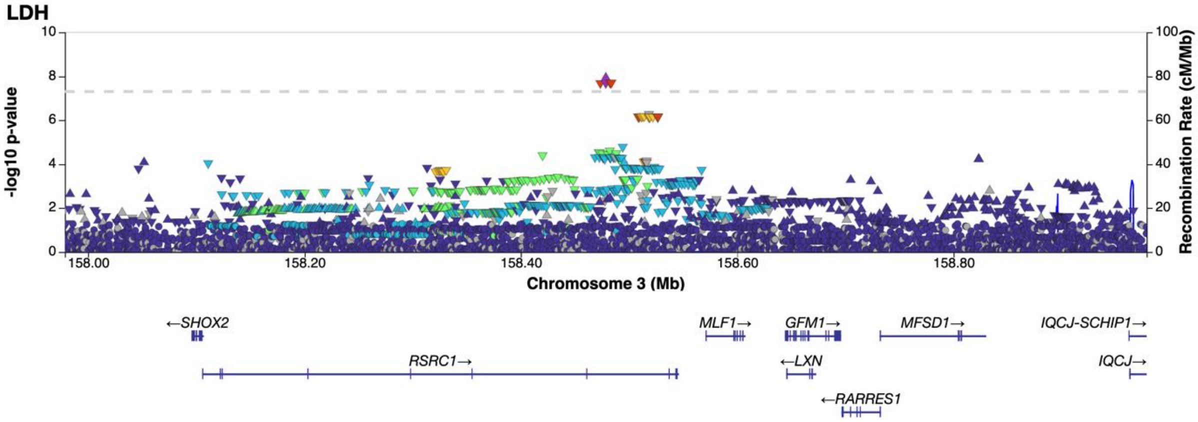


**Fig. S1.10** Regional association plot of novel LDH association on chromosome 3 limited to the area of 157978549-158978549. The association is based on the results of a meta-analysis. According to our research, the gene explaining the association of the locus is possibly *SHOX2 (short stature homeobox 2).*

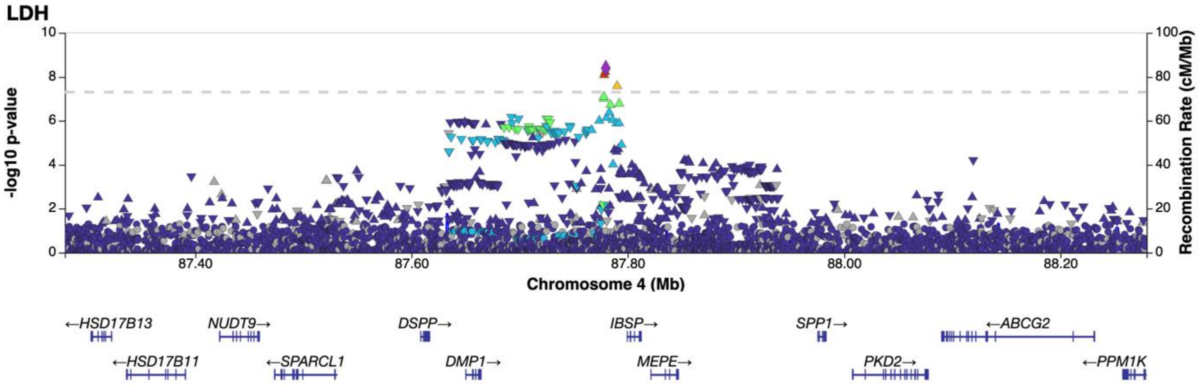


**Fig. S1.11** Regional association plot of novel LDH association on chromosome 4 limited to the area of 87279677-88279677. The association is based on the results of a meta-analysis. According to our research, the gene explaining the association of the locus is possibly *IBSP (integrin binding sialoprotein).*

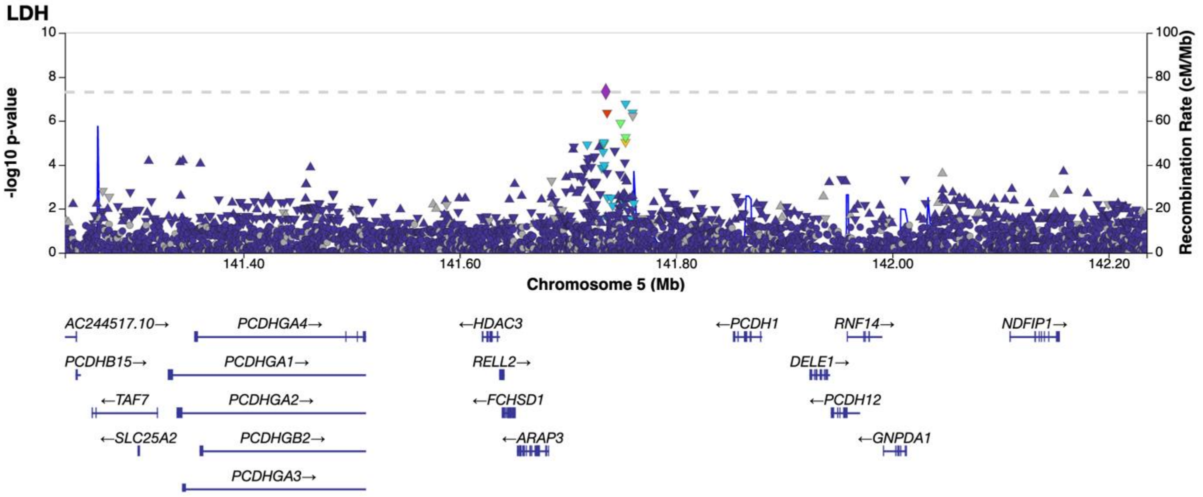


**Fig. S1.12** Regional association plot of novel LDH association on chromosome 5 limited to the area of 141235121-142235121. The association is based on the results of a meta-analysis. According to our research, the gene explaining the association of the locus is possibly *HDAC3 (histone deacetylase 3).*

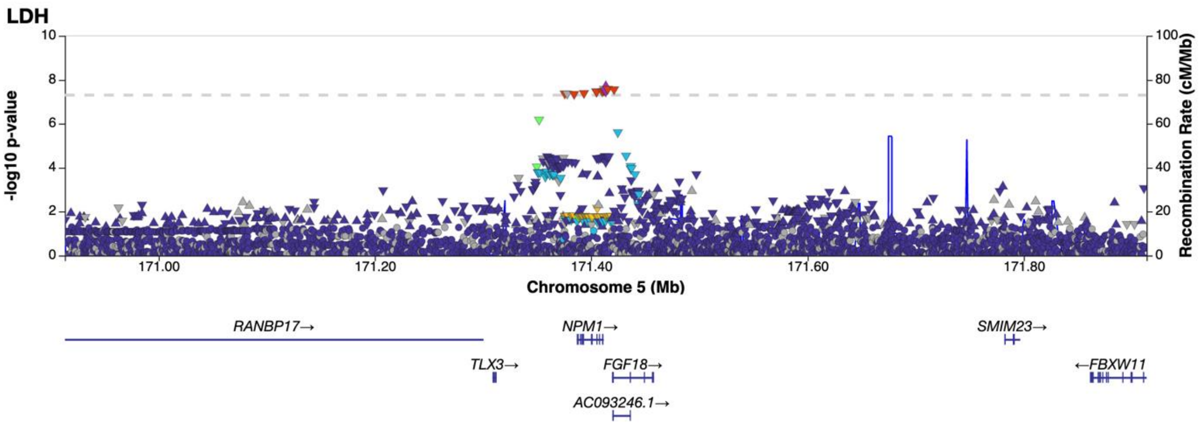


**Fig. S1.13** Regional association plot of novel LDH association on chromosome 5 limited to the area of 170913500-171913500. The association is based on the results of a meta-analysis. According to our research, the gene explaining the association of the locus is possibly *FGF18 (fibroblast growth factor 18).*

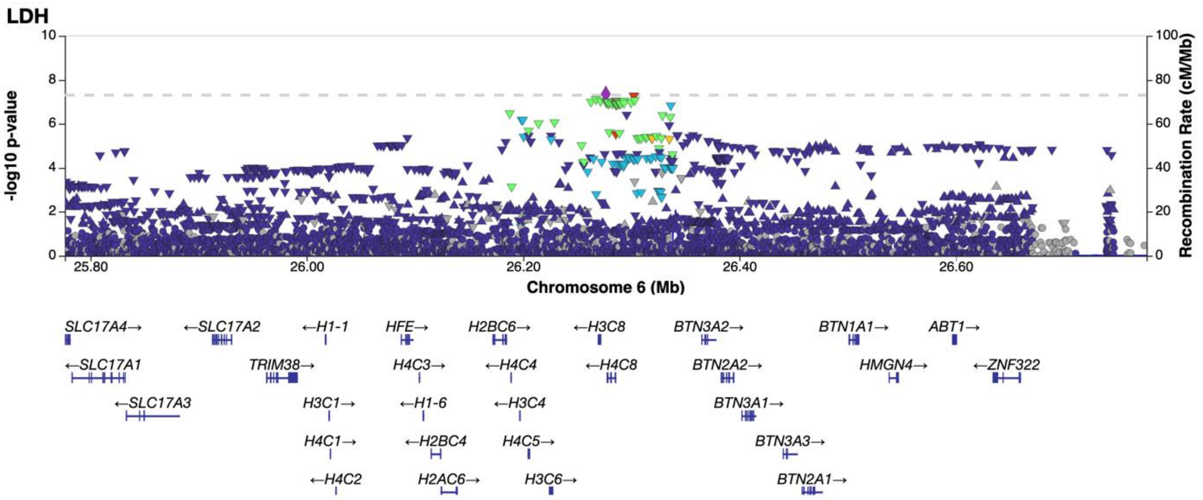


**Fig. S1.14** Regional association plot of novel LDH association on chromosome 6 limited to the area of 25776422-26776422. The association is based on the results of a meta-analysis. According to our research, the gene explaining the association of the locus is possibly *TRIM38 (tripartite motif containing 38).*

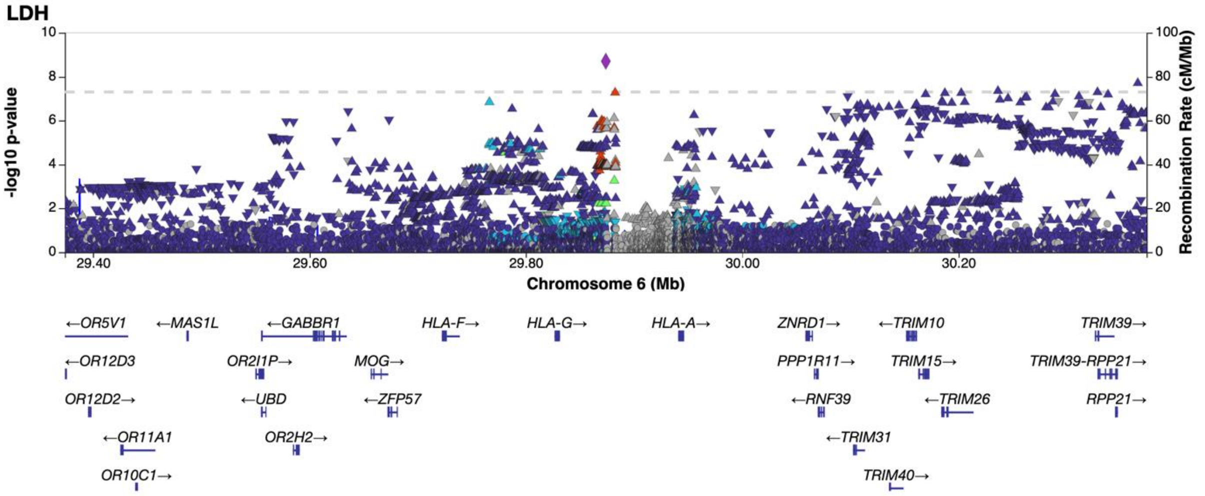


**Fig. S1.15** Regional association plot of novel LDH association on chromosome 6 limited to the area of 29373925-30373925. The association is based on the results of a meta-analysis. According to our research, the gene explaining the association of the locus is possibly *HLA*.

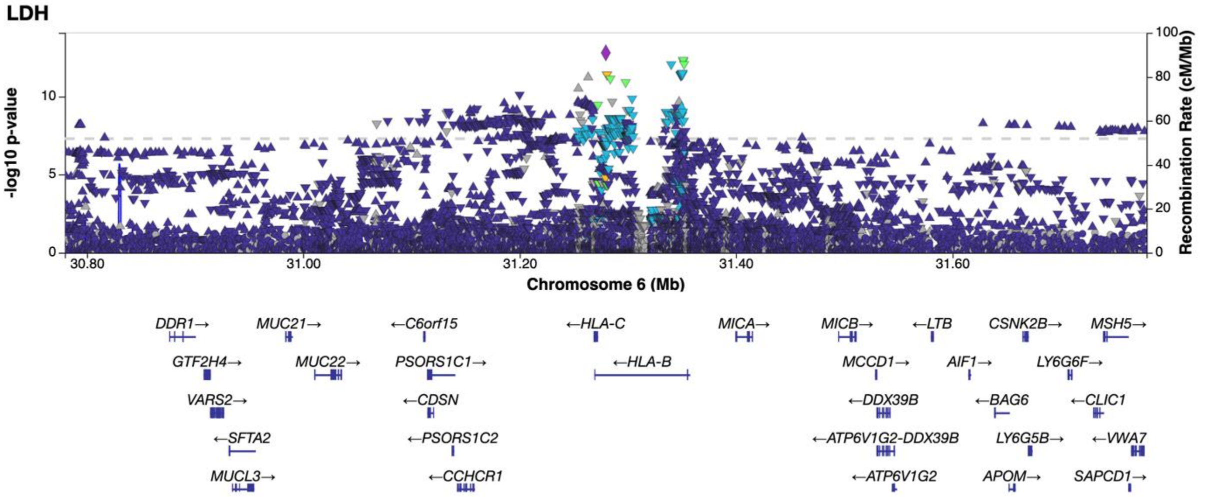


**Fig. S1.16** Regional association plot of novel LDH association on chromosome 6 limited to the area of 30779637-31779637. The association is based on the results of a meta-analysis. According to our research, the gene explaining the association of the locus is possibly *HLA*.

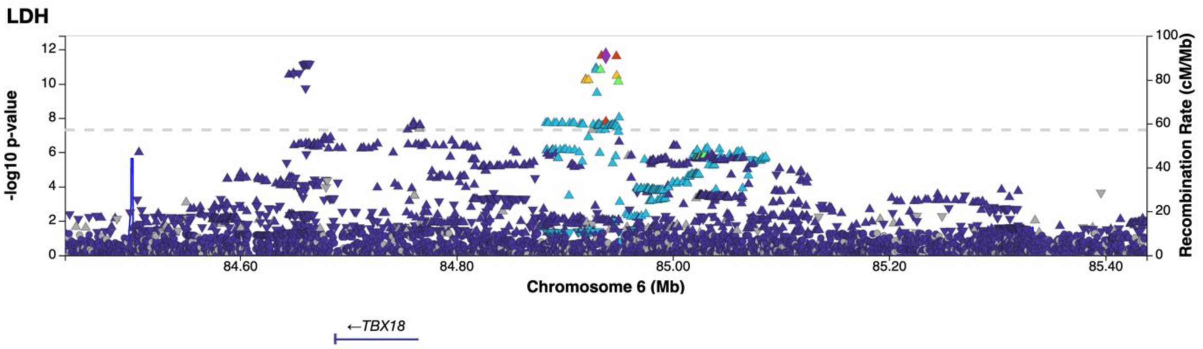


**Fig. S1.17** Regional association plot of novel LDH association on chromosome 6 limited to the area of 84438145-85438145. The association is based on the results of a meta-analysis. According to our research, the gene explaining the association of the locus is possibly *TBX18 (T-box transcription factor 18).*

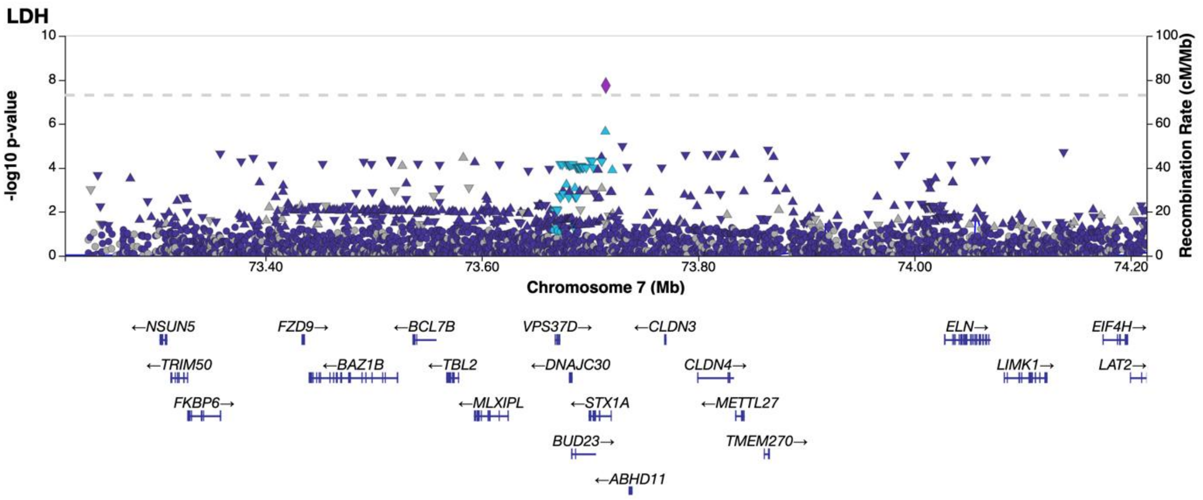


**Fig. S1.18** Regional association plot of novel LDH association on chromosome 7 limited to the area of 73214641-74214641. The association is based on the results of a meta-analysis. According to our research, the gene explaining the association of the locus is possibly *ELN (elastin).*

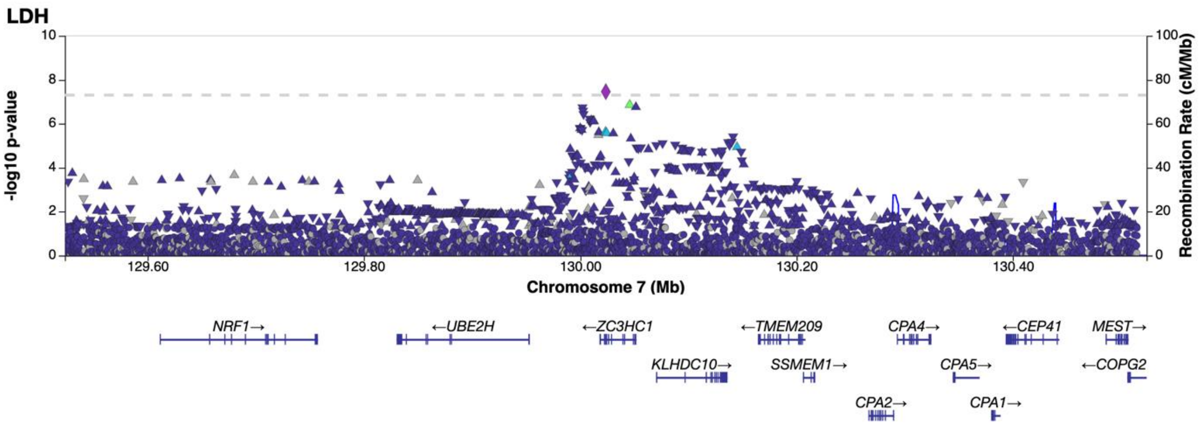


**Fig. S1.19** Regional association plot of novel LDH association on chromosome 7 limited to the area of 129523656-130523656. The association is based on the results of a meta-analysis. According to our research, the gene explaining the association of the locus is possibly *ZC3HC1 (zinc finger C3HC-type containing 1).*

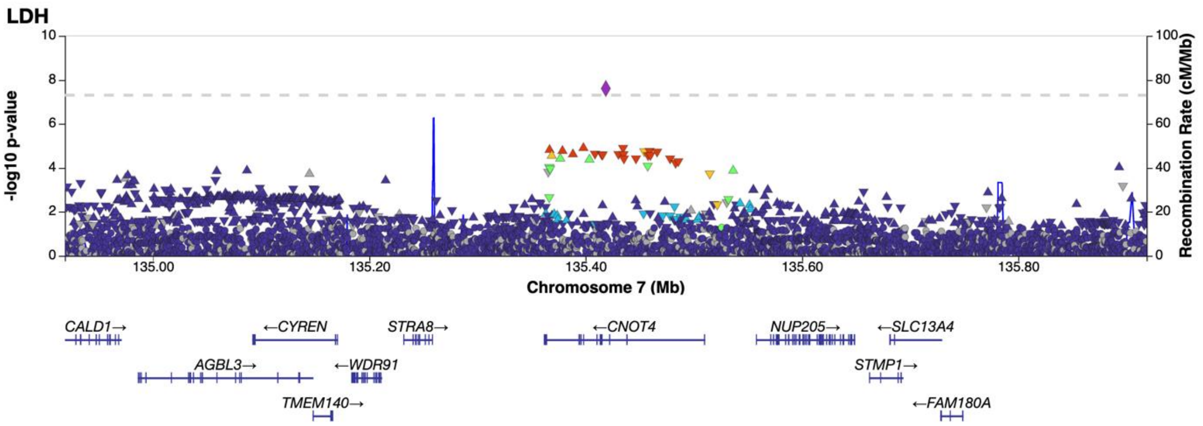


**Fig. S1.20** Regional association plot of novel LDH association on chromosome 7 limited to the area of 134918700-135918700. The association is based on the results of a meta-analysis. According to our research, the gene explaining the association of the locus is possibly *STMP1 (short transmembrane mitochondrial protein 1).*

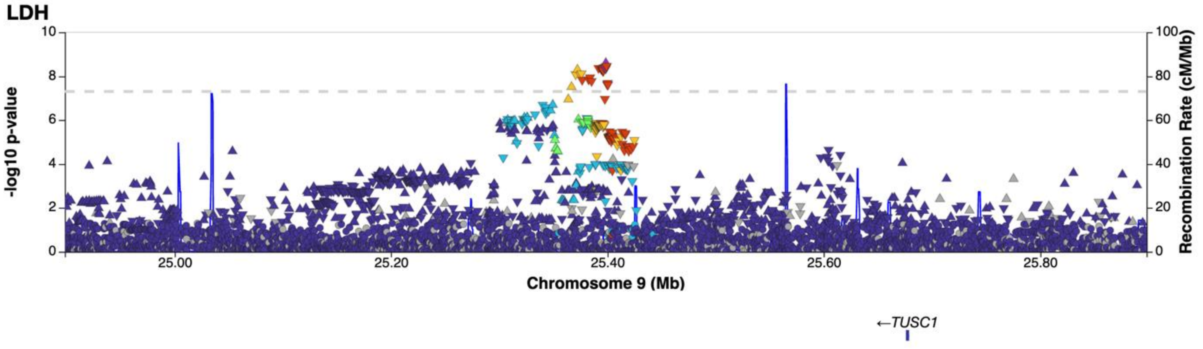


**Fig. S1.21** Regional association plot of novel LDH association on chromosome 9 limited to the area of 24898495-25898495. The association is based on the results of a meta-analysis. According to our research, the gene explaining the association of the locus is possibly *TUSC1 (tumor suppressor candidate 1).*

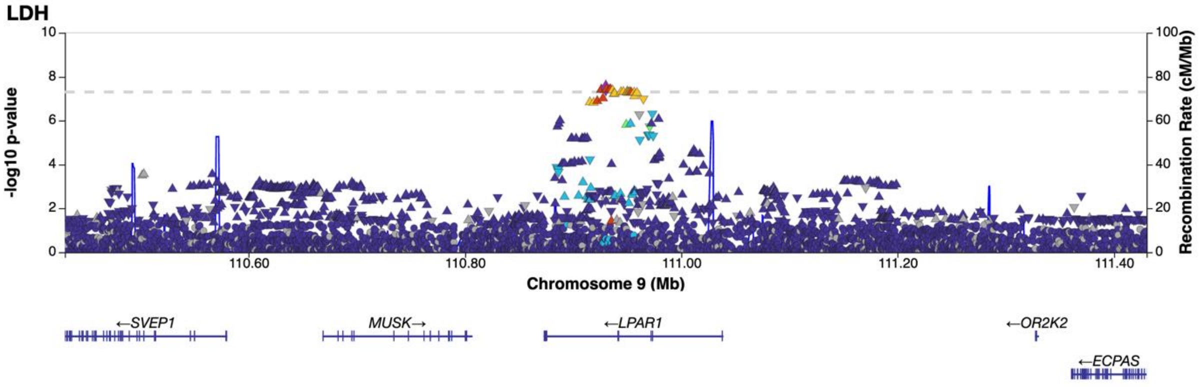


**Fig. S1.22** Regional association plot of novel LDH association on chromosome 9 limited to the area of 110430238-111430238. The association is based on the results of a meta-analysis. According to our research, the gene explaining the association of the locus is possibly *LPAR1 (lysophosphatidic acid receptor 1).*

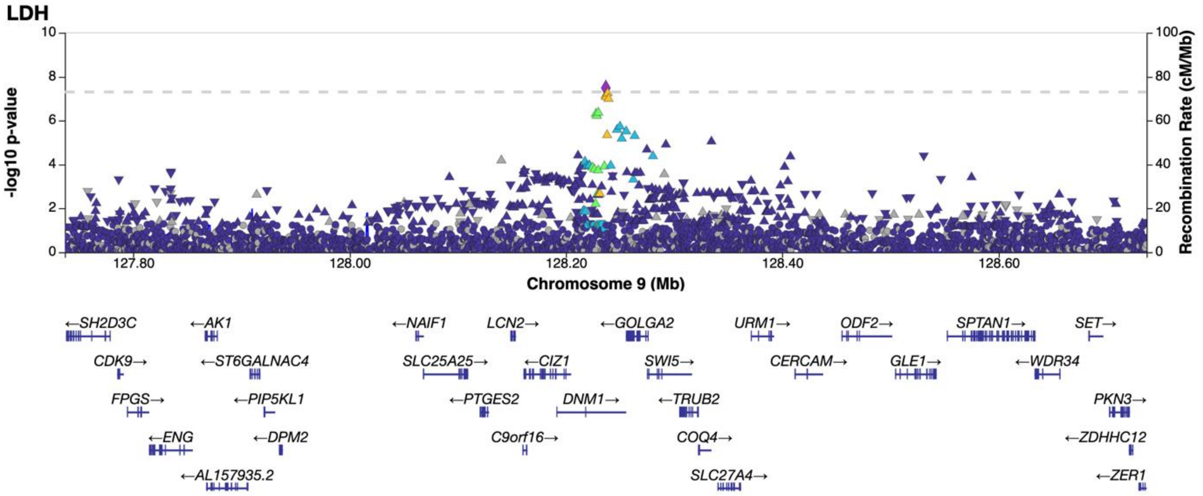


**Fig. S1.23** Regional association plot of novel LDH association on chromosome 9 limited to the area of 127736873-128736873. The association is based on the results of a meta-analysis. According to our research, the gene explaining the association of the locus is possibly *DNM1 (dynamin 1).*

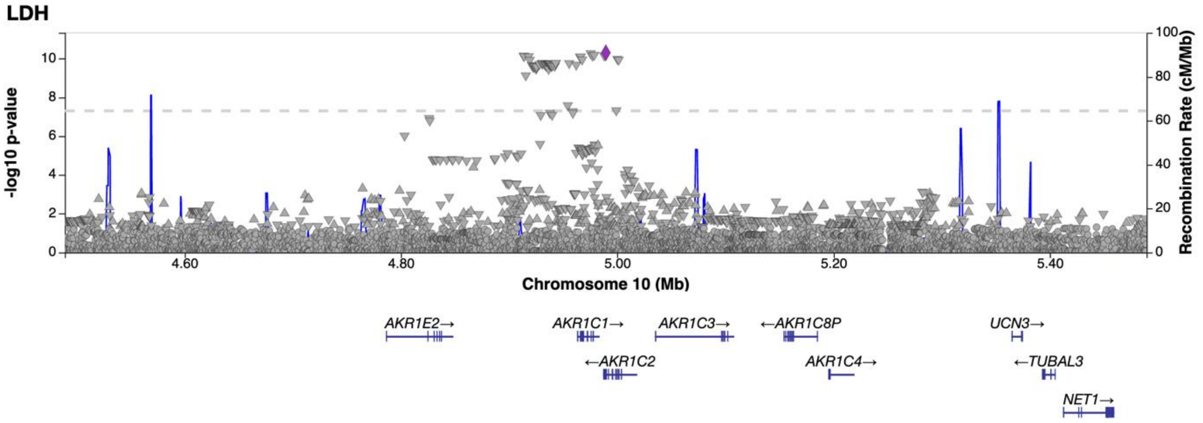


**Fig. S1.24** Regional association plot of novel LDH association on chromosome 10 limited to the area of 4489436-5489436. The association is based on the results of a meta-analysis. According to our research, the gene explaining the association of the locus is possibly *AKR1C1 (aldo-keto reductase family 1 member C1).*

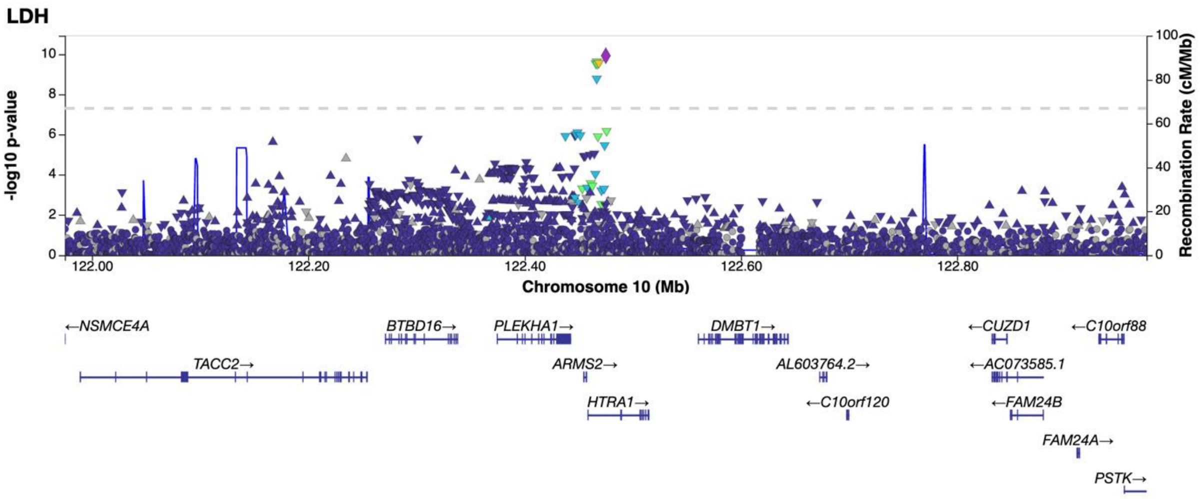


**Fig. S1.25** Regional association plot of novel LDH association on chromosome 10 limited to the area of 121975088-122975088. The association is based on the results of a meta-analysis. According to our research, the gene explaining the association of the locus is possibly *HTRA1 (HtrA serine peptidase 1).*

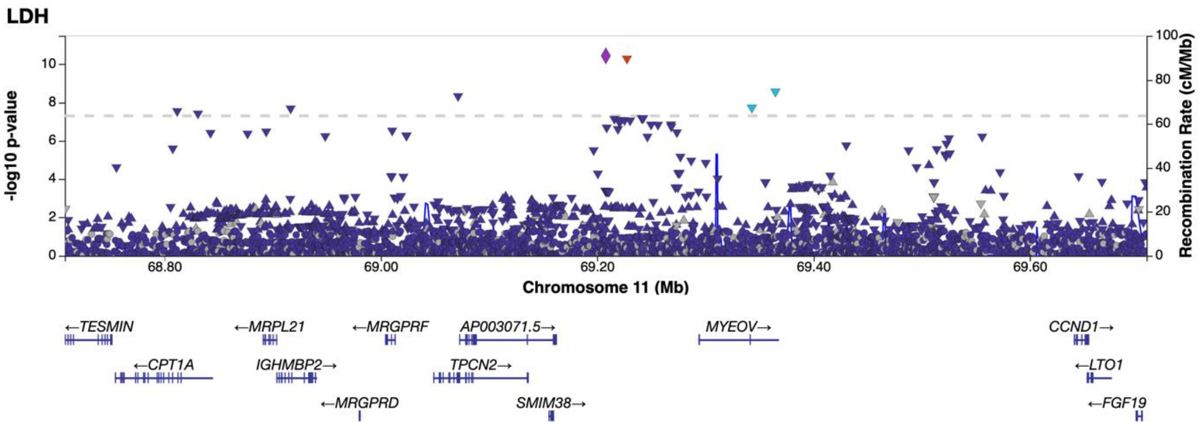


**Fig. S1.26** Regional association plot of novel LDH association on chromosome 11 limited to the area of 68708032-69708032. The association is based on the results of a meta-analysis. According to our research, the gene explaining the association of the locus is possibly *MYEOV (myeloma overexpressed).*

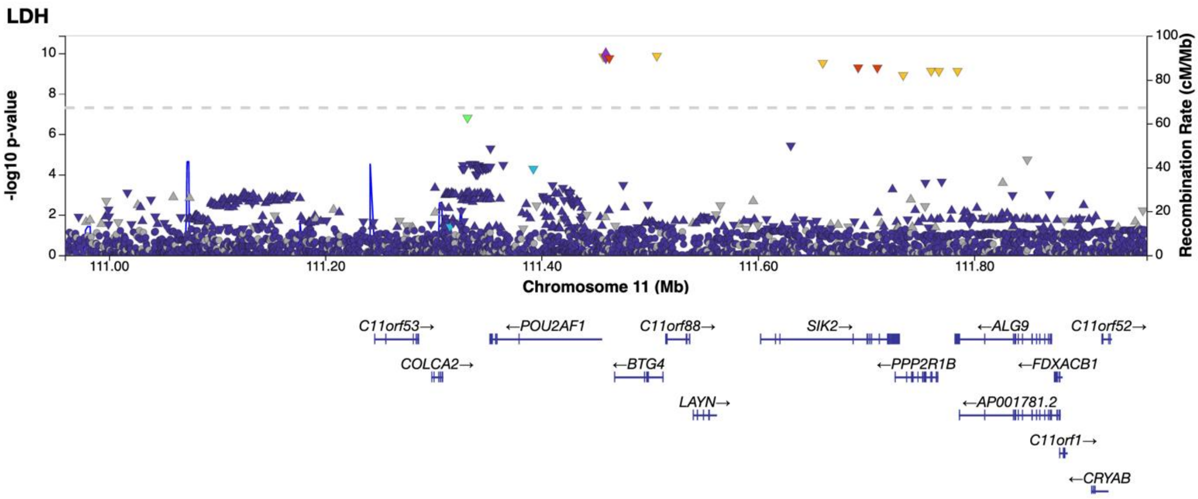


**Fig. S1.27** Regional association plot of novel LDH association on chromosome 11 limited to the area of 110959420-111959420. The association is based on the results of a meta-analysis. According to our research, the gene explaining the association of the locus is possibly *SIK2 (salt inducible kinase 2).*

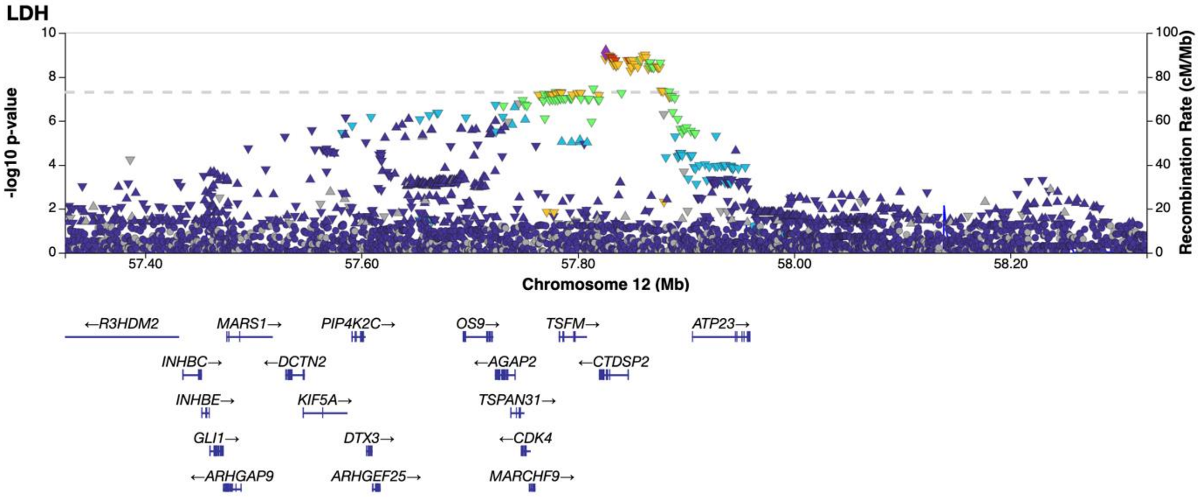


**Fig. S1.28** Regional association plot of novel LDH association on chromosome 12 limited to the area of 57325898-58325898. The association is based on the results of a meta-analysis. According to our research, the gene explaining the association of the locus is possibly *GLI1 (GLI family zinc finger 1).*

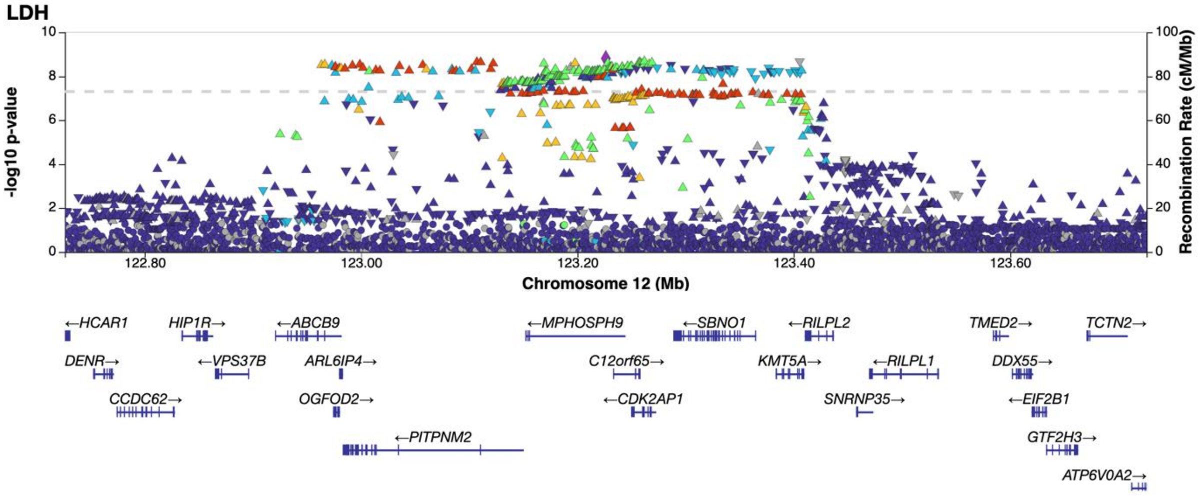


**Fig. S1.29** Regional association plot of novel LDH association on chromosome 12 limited to the area of 122726288-123726288. The association is based on the results of a meta-analysis. According to our research, the gene explaining the association of the locus is possibly *KMT5A (lysine methyltransferase 5A).*

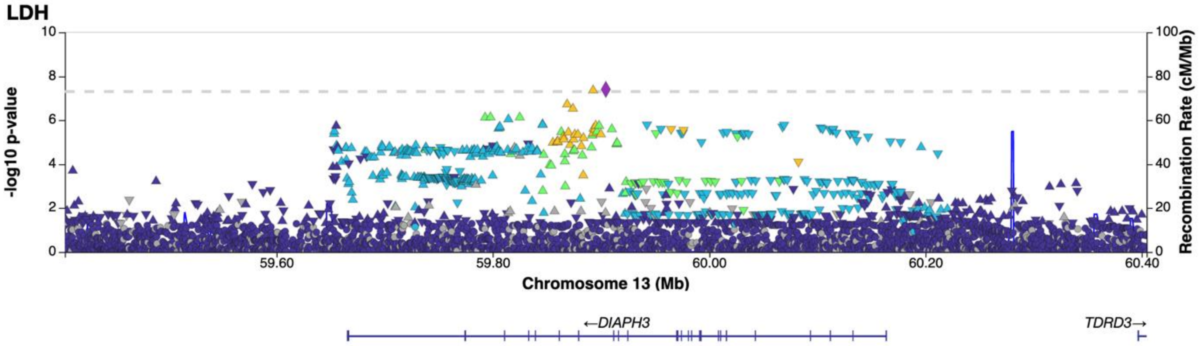


**Fig. S1.30** Regional association plot of novel LDH association on chromosome 13 limited to the area of 59404471-60404471. The association is based on the results of a meta-analysis. According to our research, the gene explaining the association of the locus is possibly *DIAPH3 (diaphanous-related formin 3).*

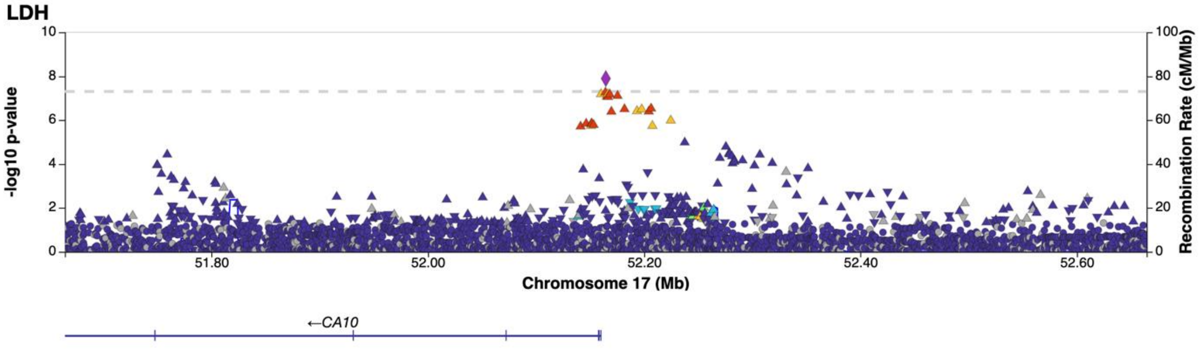


**Fig. S1.31** Regional association plot of novel LDH association on chromosome 17 limited to the area of 51664544-52664544. The association is based on the results of a meta-analysis. According to our research, the gene explaining the association of the locus is possibly *CA10 (carbonic anhydrase 10).*

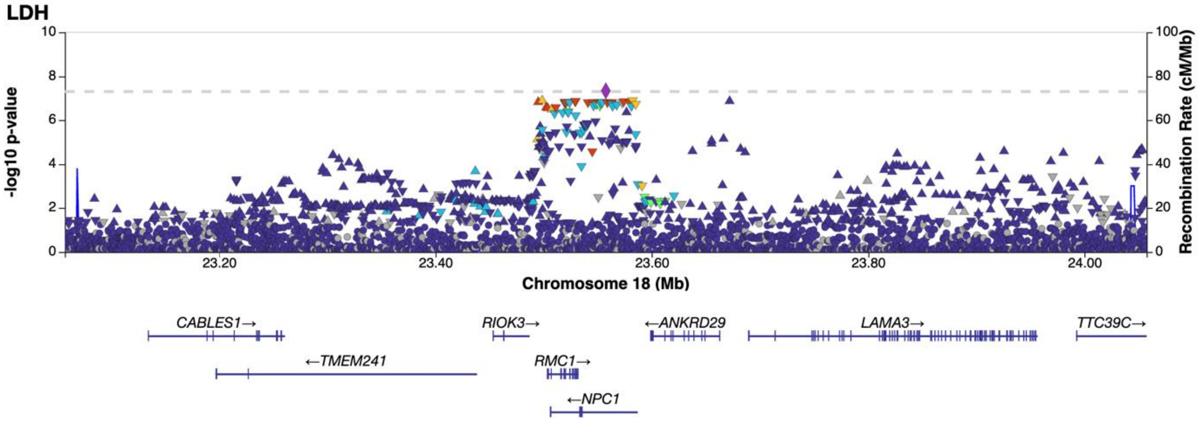


**Fig. S1.32** Regional association plot of novel LDH association on chromosome 18 limited to the area of 23057478-24057478. The association is based on the results of a meta-analysis. According to our research, the gene explaining the association of the locus is possibly *NPC1 (NPC intracellular cholesterol transporter 1).*

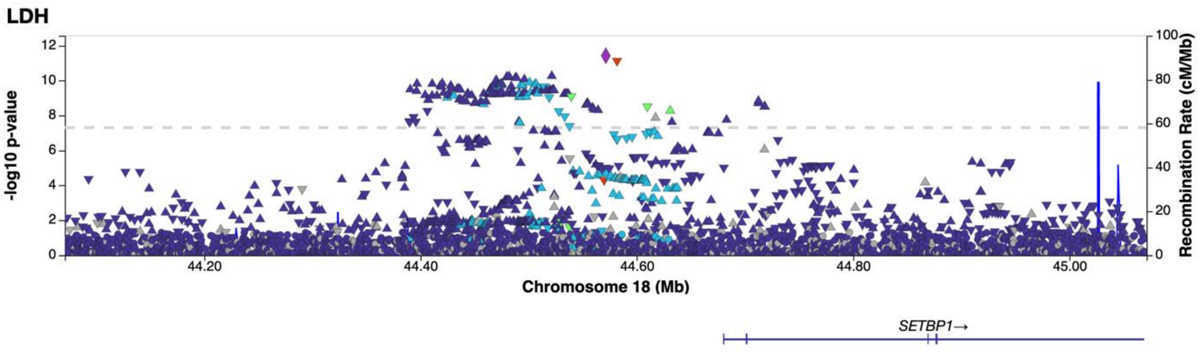


**Fig. S1.33** Regional association plot of novel LDH association on chromosome 18 limited to the area of 44071296-45071296. The association is based on the results of a meta-analysis. According to our research, the gene explaining the association of the locus is possibly *SETBP1 (SET binding protein 1).*

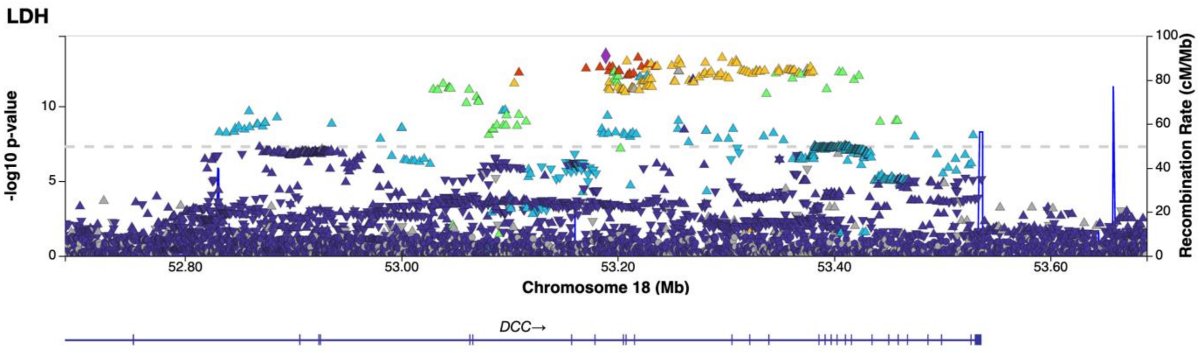


**Fig. S1.34** Regional association plot of novel LDH association on chromosome 18 limited to the area of 52689047-53689047. The association is based on the results of a meta-analysis. According to our research, the gene explaining the association of the locus is possibly *DCC (DCC netrin 1 receptor).*

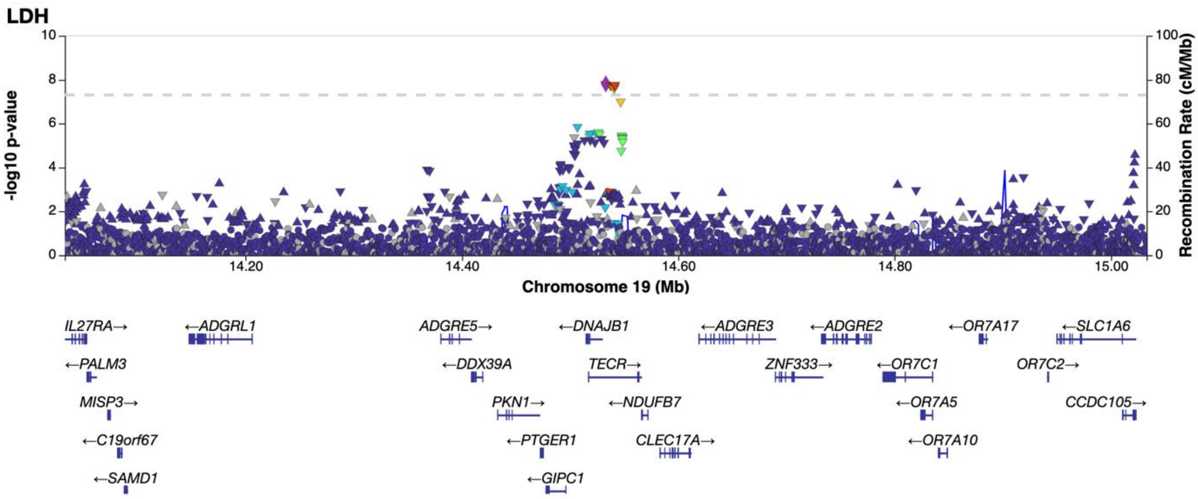


**Fig. S1.35** Regional association plot of novel LDH association on chromosome 19 limited to the area of 14033044-15033044. The association is based on the results of a meta-analysis. According to our research, the gene explaining the association of the locus is possibly *TECR (trans-2,3-enoyl-CoA reductase).*

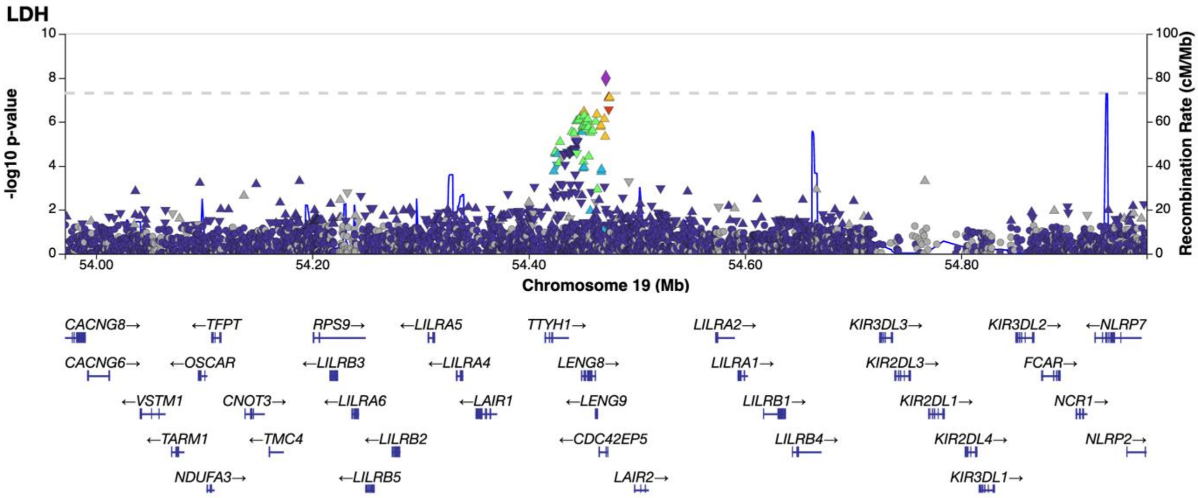


**Fig. S1.36** Regional association plot of novel LDH association on chromosome 19 limited to the area of 53971384-54971384. The association is based on the results of a meta-analysis. According to our research, the gene explaining the association of the locus is possibly *LENG8 (leukocyte receptor cluster member 8).*

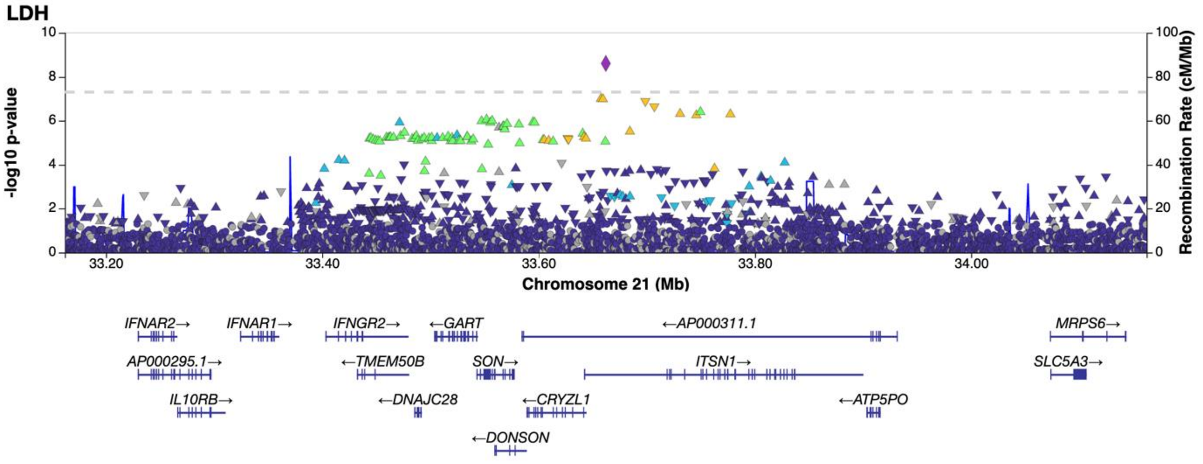


**Fig. S1.37** Regional association plot of novel LDH association on chromosome 21 limited to the area of 33162166-3416166. The association is based on the results of a meta-analysis. According to our research, the gene explaining the association of the locus is possibly *SLC5A3 (solute carrier family 5 member 3).*

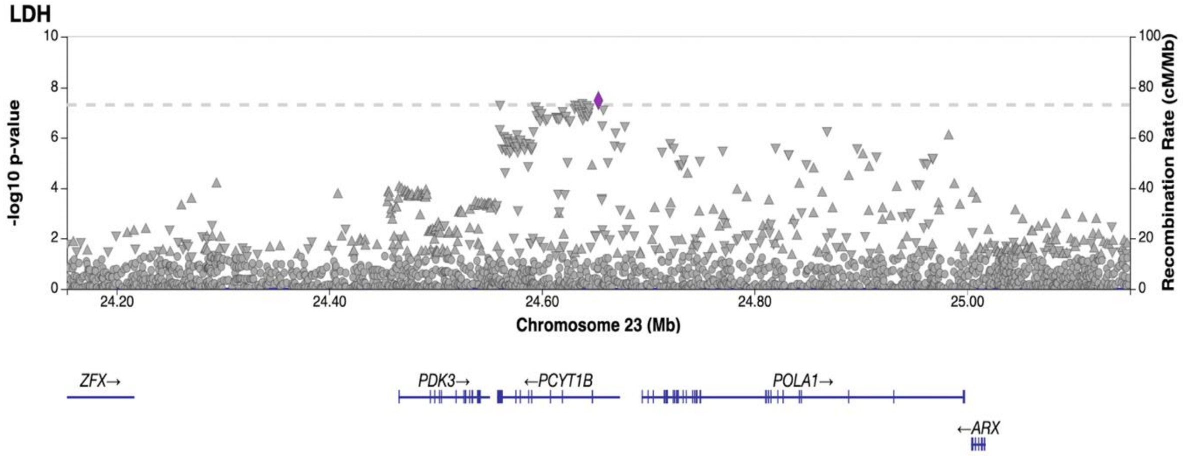


**Fig. S1.38** Regional association plot of novel LDH association on chromosome X limited to the area of 24153216-34153216. The association is based on the results of a meta-analysis. According to our research, the gene explaining the association of the locus is possibly *PCYT1B (phosphate cytidylyltransferase 1B).*

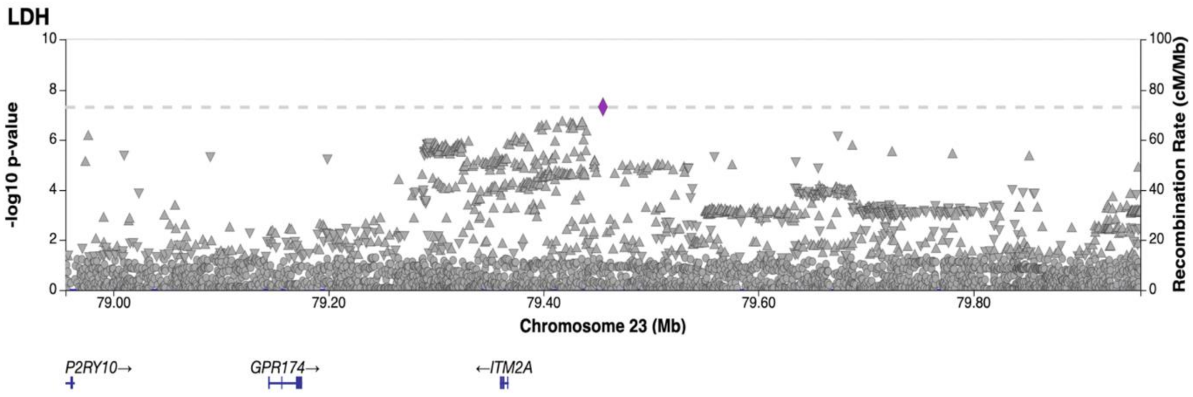


**Fig. S1.39** Regional association plot of novel LDH association on chromosome X limited to the area of 78955466-79955466. The association is based on the results of a meta-analysis. According to our research, the gene explaining the association of the locus is possibly *ITM2A (integral membrane protein 2A).*

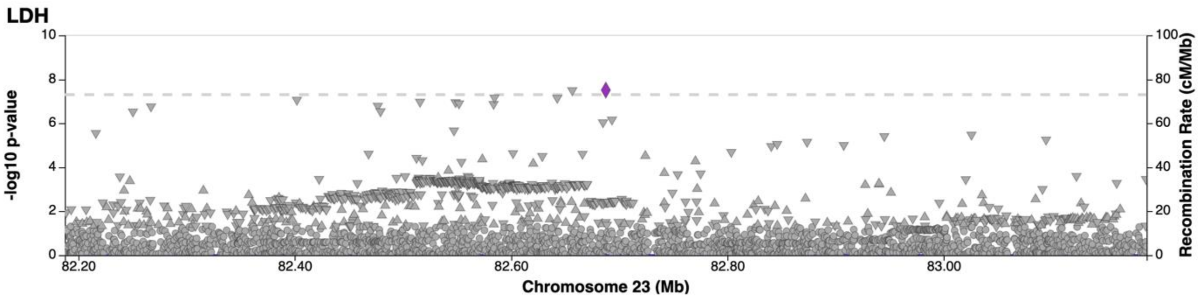


**Fig. S1.40** Regional association plot of novel LDH association on chromosome X limited to the area of 82187578-83187578. The association is based on the results of a meta-analysis. There was not any protein coding gene in this locus, so it was classified as Empty.

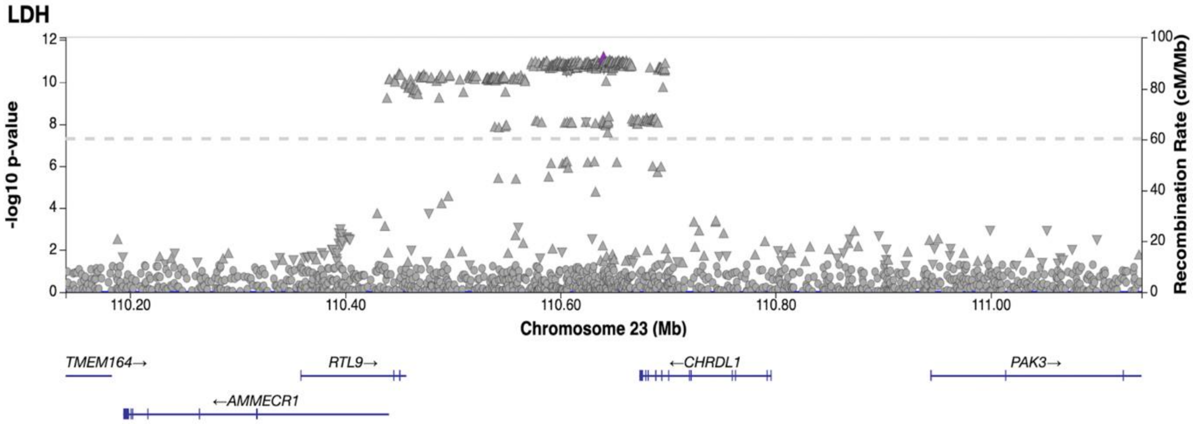


**Fig. S1.41** Regional association plot of novel LDH association on chromosome X limited to the area of 110140115-111140115. The association is based on the results of a meta-analysis. According to our research, the gene explaining the association of the locus is possibly *CHRDL1 (chordin like 1).*

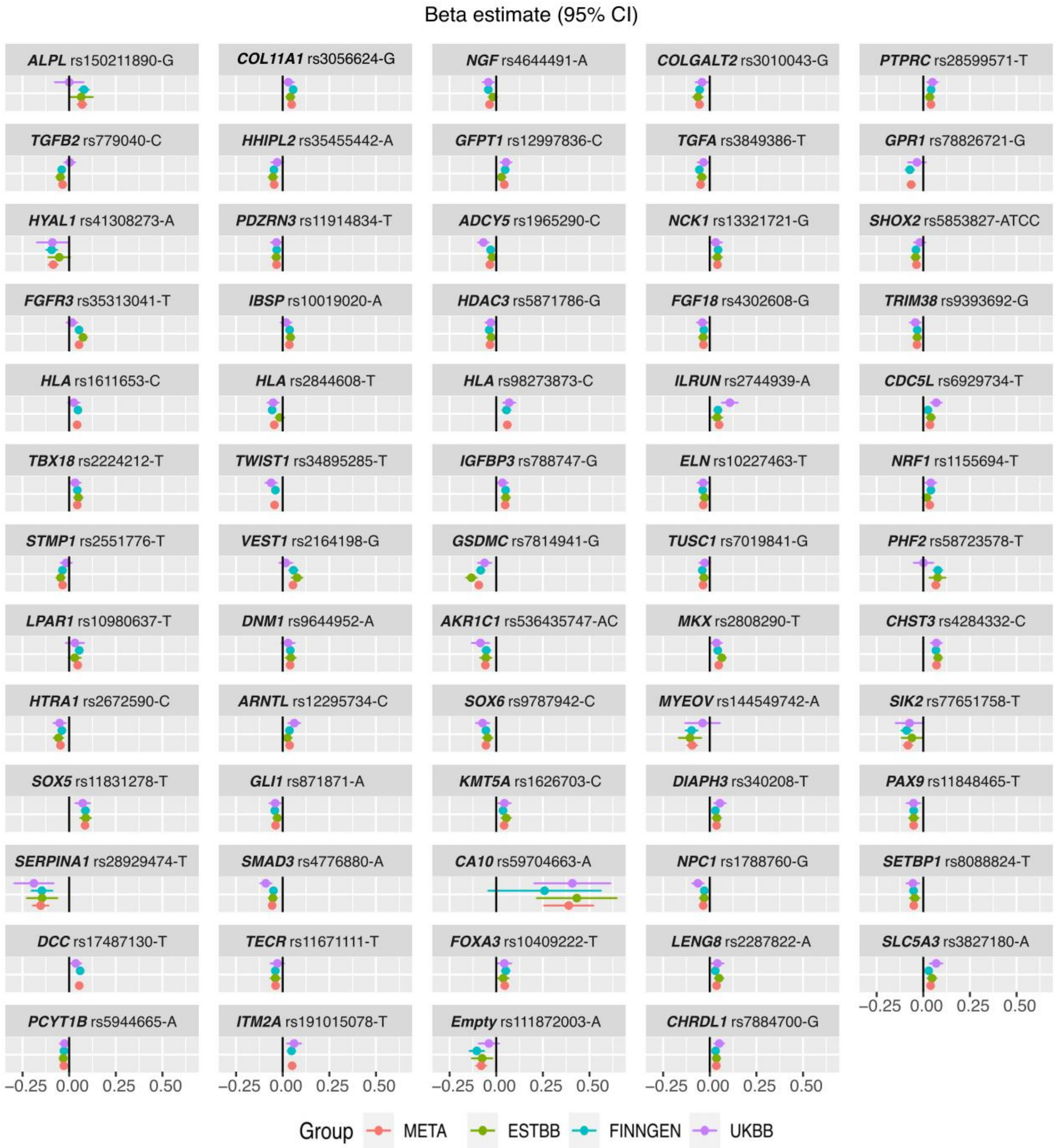


**Fig. S2.** Effect estimates of lead variants and their 95% confidence intervals in the forest plot. Variants are identified by candidate gene, rsid and effect allele. The diagram shows the effect estimates for each variant both in the meta-analysis and in each dataset separately (FinnGen, ESTBB, UKBB).

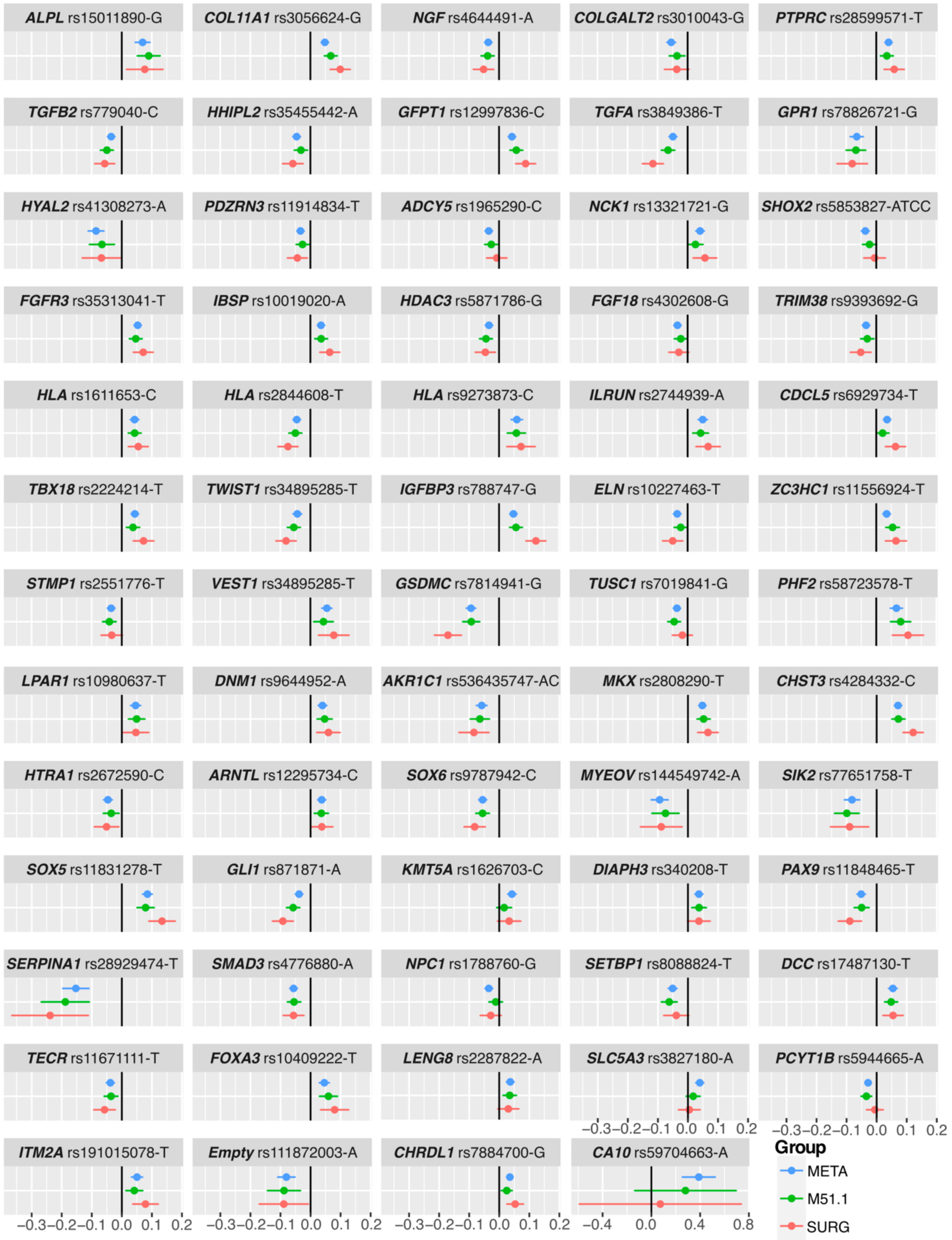


**Fig. S3.** Effect estimates of lead variants observed in meta-analysis and sensitivity analyses. Variants are identified by candidate gene, rsid and effect allele. The diagram shows the effect estimates for each variant both in the meta-analysis and sensitivity analyses (M51.1, SURG). M51.1 contains only cases that had only M51.1 diagnoses. M51.1 GWAS contains 18857 cases and 270 964 controls. Also, GWAS was performed for patients that had had an LDH related operation. This phenotype contains all patients that had surgical codes of ABC07, ABC16, ABC26. SURG GWAS contains 7347 cases and 270 964 controls. We did not observe any statistically significant effect differences between meta-analysis and M51.1. Between meta-analysis and SURG we observed statistically significant effect differences for *COL11A1*, *GFPT1*, *TGFA*, *TWIST1*, *IGFBP3*, *GSDMC*, *CHST3*, *SOX5* and *GLI1* variants. Results of the effect differences can also be seen from Table S3.

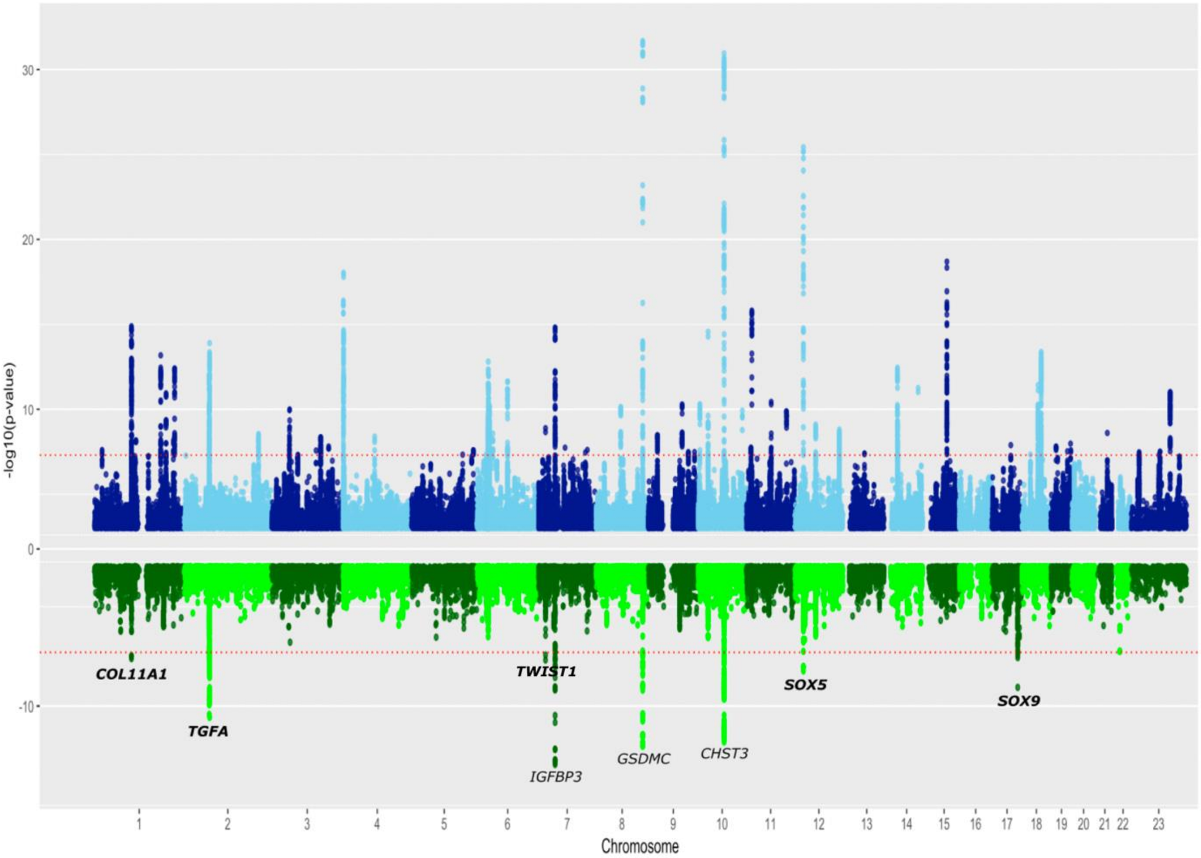


**Fig. S4** Above, Manhattan plot of the variants we detected to be associated with LDH in the meta-analysis and below variants we detected to be associated with LDH related surgical operation. Meta-analysis contains 80724 LDH cases and 748 975 controls, consisting of data from FinnGen, the Estonian Biobank, and the UK Biobank. SURG GWAS, where LDH patients who had undergone an LDH-related operation (ABC07, ABC16, ABC26) formed a case group of 7347 and controls of 270 964. Results for the SURG GWAS can be seen in Table S4.

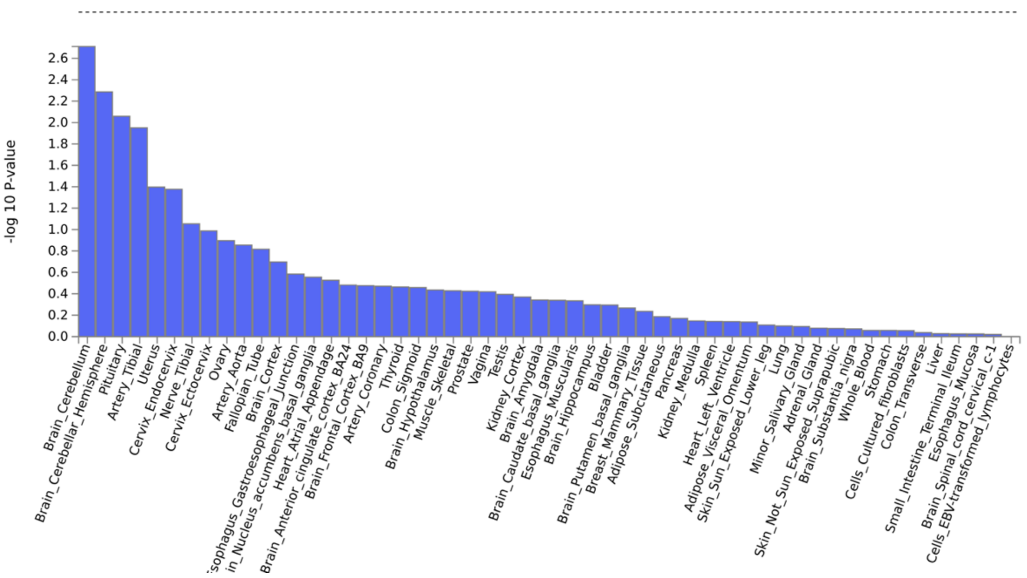


**Fig. S5.** Results of MAGMA tissue expression analysis[1], no genome-wide significant results were observed in tissue expression analysis. All bars fell below the dashed line, which serves as a marker for genome-wide significant results. y-axis, -log 10 P-value; x-axis, tissues (GTEx Output-General tissues). Analysis was done by using FUMA[2].

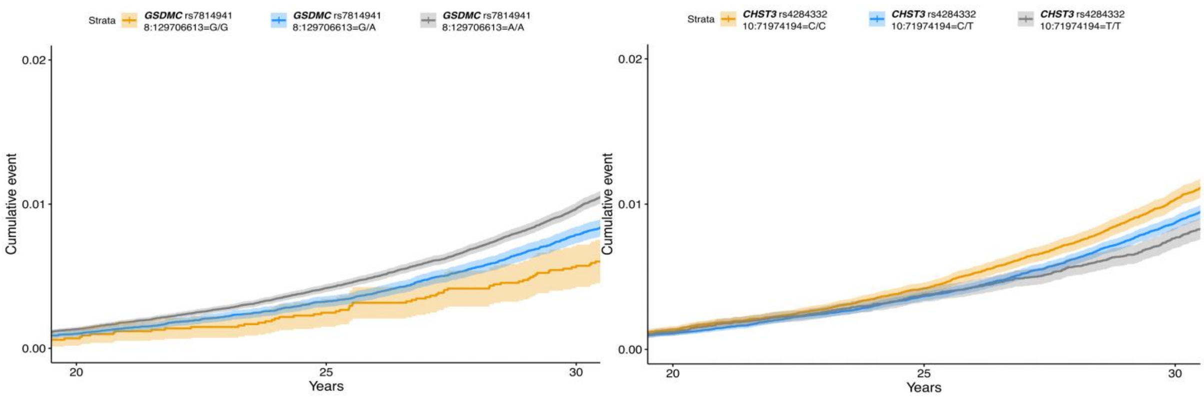


**Fig. S6** Closer look for *GSDMC (8:129706613:G:A, rs7814941)* and *CHST3 (10:719741194:C:T, rs4284332)* variants. The same variants are used, and the curves correspond to the curves in Fig. 3, but in this graph, the development can only be seen at the age of 20–30 years. For these variants, a statistically significant difference between the genotypes was observed even before the age of 30. *GSDMC* 8:12970613-A/A differed from the variant's other genotypes at the age of 26 (p=0.0005). *CHST3* 10:71974194-C/C homozygotes became significantly different at the age of 25 ($p=2.22e^{-5}$).

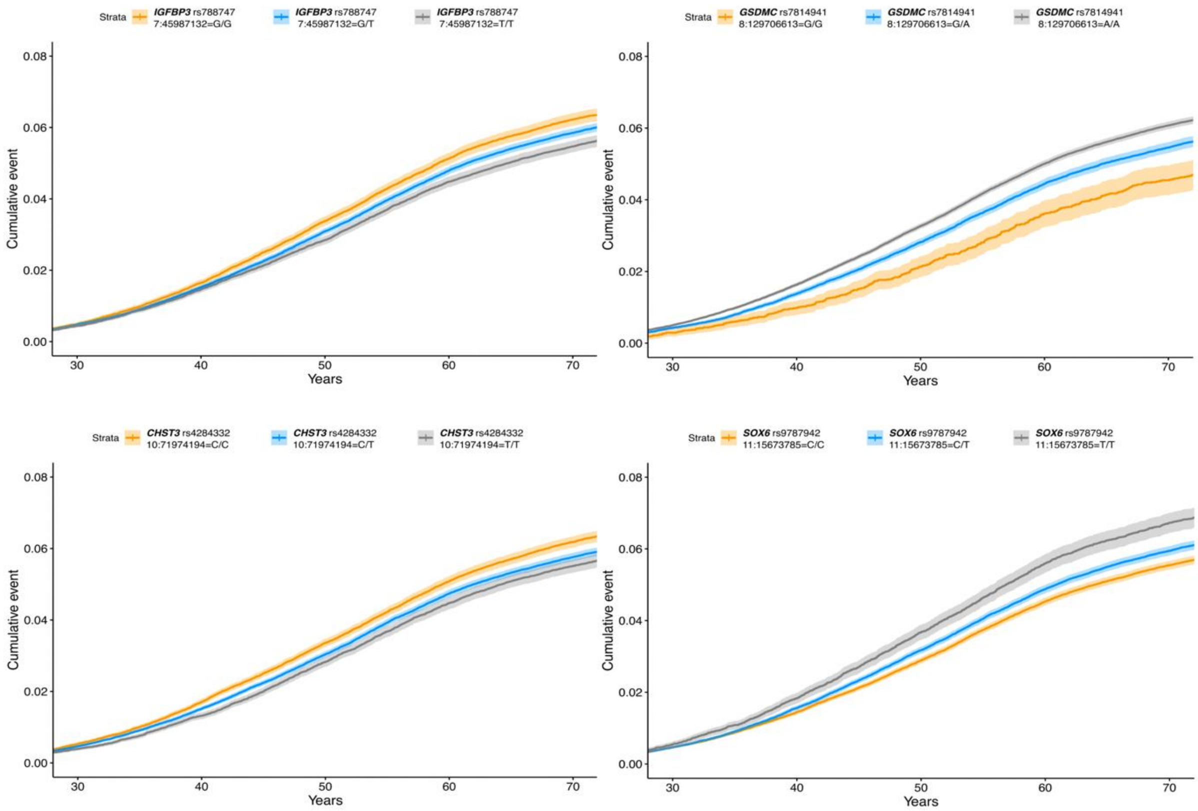


**Fig. S7** Kaplan-Meier plots for *IGFBP3 (7:45987132:G:T), CHST3 (10:719741194:C:T), GSDMC (8:129706613:G:A), SOX6 (11:15673785:C:T).* These plots were done as a sensitivity analysis based only M51.1 cases. Age can be seen on the x-axis of the graphs, and cumulative disease severity on the y-axis. The orange line of the graphs depicts homozygotes for the effect allele, gray homozygotes for the other allele, and blue correspondingly heterozygotes who have one of each allele.

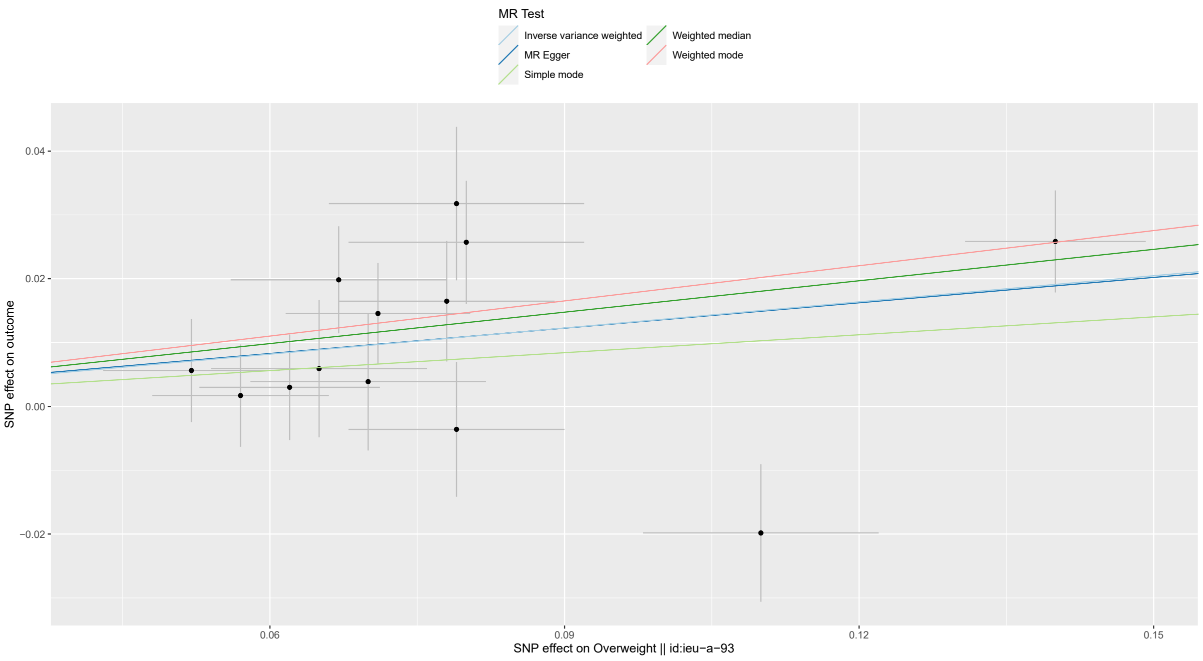


**Fig. S8.1** Scatterplot for overweight that could possibly be causal for lumbar disc herniation. In Mendelian randomization analysis, performed with TwoSampleMR-database. We focused on the results of the Inverse variance Weighted (IVW) model, with the significance limit of ($P < 0.05$).

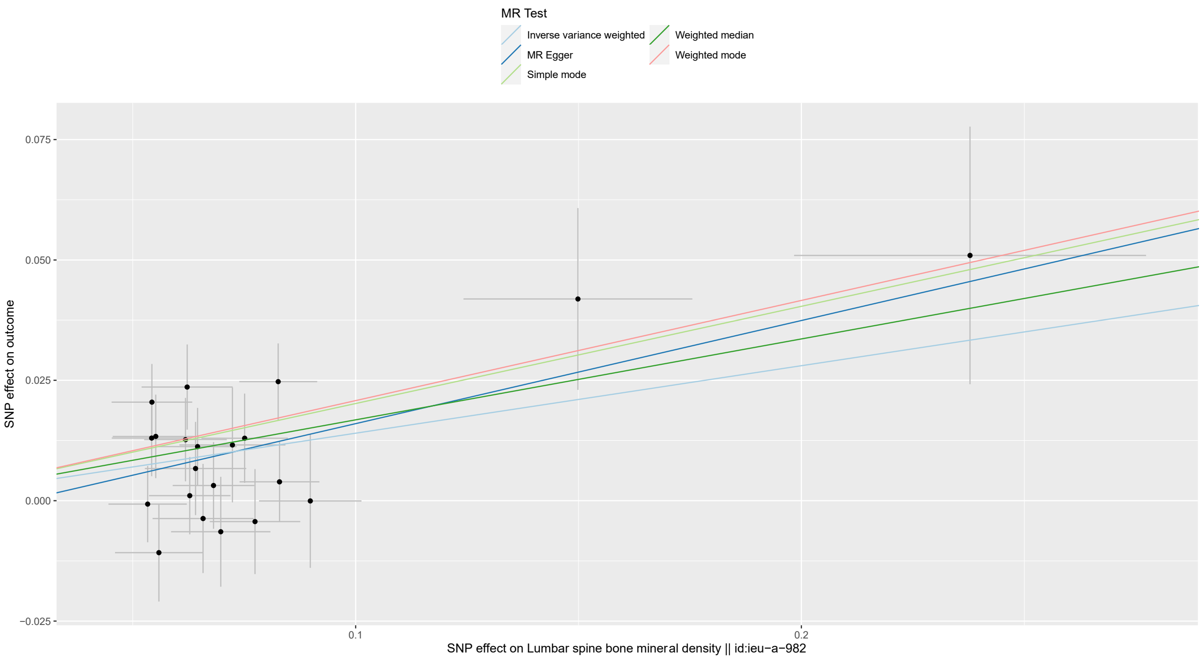


**Fig. S8.2** Scatterplot for lumbar spine bone mineral density that could possibly be causal for lumbar disc herniation. In Mendelian randomization analysis, performed with TwoSampleMR-database. We focused on the results of the Inverse variance Weighted (IVW) model, with the significance limit of ($P < 0.05$).

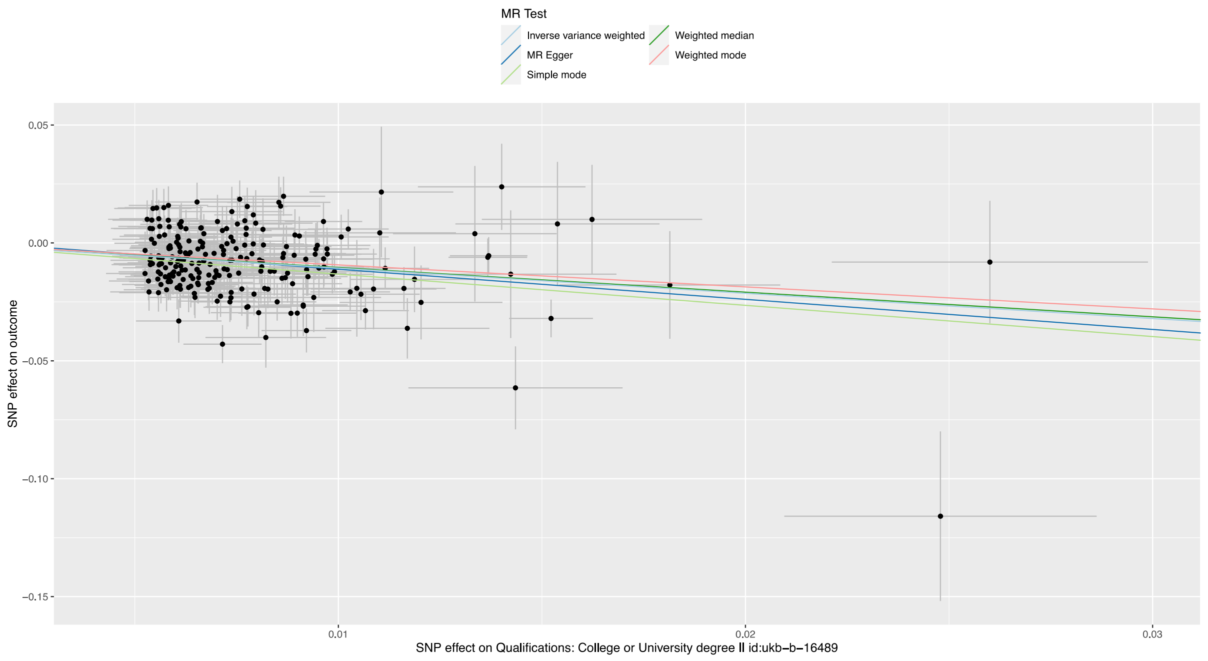


**Fig. S8.3** Scatterplot for Higher level of education that could possibly be causal for lumbar disc herniation. In Mendelian randomization analysis, performed with TwoSampleMR-database. We focused on the results of the Inverse variance Weighted (IVW) model, with the significance limit of ($P < 0.05$). Original endpoint name is Qualifications: College or University degree || id:ukb-b-16489.

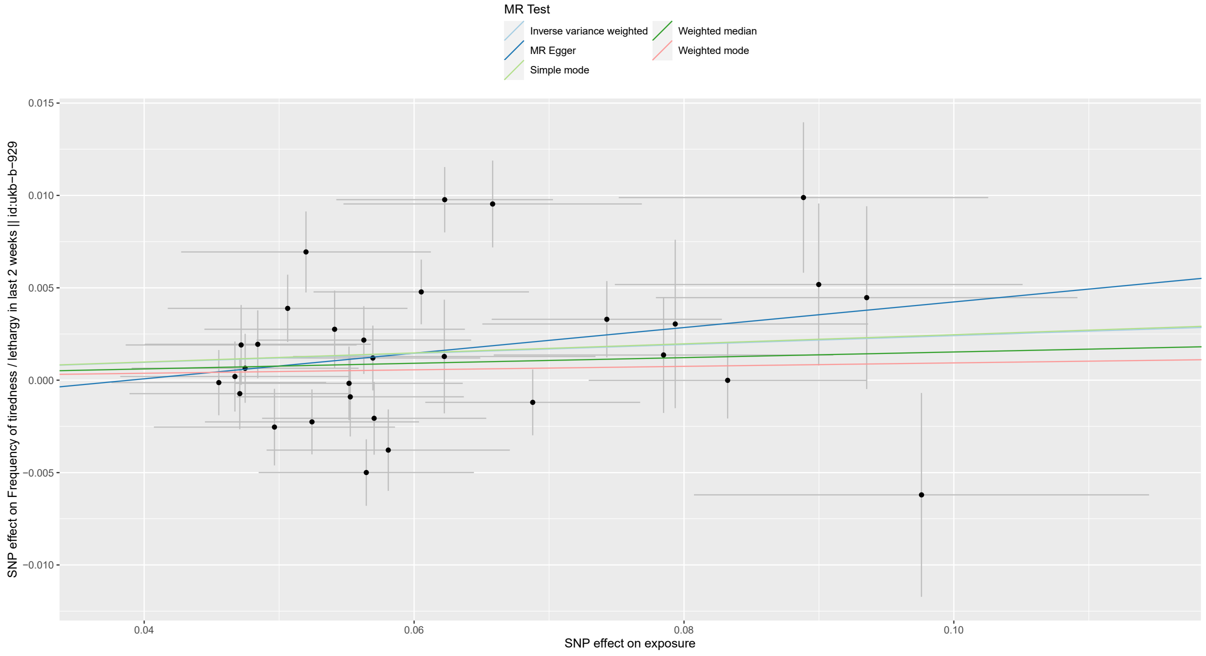


**Fig. S9.1** Scatterplot for frequency of tiredness in last 2 weeks-endpoint for which lumbar disc herniation is potentially causal. In Mendelian randomization analysis, performed with TwoSampleMR-database. We focused on the results of the Inverse variance Weighted (IVW) model, with the significance limit of ($P < 0.05$).

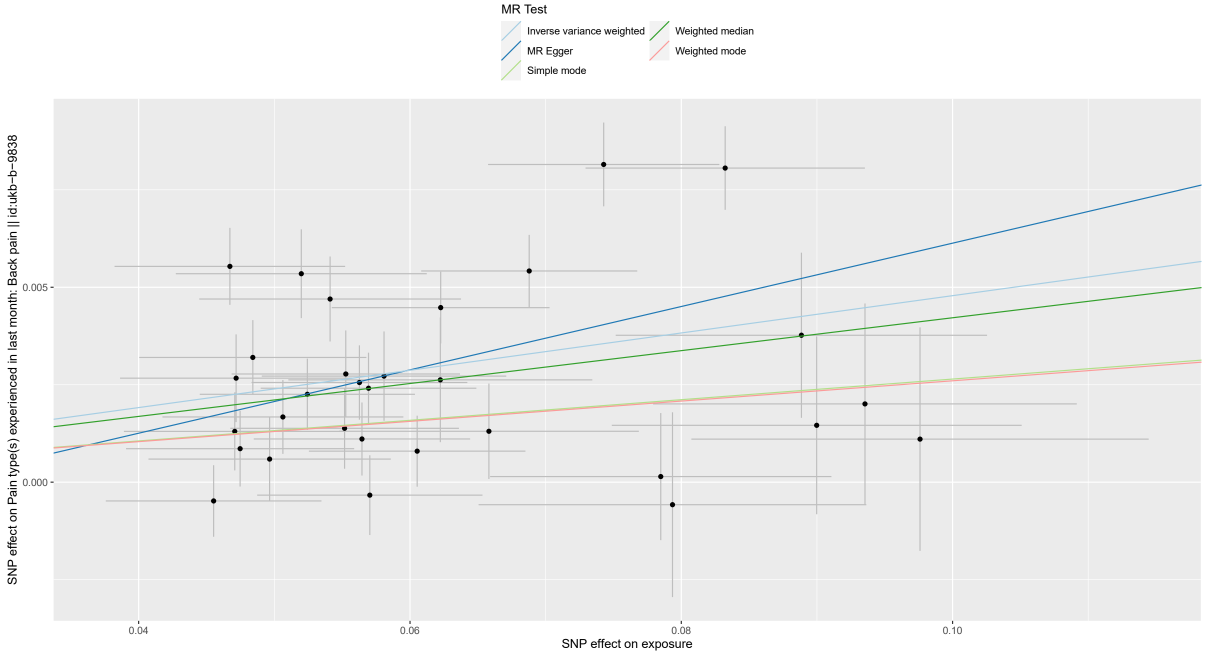


**Fig. S9.2** Scatterplot for back pain for which lumbar disc herniation is potentially causal. In Mendelian randomization analysis, performed with TwoSampleMR-database. We focused on the results of the Inverse variance Weighted (IVW) model, with the significance limit of ($P < 0.05$).

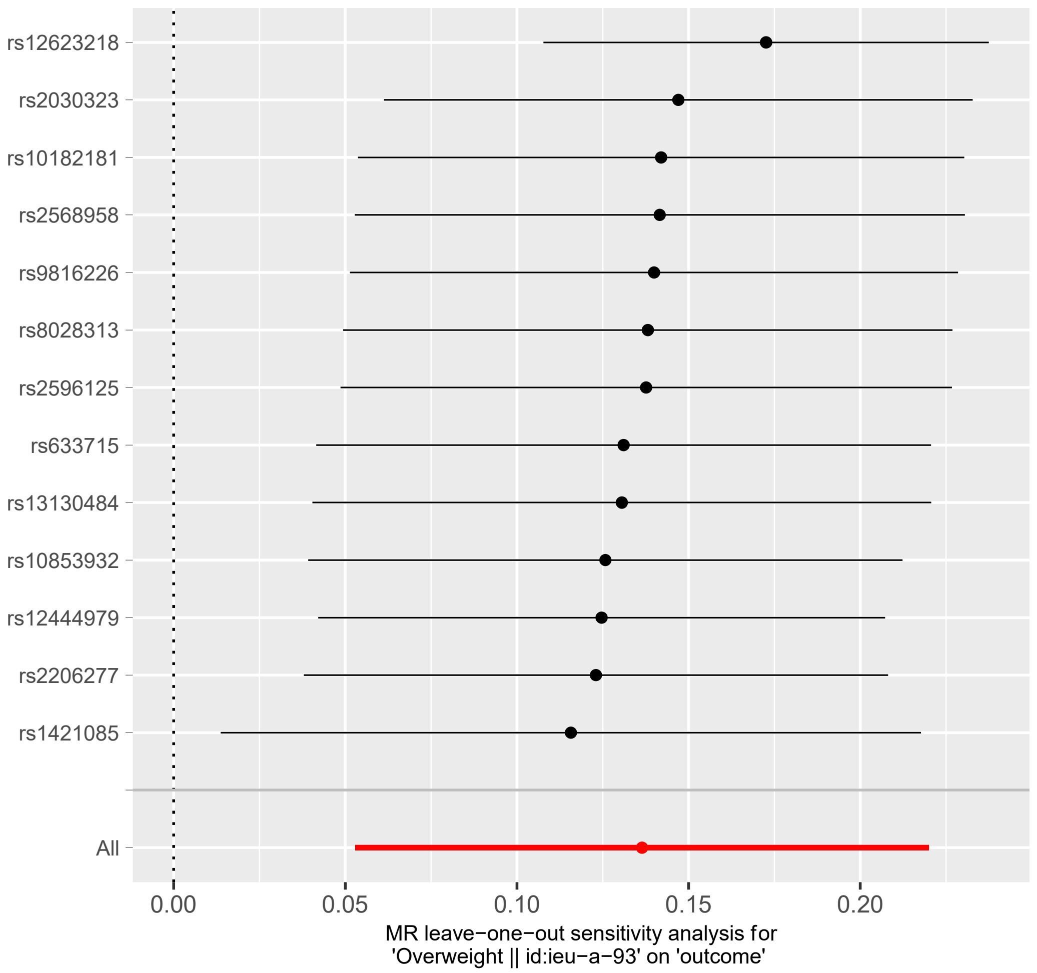


**Fig. S10.1** Leave-one-out, overweight (id:ieu-a-93) as an exposure and lumbar disc herniation as an outcome

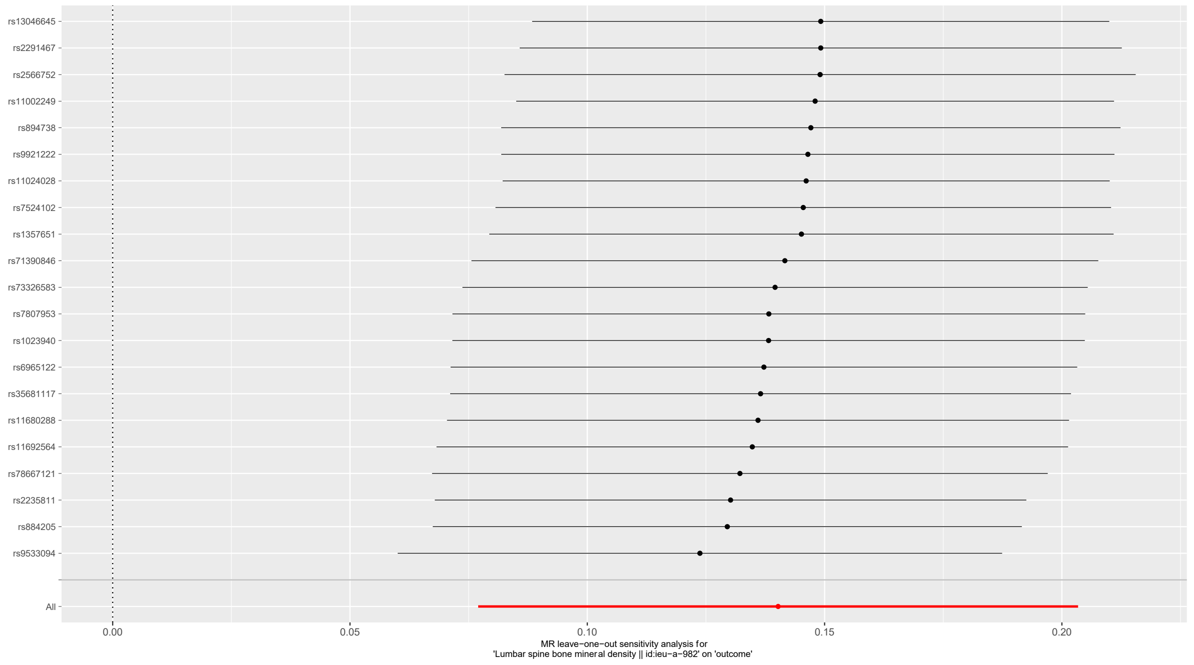


**Fig. S10.2** Leave-one-out, lumbar spine bone mineral density (id:ieu-a-982) as an exposure and lumbar disc herniation as an outcome

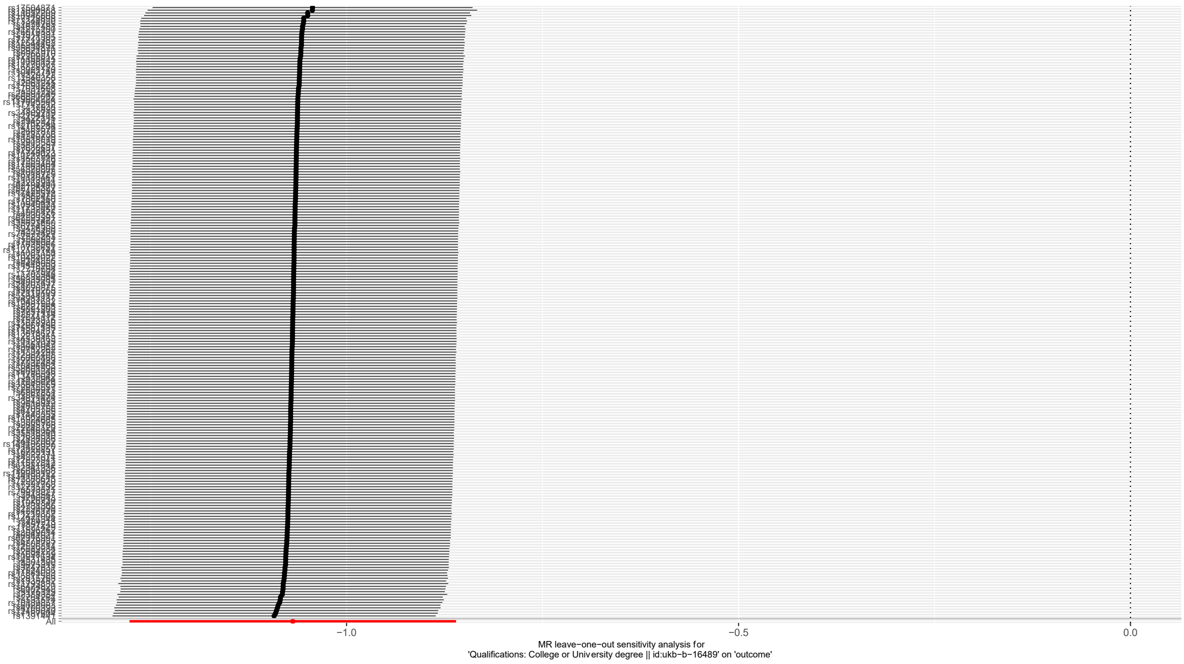


**Fig. S10.3** Leave-one-out, higher level of education as an exposure and lumbar disc herniation as an outcome. Original endpoint name is Qualifications: College or University degree || id:ukb-b16489.

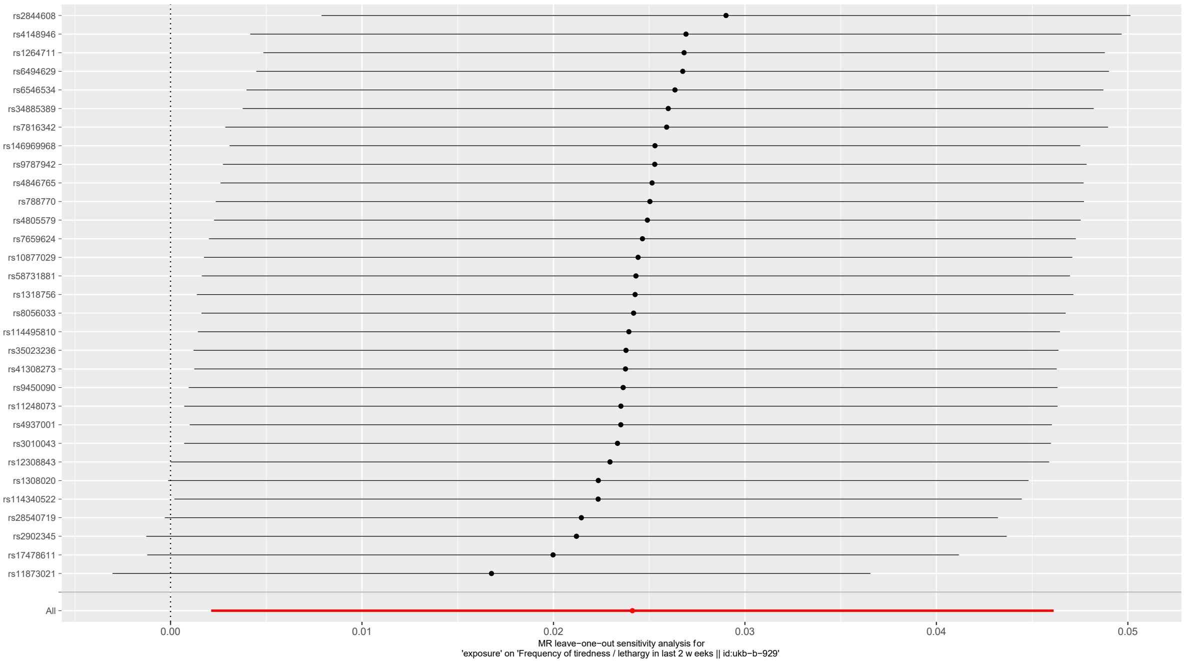


**Fig. S11.1** Leave-one-out, lumbar disc herniation as an exposure and frequency of tiredness in last 2 weeks (id:ukb-b-929) as an outcome.

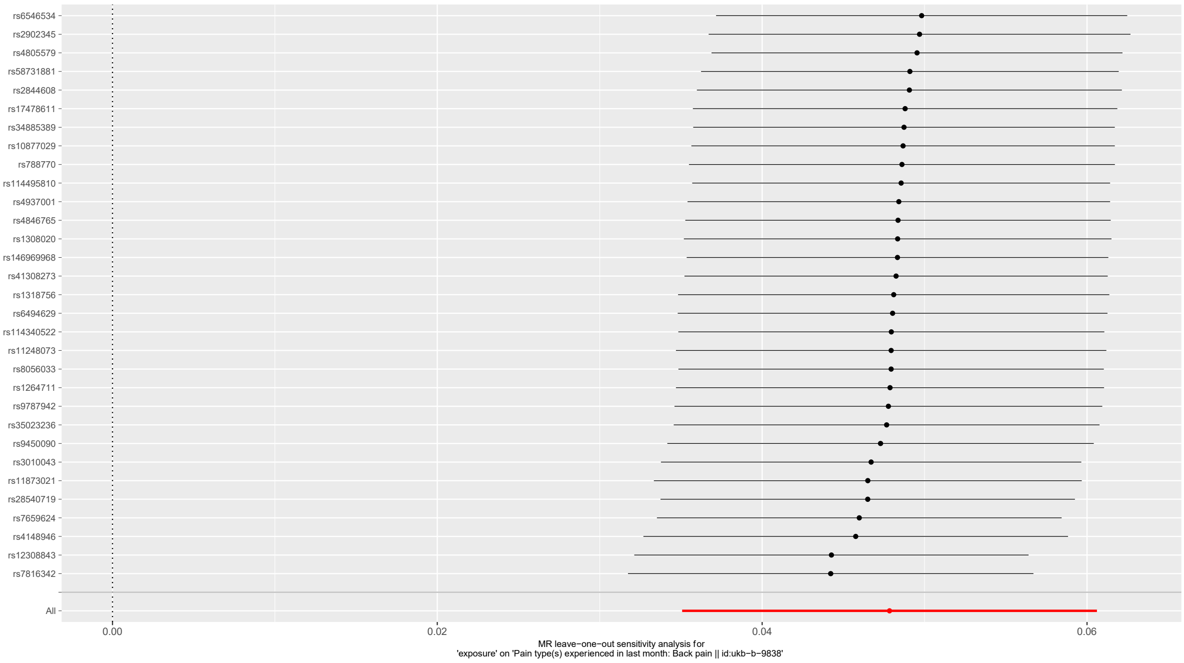


**Fig. S11.2** Leave-one-out, lumbar disc herniation as an exposure and back pain (id:ukb-b-9838) as an outcome.

**Table S1** General information about the study populations. The International Classification of Diseases codes that were used to characterize phenotypes. Below, the number of cases and controls in the study.

| Study | Revision / Codes | Codes |
|---|---|---|
| **Meta-analysis** | | |
| EstBB | ICD-10 | M51 (M510, M511, M512, M513, M514, M518, M519) |
| UKBB | ICD-10 | M51 (M510, M511, M512, M513, M514, M518, M519) |
| FinnGen | ICD-10 | M51 (M510, M511, M512, M513, M514, M518, M519) |
| | ICD-9 | 7221A, 7221C, 7223A, 7225A, 7225B, 7226X, 7228C, 7229X |
| | ICD-8 | 72510, 72519, 72520, 72551, 72559, 72588, 72599 |
| **Sensitivity analysis** | | |
| **LDH cases with radiculopathy** | | |
| Step 1) Characterization of patients with radiculopathy | | |
| | ICD-10 | M51.1 |
| | ICD-9 | 7221A |
| | ICD-8 | 72510 |
| Step 2) Patients with other LDH diagnosis codes were excluded from the analysis. | | |
| **Surgical GWAS** | | |
| Step 1) Characterization of LDH patients in FinnGen | | |
| | ICD-10 | M51 (M510, M511, M512, M513, M514, M518, M519) |
| | ICD-9 | 7221A, 7225A, 7225B, 7225C |
| | ICD-8 | 72510, 72519 |
| Step 2) Patients that have been surgically operated were picked from the characterized LDH patients group | | |
| | NOMESCO, version 1.15 | ABC07, ABC16, ABC26 |
| Step 3) LDH cases that have not been surgically operated were excluded from the analysis. Also, surgically operated patients without LDH diagnosis were excluded | | |

| Study population | Cases | Controls | Sample prevalence | Total |
|---|---|---|---|---|
| FinnGen | 37 636 | 270 964 | 12.2 % | 308 600 |
| Estonian Biobank | 34 035 | 66 533 | 33.8 % | 100 568 |
| UK Biobank | 9053 | 411 478 | 2.2 % | 420 531 |
| **Meta-analysis** | **80 724** | **748 975** | **9.7 %** | **829 699** |
| FinnGen subgroup analysis | | | | |
| LDH cases with radiculopathy | 18 857 | 270 964 | 6.5 % | 289 821 |
| Surgical GWAS | 7347 | 270 964 | 2.6 % | 278 311 |

**Table S2.** The list of lead variants at the 64 genome-wide significant (p<5x10$^{-8}$) loci that were associated with LDH in the meta-analysis, that contained total of 80 724 LDH cases and 748 975 controls from FinnGen, the Estonian Biobank, and the UK Biobank. We also performed conditional analyzes for the loci to identify possible secondary signals. The analyzes were performed with the GCTA software package[3], and the lead variants detected from the loci were used as a covariate. Those observed secondary signals The detected secondary signals are in the table under the lead variant used as a covariate, and the results of the secondary signal are also conditional on the lead variant used as a covariate.

| Locus | Candidate gene | CHR: POS | rsid | EA | OA | OR | OR 95CI | p-value | EAF | Fin. | Ref. |
|---|---|---|---|---|---|---|---|---|---|---|---|
| **1p36.12** | ***ALPL*** | **1:21559185** | **rs150211890** | **G** | **T** | **1.07** | **1.05-1.10** | **2.5e-08** | **0.05** | **2.6** | **Novel** |
| 1p21.1 | *COL11A1* | 1:102882172 | rs3056624 | G | GTATT | 1.05 | 1.04-1.06 | 1.29e-15 | 0.51 | 1.1 | [4] |
| **1p13.2** | ***NGF*** | **1:115310363** | **rs4644491** | **A** | **G** | **0.96** | **0.95-0.98** | **6.78e-09** | **0.63** | **0.96** | **Novel** |
| 1q25.3 | *COLGALT2* | 1:183973041 | rs3010043 | G | A | 0.95 | 0.93-0.96 | 6.64e-14 | 0.78 | 1.0 | [4] |
| 1q32.1 | *PTPRC* | 1:198768851 | rs28599571 | T | G | 1.04 | 1.03-1.05 | 1.15e-11 | 0.55 | 1.1 | [4] |
| **1q41** | ***TGFB2*** | **1:218924545** | **rs779040** | **C** | **G** | **0.97** | **0.95-0.98** | **7.13e-09** | **0.55** | **1.0** | **Novel** |
| **1q41** | ***HHIPL2*** | **1:222541797** | **rs35455442** | **A** | **C** | **0.96** | **0.94-0.97** | **3.96e-13** | **0.33** | **1.3** | **Novel** |
| 2p13.3 | *GFPT1* | 2:69304791 | rs12997836 | C | T | 1.04 | 1.03-1.06 | 9.44e-13 | 0.43 | 1.1 | [4] |
| 2p13.3 | *TGFA* | 2:70496764 | rs3849386 | T | C | 0.95 | 0.94-0.96 | 1.27e-14 | 0.35 | 1.1 | [4] |
| “ | *TGFA** | 2:69569876 | rs6546534 | T | G | 1.05 | 1.04-1.07 | 4.47e-12 | 0.30 | 1.29 | |
| **2q33.3** | ***GPR1*** | **2:206137590** | **rs78826721** | **G** | **A** | **0.94** | **0.92-0.96** | **2.85e-09** | **0.11** | **1.22** | **Novel** |
| **3p21.31** | ***HYAL2*** | **3:50380254** | **rs41308273** | **A** | **T** | **0.92** | **0.89-0.94** | **1.02e-10** | **0.05** | **2.4** | **Novel** |
| **3p13** | ***PDZRN3*** | **3:73200973** | **rs11914834** | **T** | **C** | **0.97** | **0.96-0.98** | **4.85e-08** | **0.44** | **1.1** | **Novel** |
| **3q21.1** | ***ADCY5*** | **3:123568959** | **rs1965290** | **C** | **T** | **0.97** | **0.95-0.98** | **2.67e-08** | **0.55** | **1.1** | **Novel** |
| **3q22.3** | ***NCK1*** | **3:136490708** | **rs13321721** | **G** | **A** | **1.04** | **1.03-1.06** | **4.49e-09** | **0.24** | **1.1** | **Novel** |
| **3q25.32** | ***SHOX2*** | **3:158478549** | **rs5853827** | **ATCC** | **A** | **0.96** | **0.95-0.98** | **1.55e-08** | **0.33** | **0.91** | **Novel** |
| 4p16.3 | *FGFR3* | 4:1694376 | rs35313041 | T | C | 1.05 | 1.04-1.07 | 9.14e-19 | 0.52 | 0.98 | [4] |
| “ | *FGFR3** | 4:1185425 | rs7659624 | T | C | 1.05 | 1.04-1.07 | 2.99e-15 | 0.69 | 0.97 | |
| **4q22.1** | ***IBSP*** | **4:87779677** | **rs10019020** | **A** | **G** | **1.04** | **1.02-1.05** | **4.07e-09** | **0.49** | **0.85** | **Novel** |
| **5q31.3** | ***HDAC3*** | **5:141735121** | **rs5871786** | **G** | **GT** | **0.97** | **0.96-0.98** | **4.71e-08** | **0.44** | **1.0** | **Novel** |
| **5q35.1** | ***FGF18*** | **5:171413500** | **rs4302608** | **G** | **A** | **0.97** | **0.95-0.98** | **2.61e-08** | **0.55** | **0.98** | **Novel** |
| **6p22.2** | ***TRIM38*** | **6:26276422** | **rs9393692** | **G** | **A** | **0.97** | **0.95-0.98** | **4.46e-08** | **0.58** | **1.2** | **Novel** |
| **6p22.1** | ***HLA*** | **6:29873925** | **rs1611653** | **C** | **G** | **1.04** | **1.03-1.06** | **1.98e-09** | **0.58** | **0.93** | **Novel** |
| **6p21.33** | ***HLA*** | **6:31279637** | **rs2844608** | **T** | **C** | **0.96** | **0.94-0.97** | **1.58e-13** | **0.39** | **1.1** | **Novel** |
| **“** | ***HLA**** | **6:32236907** | **rs3134924** | **T** | **G** | **1.06** | **1.04.1.08** | **4.51e-08** | **0.15** | **0.89** | |
| 6p21.32 | *HLA* | 6:32661873 | rs9273873 | C | T | 1.06 | 1.04-1.08 | 4.42e-10 | 0.21 | NA | [5] |
| 6p21.31 | *ILRUN* | 6:34580429 | rs2744939 | A | T | 1.05 | 1.04-1.07 | 6.36e-11 | 0.17 | 1.4 | [4] |
| 6p21.1 | *CDC5L* | 6:44478351 | rs6929734 | T | G | 1.04 | 1.02-1.05 | 2.72e-09 | 0.54 | 0.98 | [4] |
| **6q14.3** | ***TBX18*** | **6:84938145** | **rs2224214** | **T** | **C** | **1.04** | **1.03-1.06** | **2.33e-12** | **0.37** | **1.0** | **Novel** |
| 7p21.1 | *TWIST1* | 7:19442778 | rs34895285 | T | TA | 0.96 | 0.94-0.97 | 1.29e-09 | 0.52 | 0.99 | [4] |
| 7p12.3 | *IGFBP3* | 7:45987132 | rs788747 | G | T | 1.05 | 1.04-1.06 | 1.53e-15 | 0.50 | 0.96 | [4] |
| **7q11.23** | ***ELN*** | **7:73714641** | **rs10227463** | **T** | **C** | **0.97** | **0.95-0.98** | **1.82e-08** | **0.41** | **1.2** | **Novel** |
| **7q32.2** | ***ZC3HC1*** | **7:130023656** | **rs11556924** | **T** | **C** | **1.04** | **1.02-1.05** | **3.48e-08** | **0.36** | **0.84** | **Novel** |
| **7q33** | ***STMP1*** | **7:135418700** | **rs2551776** | **T** | **C** | **0.97** | **0.95-0.98** | **2.48e-08** | **0.64** | **1.1** | **Novel** |
| 8q13.2 | *VEST1* | 8:68665207 | rs2164198 | G | A | 1.06 | 1.04-1.07 | 7.35e-11 | 0.18 | 0.45 | [4] |
| “ | *VEST1** | 8:69499061 | rs1968555 | C | G | 1.03 | 1.02-1.05 | 2.18e-08 | 0.40 | 1.32 | |
| 8q24.21 | *GSDMC* | 8:129706613 | rs7814941 | G | A | 0.91 | 0.90-0.93 | 2.1e-32 | 0.19 | 0.94 | [6] |
| **9p21.3** | ***TUSC1*** | **9:25398495** | **rs7019841** | **G** | **A** | **0.96** | **0.95-0.98** | **3.34e-09** | **0.54** | **0.96** | **Novel** |
| 9q22.32 | *PHF2* | 9:93911476 | rs58723578 | T | C | 1.07 | 1.05-1.09 | 5.18e-11 | 0.10 | 1.2 | [4] |
| **9q31.3** | ***LPAR1*** | **9:110930238** | **rs10980637** | **T** | **C** | **1.05** | **1.03-1.06** | **3.28e-08** | **0.13** | **1.9** | **Novel** |
| **9q34.11** | ***DNM1*** | **9:128236873** | **rs9644952** | **A** | **C** | **104** | **1.03-1.05** | **3.3e-08** | **0.22** | **1.1** | **Novel** |
| **10p15.1** | ***AKR1C1*** | **10:4989436** | **rs536435747** | **AC** | **A** | **0.94** | **0.93-0.96** | **5.02e-11** | **0.13** | **1.2** | **Novel** |
| 10p21.1 | *MKX* | 10:27611953 | rs2808290 | T | C | 1.05 | 1.04-1.06 | 2.75e-15 | 0.46 | 0.78 | [4] |
| 10q22.1 | *CHST3* | 10:71974194 | rs4284332 | C | T | 1.07 | 1.06-1.09 | 1.13e-31 | 0.58 | 0.89 | [7] |
| **10q26.13** | ***HTRA1*** | **10:122475088** | **rs2672590** | **C** | **A** | **0.95** | **094-0.97** | **1.2e-10** | **0.24** | **0.89** | **Novel** |
| **“** | ***HTRA1**** | **10:122301287** | **rs10788274** | **A** | **G** | **0.96** | **0.95-0.97** | **4.52e-11** | **0.55** | **1.18** | |
| 11p15.3 | *ARNTL* | 11:13270734 | rs12295734 | C | G | 1.04 | 1.02-1.05 | 1.81e-08 | 0.71 | 1.0 | [4] |
| 11p15.2 | *SOX6* | 11:15673785 | rs9787942 | C | T | 0.95 | 0.93-0.96 | 1.55e-16 | 0.75 | 0.85 | [4] |
| **11q13.3** | ***MYEOV*** | **11:69208032** | **rs144549742** | **A** | **T** | **0.91** | **0.88-0.94** | **3.66e-11** | **0.04** | **2.8** | **Novel** |
| **11q23.1** | ***SIK2*** | **11:111459420** | **rs77651758** | **T** | **C** | **0.92** | **0.90-0.95** | **1.32e-10** | **0.05** | **2.1** | **Novel** |
| 12p12.1 | *SOX5* | 12:23807795 | rs11831278 | T | C | 1.09 | 1.07-1.10 | 3.73e-26 | 0.16 | 1.0 | [4] |
| **12q14.1** | ***GLI1*** | **12:57825898** | **rs871871** | **A** | **G** | **0.96** | **0.95-0.97** | **7.98e-10** | **0.35** | **1.3** | **Novel** |
| **12q24.31** | ***KMT5A*** | **12:123226288** | **rs1626703** | **C** | **A** | **1.04** | **1.03-1.06** | **1.55e-09** | **0.75** | **1.0** | **Novel** |
| **13q21.2** | ***DIAPH3*** | **13:59904471** | **rs340208** | **T** | **A** | **1.04** | **1.02-1.05** | **3.98e-08** | **0.70** | **1.1** | **Novel** |
| 14q13.3 | *PAX9* | 14:37002513 | rs11848465 | T | C | 0.95 | 0.94-0.96 | 3.58e-13 | 0.22 | 1.2 | [4] |
| 14q32.13 | *SERPINA1* | 14:94378610 | rs28929474 | T | C | 0.86 | 0.81-0.90 | 5.59e-12 | 0.02 | 1.0 | [4] |
| 15q22.33 | *SMAD3* | 15:67072653 | rs4776880 | A | G | 0.95 | 0.93-0.96 | 2.02e-19 | 0.35 | 1.1 | [4] |
| **17q22** | ***CA10*** | **17:52164544** | **rs59704663** | **A** | **G** | **1.48** | **1.34-1.61** | **1.29e-08** | **0.003** | **0.12** | **Novel** |
| **18q11.2** | ***NPC1*** | **18:23557478** | **rs1788760** | **G** | **A** | **0.97** | **0.95-0.98** | **4.62e-08** | **0.67** | **1.0** | **Novel** |
| **18q12.3** | ***SETBP1*** | **18:44571296** | **rs8088824** | **T** | **C** | **0.95** | **0.94-0.96** | **3.76e-12** | **0.78** | **1.1** | **Novel** |
| **18q21.2** | ***DCC*** | **18:53189047** | **rs17487130** | **T** | **C** | **1.06** | **1.04-1.07** | **4.16e-14** | **0.40** | **0.92** | **Novel** |
| **19p13.12** | ***TECR*** | **19:14533044** | **rs11671111** | **T** | **C** | **0.96** | **0.95-0.98** | **1.56e-08** | **0.25** | **1.6** | **Novel** |
| 19q13.32 | *FOXA3* | 19:45876389 | rs10409222 | T | C | 1.05 | 1.03-1.06 | 2.97e-08 | 0.17 | 0.8 | [4] |
| **19q13.42** | ***LENG8*** | **19:54471384** | **rs2287822** | **A** | **G** | **1.04** | **1.02-1.05** | **1.05e-08** | **0.30** | **1.33** | **Novel** |
| **21q22.11** | ***SLC5A3*** | **21:33662166** | **rs3827180** | **A** | **G** | **1.04** | **1.03-1.05** | **2.48e-09** | **0.28** | **1.3** | **Novel** |
| **Xp22.11** | ***PCYT1B*** | **23:24653216** | **rs5944665** | **A** | **G** | **0.97** | **0.96-0.98** | **3.36e-08** | **0.60** | **NA** | **Novel** |
| **Xq21.1** | ***ITM2A*** | **23:79455466** | **rs191015078** | **T** | **C** | **1.05** | **1.03-1.07** | **4.85e-08** | **0.12** | **NA** | **Novel** |
| **Xq21.1** | **Empty** | **23:82687578** | **rs111872003** | **A** | **T** | **0.92** | **0.89-0.95** | **3.11e-08** | **0.04** | **NA** | **Novel** |

| | | | | | | | | | | | |
|---|---|---|---|---|---|---|---|---|---|---|---|
| **Xq23** | ***CHRDL1*** | **23:110640115** | **rs7884700** | **G** | **A** | **1.04** | **1.03-1.05** | **9.64e-12** | **0.40** | **NA** | **Novel** |

Candidate gene, a gene at a new locus the biological function of which is likely to explain the LDH association; CHR: POS, chromosome and position (genome build hg38); rsid; SNP markers identification number; EA, effect allele; OA, other allele; OR, odds ratio; 95% Cl, odds ratio 95% confidence interval; EAF, effect allele frequency; Fin Enric., enrichment in Finns (calculated FIN AF/NFEE AF in the Genome Aggregation Database [gnomAD], FIN AF is the allele frequency in Finns and NFEE AF is the allele frequency in Europeans (does not include Finns or Estonians)); NA, not available; REF, reference to literature reporting an LDH association in the vicinity (± 1MB) of the lead variant, if lead variant was novel it was also bolded. *, for this variant p-value and OR have been adjusted for the effect of original lead variant at the locus through conditional analysis, the original lead variant is always above the conditional variant in the table.

**Table S3** Genomic locations and potential biological role of the association signals. Candidate gene, a gene at a novel locus which biological function is likely to explain the lumbar disc herniation association; rsid, SNP markers identification number, OR, odds ratio, Variant type, genomic location of associated variant, Het pval, p-value for heterogeneity. For the loci that had not been reported in association with LDH in prior studies, we determined a potential candidate gene with a relevant biological function with the help of literature and databases (Genbank[8], UniProt[9], GTEX-Portal[10]). Even though potential genes have been systematically identified, there is little clarity regarding their causality, so further studies are needed.

| Candidate gene | rsid | OR | Variant type | Het Pval | A possible function related to LDH pathogenesis |
|---|---|---|---|---|---|
| ***ALPL*** | rs150211890 | 1.07 | intron | 0.178 | *ALPL* boosts inorganic phosphate local rates, promotes mineralization, and lowers extracellular pyrophosphate concentration, which acts as a mineral formation inhibitor[11]. Greater *ALPL* activity has been associated with IVD degeneration and calcification[12], as according to findings from earlier studies that found that IVD's with degenerative changes had higher levels of *ALPL* activity and calcification potential than control IVD's without degenerative changes[13]. |
| ***COL11A1*** | rs3056624 | 1.05 | intron | 0.263 | - |
| ***NGF*** | rs4644491 | 0.96 | intron | 0.215 | *NGF* promotes collateral sprouting of the peripheral sensory nerves and axonal regeneration in the central nervous system. The IVD may experience nociceptive nerve ingrowth as a result of *NGF* production[14,15]. |
| ***COLGALT2*** | rs3010043 | 0.95 | intron | 0.612 | - |
| ***PTPRC*** | rs28599571 | 1.04 | intergenic | 0.738 | - |
| ***TGFB2*** | rs779040 | 0.97 | intergenic | 0.0225 | This gene encodes a secreted ligand of the TGF-β superfamily of proteins. Ligands of this family bind various TGF-β receptors leading to recruitment and activation of *SMAD* family transcription factors that regulate gene expression[16]. TGF-β signaling is necessary for the development and growth of IVD, and can play a protective role in the restoration of IVD tissues by stimulating matrix synthesis, inhibiting matrix catabolism, inflammatory response and cell loss. However, excessive activation of TGF-β signaling is detrimental to the IVD, and inhibition of the aberrant TGF-β signaling can delay IVD degeneration[17]. |
| ***HHIPL2*** | rs35455442 | 0.96 | intron | 0.539 | Iron excess can limit *HHIPL2* gene expression and decrease osteoblastic activity in human MG-63 cells[18]. *HHIPL2* is an inhibitor of the hedgehog signalling pathway, this pathway has previously been linked together with *GLI1* to hypertrophy of the ligamentum flavum[19]. *HHIPL2* has also been found to be involved in extracellular matrix synthesis in IVDs[20]. |
| ***GFPT1*** | rs12997836 | 1.04 | intergenic | 0.298 | - |
| ***TGFA*** | rs3849386 | 0.95 | intron | 0.402 | - |
| ***GPR1*** | rs78826721 | 0.94 | intron | 0.171 | *GPR1* is a chemerin receptor that may mediate chemerin activities in inflammation[21]. Chemerin can induce many inflammatory cytokines, activate NF-kB signaling pathway[22]. *GPR1* deficiency has also been found to reduce bone mass and BMD (osteopenia) in mouse studies[23,24]. |
| ***HYAL2*** | rs41308273 | 0.92 | intron | 0.498 | *HYAL2* codes for hyaluronidase that degrades hyaluronan. Degradation of hualyronan is not a benign event, as it reduces the size of the aggrecan aggregates and can promote their diffusion[25,26]. |
| ***PDZRN3*** | rs11914834 | 0.97 | regulatory region | 0.984 | Possibly a negative regulator of Wnt/β-catenin signaling[27]. Activation of the Wnt/β-catenin pathway has been found to be associated with endplate degeneration, increased IVD cell senescence, and extracellular matrix degradation[28,29]. |
| ***ADCY5*** | rs1965290 | 0.97 | intron | 0.0391 | *ADCY5* has a role in osteogenic differentiation and may be associated with vertebral fractures and BMD[30,31]. |
| ***NCK1*** | rs13321721 | 1.04 | intron | 0.822 | *NCK1* inhibits the ability of *DCC* to induce neurite outgrowth, thus playing role in axonal guidance[32]. |
| ***SHOX2*** | rs5853827 | 0.96 | intron | 0.492 | SHOX2 prevents the onset of early chondrocyte differentiation and controls the transition of chondrocytes from mature to hypertrophic by regulating the expression of transcription factors, such as *SOX6*, *SOX9*, and *RUNX2*. Inhibition of *SHOX2* decreases expression of aggrecan and collagen II in nucleus pulposus and thus can lead to degenerative changes in IVDs[33]. |
| ***FGFR3*** | rs35313041 | 1.05 | intron | 0.0086 | - |
| ***IBSP*** | rs10019020 | 1.04 | intergenic | 0.439 | Dysregulation of *IBSP* may induce de-adhesion, characterized by disruption of extracellular matrix organization and focal adhesions, which can accelerate IVD degeneration[34]. |

| | | | | | |
|---|---|---|---|---|---|
| ***HDAC3*** | rs5871786 | 0.97 | intron | 0.729 | *HDAC3* is a highly pleiotropic epigenetic regulator. TGF-β signaling pathway is regulated by *HDAC*s and acetylation, and a number of TGF-β-induced genes involved in the remodelling and regulation of the extracellular matrix are regulated by *HDAC* inhibition. HDAC3 plays a central role in these reactions.[35,36]. *HDAC3* can also mediate HIF-1α stability in IVDs[37]. In addition, it has been found to interact with *RUNX2* to repress the osteocalcin promoter and thus regulate osteoblast differentiation[38]. |
| ***FGF18*** | rs4302608 | 0.97 | downstream | 0.897 | *FGF18* is a well-characterised anabolic growth factor involved in cartilage homeostasis[39]. Its function in the IVD is not fully understood, but it has been found that *FGF18* could delay the degeneration of the IVD by inhibiting the apoptosis of NPs and the expression of matrix-degrading enzymes[40]. |
| ***TRIM38*** | rs9393692 | 0.97 | upstream | 0.831 | *TRIM38* protects chondrocytes from IL-1β-induced apoptosis and degeneration via negatively modulating nuclear factor (NF)-κB signalling[41]. IL-1β is also a key risk factor for intervertebral disc degeneration[42]. |
| ***HLA*** | rs1611653 | 1.04 | intron | 0.224 | - |
| ***HLA*** | rs2844608 | 0.96 | intron | 0.0185 | - |
| ***HLA*** | rs9273873 | 1.06 | noncoding transcript exon variant | 0.491 | - |
| ***ILRUN*** | rs2744939 | 1.05 | upstream | 0.0231 | - |
| ***CDC5L*** | rs6929734 | 1.04 | regulatory region | 0.0318 | - |
| ***TBX18*** | rs2224214 | 1.04 | intergenic | 0.664 | *TBX18* acts as a transcriptional repressor involved in the developmental processes of the vertebral column. In studies conducted in mice, the *TBX18* null mutation has been found to affect perinatal lethality with shortened axial skeletons; effects included kinks in the thoracic vertebral column, malformed, flattened discs, and vertebral bodies, expanded pedicles, and transverse processes[43]. |
| ***TWIST1*** | rs34895285 | 0.96 | intron | 0.202 | - |
| ***IGFBP3*** | rs788747 | 1.05 | upstream gene variant | 0.578 | - |
| ***ELN*** | rs10227463 | 0.97 | intron | 0.768 | Elastin is important structural component of ECM and present in connective tissues. In IVD's elastin may function to restore lamellar structure under radial loads that potentially cause delamination. The observed increases in elastin with degeneration may reflect both newly synthesized elastic fibers and/or degraded elastin peptides trapped in matrix as part of the degenerative cascade[44]. |
| ***ZC3HC1*** | rs11556924 | 1.04 | missense | 0.286 | *ZC3HC1* encodes an F-box-containing protein that is a component of an SCF-type E3 ubiquitin ligase complex that regulates the onset of cell division. The G2/M transition in the cell cycle requires the interaction of the proteins cyclin B1 and cyclin-dependent kinase 1. The activated ubiquitin ligase complex targets the protein cyclin B1 for degradation, preventing this transition to mitosis[45]. |
| ***STMP1*** | rs2551776 | 0.97 | intron | 0.318 | *STMP1* potentially has a role in bone remodeling, it has previously been associated with Paget's disease of bone. It also potentially plays a role in regulating the NLRP3 inflammasome[46,47]. |
| ***VEST1*** | rs2164198 | 1.06 | intron | 0.0367 | - |
| ***GSDMC*** | rs7814941 | 0.91 | splice region variant | 0.00552 | - |
| ***TUSC1*** | rs7019841 | 0.96 | intergenic | 0.696 | The connection between *TUSC1* and LDH is not yet known, and it has become known mainly as a potential tumor suppressor in human cancers. It has been found to suppress cell proliferation and cell cycle progression[48]. It is possible that it also affects intervertebral disc degeneration by regulating these pathways, but there is no research evidence for this yet. |
| ***PHF2*** | rs58723578 | 1.07 | intergenic | 0.034 | - |
| ***LPAR1*** | rs10980637 | 1.05 | intron | 0.395 | The increased expression of and *LPAR1* has been associated with the fibrosis and hypertrophy of the ligamentum flavum[49]. *LPAR1* deletion can also cause neurodevelopmental disorders such as demyelination diseases and neuropathic pain[50]. |
| ***DNM1*** | rs9644952 | 1.04 | intron | 0.807 | *DNM1* plays a central role in the transmission of nociceptive messages within the nociceptive circuits in the dorsal horn of the spinal cord, where *DNM1*-mediated endocytosis of synaptic vesicles enables sustained neurotransmission[51]. |

| | | | | | |
|---|---|---|---|---|---|
| ***AKR1C1*** | rs536435747 | 0.94 | 3 prime UTR | 0.513 | *AKR1C1* is possibly involved in the regulation of inflammatory factors; it is possibly involved in the regulation of *IL-1*, *TNF*, and *TGFB1*-related pathways[52,53]. |
| ***MKX*** | rs2808290 | 1.05 | intergenic | 0.211 | - |
| ***CHST3*** | rs4284332 | 1.07 | intron | 0.741 | - |
| ***HTRA1*** | rs2672590 | 0.95 | intron | 0.521 | *HTRA1* is serine protease that antagonizes TGF-β signaling and has a role in regulation of bone formation[54]. It may also degrade proteoglycans such as aggrecan, decorin, and fibromodulin, and thus contribute to the cartilage degradation[55]. |
| ***ARNTL*** | rs12295734 | 1.04 | intergenic | 0.195 | - |
| ***SOX6*** | rs9787942 | 0.95 | intron | 0.507 | - |
| ***MYEOV*** | rs144549742 | 0.91 | intergenic | 0.469 | *MYEOV* has been observed to interact with SOX9 and potentially enhance its transactivity[56]. |
| ***SIK2*** | rs77651758 | 0.92 | intron | 0.67 | Parathyroid hormone can modulate *SIK2* to promote bone formation and resorption, by inhibiting it's activity[57]. |
| ***SOX5*** | rs11831278 | 1.09 | intron | 0.805 | - |
| ***GLI1*** | rs871871 | 0.96 | intron | 0.635 | The growth plate mediates bone growth where *SOX9* and *GLI* factors control chondrocyte proliferation, differentiation and entry into hypertrophy. *GLI1* functions as an activator which is highly expressed in proliferating chondrocytes and perichondrium adjacent to the prehypertrophic and hypertrophic zones[58]. |
| ***KMT5A*** | rs1626703 | 1.04 | splice region | 0.578 | *KMT5A* inhibits oxidative stress induced autophagy and glial scar formation in astrocytes via the *KEAP1-NRF2-ARE* signaling pathway[59]. *KMT5A* also has a role in TGF-β response regulation where it is important for turning of activation of *SMAD2*[60]. |
| ***DIAPH3*** | rs340208 | 1.04 | intron | 0.445 | *DIAPH3* is involved in cell migration, axon guidance and neuritogenesis[61]. It has been found to affect spine length and density of neurons[62]. |
| ***PAX9*** | rs11848465 | 0.95 | intron | 0.999 | - |
| ***SERPINA1*** | rs28929474 | 0.86 | missense | 0.762 | - |
| ***SMAD3*** | rs4776880 | 0.95 | intron | 0.0729 | - |
| ***CA10*** | rs59704663 | 1.48 | upstream gene variant | 0.156 | *CA10* blocks the binding of heparan sulfate to neurexin[63], which possibly raises neurexin surface levels[64]. Neurexins are pre-synaptic cell adhesion molecules that play a role in connecting neurons at synapses. Heparan sulfate has been found to potentially expand the interactome of neurexins, and they also play a role in fine-tuning synaptic transmission[65]. *CA10* is expressed especially in the central nervous system, and it has been associated with chronic pain in previous studies[64,66]. |
| ***NPC1*** | rs1788760 | 0.97 | intron | 0.89 | *NPC1* has a role in cholesterol trafficking, which has a role in mTOR regulation. Changes in this pathway have been found to lead to increased angiogenesis[67,68], which in turn can lead to IVD innervation[69]. |
| ***SETBP1*** | rs8088824 | 0.95 | intergenic | 0.185 | *SETBP1* may have a regulatory role in Wnt/β-catenin pathway in neural cells[70], *SETBP1* has also been associated with adolescent idiopathic scoliosis[71]. |
| ***DCC*** | rs17487130 | 1.06 | intron | 0.862 | In diseased IVD's *DCC* might play an important role in neurovascular ingrowth/axonal guidance[32,72]. |
| ***TECR*** | rs11671111 | 0.96 | intron | 0.769 | *TECR* is involved in both the production of very long-chain fatty acids for sphingolipid synthesis and the degradation of the sphingosine moiety in sphingolipids through the sphingosine 1-phosphate metabolic pathway[73]. It has been proposed that *TECR*, as a synaptic glycoprotein, may have a specialized, as yet unknown, function in the nervous system that affects communication between neurons or synaptic plasticity[74]. |
| ***FOXA3*** | rs10409222 | 1.05 | downstream | 0.394 | - |
| **LENG8** | rs2287822 | 1.04 | intron | 0.394 | *LENG8* regulates inflammation cascades and can interact with many proteins involved in rheumatoid arthritis, like interleukin-18[75]. Interleukin-18 has also been implicated in the pathogenesis of IVD degeneration[76]. |
| ***SLC5A3*** | rs3827180 | 1.04 | intron | 0.101 | *SLC5A3* cotransports Na+ and myo-inositol, a critical osmotic regulator for cells[77]. Changes in the concentration of myo-inositol have been observed in connection with IVD degeneration. *SLC5A3* may contribute to the imbalance of disc osmotic activity in degenerative diseases[78]. *SLC5A3* is also attributed a specific role regarding the p53-mediated G1 checkpoint activation and/or the p38 MAPK-dependent G2/M arrest; these cascades are therefore triggered by high osmolality[79]. |
| ***PCYT1B*** | rs5944665 | 0.97 | intron | 0.925 | *PCYT1B* could have a role in axon regeneration and branching. It is involved in the regulation of phosphatidylcholine |

| | | | | | |
|---|---|---|---|---|---|
| | | | | | biosynthesis, that has been proposed to be key regulatory mechanism for axon regeneration[80–82]. |
| ***ITM2A*** | rs191015078 | 1.05 | intergenic | 0.536 | *ITM2A* may inhibit the initiation of chondrogenesis and elevated expression of *ITM2A* can therefore be linked to poorer chondrogenic differentiation potential[83,84]. |
| **Empty** | rs111872003 | 0.92 | intergenic | 0.176 | - |
| ***CHRDL1*** | rs7884700 | 1.04 | downstream | 0.438 | *CHRDL1* can possibly influence *BMP-4-SMAD1/5/9* pathways activity and through that it might have an important role in hBMSCs osteogenic differentiation and bone remodeling[85]. |

**Table S4.** Effect differences between meta-analysis and SURG GWAS. Lead variants are variants that were observed in the meta-analysis. Beta estimates and their standard errors were collected from the GWAS result files of meta-analysis and SURG GWAS. Values of $P_{diff}$<0.05 were considered statistically significant.

| Lead variant | Candidate Gene | Beta meta | Beta SURG | Se meta | Se SURG | $P_{diff}$ |
|---|---|---|---|---|---|---|
| 1:21559185:T:G | *ALPL* | 0.069 | 0.077 | 0.012 | 0.031 | 0.83 |
| **1:102882172:GTATT:G** | ***COL11A1*** | **0.048** | **0.099** | **0.006** | **0.017** | **0.043** |
| 1:115310363:G:A | *NGF* | -0.036 | -0.051 | 0.006 | 0.017 | 0.39 |
| 1:183973041:A:G | *COLGALT2* | -0.055 | -0.036 | 0.007 | 0.021 | 0.40 |
| 1:198768851:G:T | *PTPRC* | 0.041 | 0.060 | 0.006 | 0.017 | 0.30 |
| 1:218924545:G:C | *TGFB2* | -0.035 | -0.057 | 0.006 | 0.017 | 0.23 |
| 1:222541797:C:A | *HHIPL2* | -0.046 | -0.058 | 0.006 | 0.017 | 0.50 |
| **2:69304791:T:C** | ***GFPT1*** | **0.043** | **0.089** | **0.006** | **0.017** | **0.011** |
| **2:70496764:C:T** | ***TGFA*** | **-0.049** | **-0.116** | **0.006** | **0.018** | **0.0004** |
| 2:206137590:A:G | *GPR1* | -0.065 | -0.080 | 0.011 | 0.026 | 0.59 |
| 3:50380254:T:A | *HYAL2* | -0.086 | -0.068 | 0.013 | 0.032 | 0.61 |
| 3:73200973:C:T | *PDZRN3* | -0.033 | -0.043 | 0.006 | 0.017 | 0.56 |
| 3:123568959:T:C | *ADCY5* | -0.034 | -0.008 | 0.006 | 0.017 | 0.15 |
| 3:136490708:A:G | *NCK1* | 0.041 | 0.057 | 0.007 | 0.020 | 0.45 |
| 3:158478549:A:ATCC | *SHOX2* | -0.036 | -0.005 | 0.006 | 0.018 | 0.11 |
| 4:1694376:C:T | *FGFR3* | 0.053 | 0.072 | 0.006 | 0.017 | 0.30 |
| 4:87779677:G:A | *IBSP* | 0.035 | 0.064 | 0.006 | 0.017 | 0.10 |
| 5:141735121:GT:G | *HDAC3* | -0.033 | -0.046 | 0.006 | 0.017 | 0.50 |
| 5:171413500:A:G | *FGF18* | -0.034 | -0.029 | 0.006 | 0.017 | 0.80 |
| 6:26276422:A:G | *TRIM38* | -0.034 | -0.052 | 0.006 | 0.018 | 0.34 |
| 6:29873925:G:C | *HLA* | 0.043 | 0.055 | 0.007 | 0.017 | 0.50 |
| 6:31279637:C:T | *HLA* | -0.045 | -0.075 | 0.006 | 0.017 | 0.10 |
| 6:32661873:T:C | *HLA* | 0.059 | 0.073 | 0.010 | 0.024 | 0.59 |
| 6:34580429:T:A | *ILRUN* | 0.050 | 0.068 | 0.008 | 0.021 | 0.41 |
| 6:44478351:G:T | *CDC5L* | 0.036 | 0.064 | 0.006 | 0.017 | 0.12 |
| 6:84938145:C:T | *TBX18* | 0.044 | 0.073 | 0.006 | 0.017 | 0.12 |
| **7:19442778:TA:T** | ***TWIST1*** | **-0.043** | **-0.081** | **0.007** | **0.017** | **0.041** |
| **7:45987132:T:G** | ***IGFBP3*** | **0.048** | **0.123** | **0.006** | **0.017** | **0.00003** |
| 7:73714641:C:T | *ELN* | -0.035 | -0.050 | 0.006 | 0.017 | 0.41 |
| 7:130023656:C:T | *ZC3HC1* | 0.035 | 0.065 | 0.006 | 0.018 | 0.10 |
| 7:135418700:C:T | *STMP1* | -0.035 | -0.033 | 0.006 | 0.018 | 0.92 |
| 8:68665207:A:G | *VEST1* | 0.055 | 0.078 | 0.008 | 0.026 | 0.40 |
| **8:129706613:A:G** | ***GSDMC*** | **-0.093** | **-0.170** | **0.008** | **0.023** | **0.001** |
| 9:25398495:A:G | *TUSC1* | -0.036 | -0.018 | 0.006 | 0.017 | 0.31 |
| 9:93911476:C:T | *PHF2* | 0.067 | 0.105 | 0.010 | 0.026 | 0.18 |
| 9:110930238:C:T | *LPAR1* | 0.046 | 0.047 | 0.008 | 0.022 | 0.97 |
| 9:128236873:C:A | *DNM1* | 0.040 | 0.060 | 0.007 | 0.020 | 0.34 |
| 10:4989436:A:AC | *AKR1C1* | -0.058 | -0.084 | 0.009 | 0.025 | 0.33 |
| 10:27611953:C:T | *MKX* | 0.048 | 0.067 | 0.006 | 0.017 | 0.31 |
| **10:71974194:T:C** | ***CHST3*** | **0.071** | **0.122** | **0.006** | **0.017** | **0.005** |
| 10:122475088:A:C | *HTRA1* | -0.046 | -0.051 | 0.007 | 0.021 | 0.84 |
| 11:13270734:G:C | *ARNTL* | 0.037 | 0.038 | 0.007 | 0.018 | 0.98 |
| 11:15673785:T:C | *SOX6* | -0.055 | -0.082 | 0.007 | 0.018 | 0.16 |
| 11:69208032:T:A | *MYEOV* | -0.094 | -0.089 | 0.014 | 0.035 | 0.89 |
| 11:111459420:C:T | *SIK2* | -0.082 | -0.090 | 0.013 | 0.032 | 0.82 |
| **12:23807795:C:T** | ***SOX5*** | **0.085** | **0.134** | **0.008** | **0.022** | **0.041** |
| **12:57825898:G:A** | ***GLI1*** | **-0.038** | **-0.092** | **0.006** | **0.017** | **0.004** |
| 12:123226288:A:C | *KMT5A* | 0.042 | 0.033 | 0.007 | 0.020 | 0.65 |
| 13:59904471:A:T | *DIAPH3* | 0.037 | 0.036 | 0.007 | 0.019 | 0.99 |
| 14:37002513:C:T | *PAX9* | -0.052 | -0.089 | 0.007 | 0.020 | 0.07 |
| 14:94378610:C:T | *SERPINA1* | -0.153 | -0.239 | 0.022 | 0.065 | 0.27 |
| 15:67072653:G:A | *SMAD3* | -0.056 | -0.057 | 0.006 | 0.017 | 0.99 |
| 17:52164544:G:A | *CA10* | 0.389 | 0.073 | 0.068 | 0.339 | 0.36 |
| 18:23557478:A:G | *NPC1* | -0.035 | -0.028 | 0.006 | 0.018 | 0.73 |
| 18:44571296:C:T | *SETBP1* | -0.051 | -0.039 | 0.007 | 0.021 | 0.59 |
| 18:53189047:C:T | *DCC* | 0.054 | 0.055 | 0.007 | 0.017 | 0.97 |
| 19:14533044:C:T | *TECR* | -0.038 | -0.058 | 0.007 | 0.018 | 0.31 |
| 19:45876389:C:T | *FOXA3* | 0.046 | 0.080 | 0.008 | 0.024 | 0.17 |
| 19:54471384:G:A | LENG8 | 0.037 | 0.030 | 0.006 | 0.018 | 0.73 |
| 21:33662166:G:A | *SLC5A3* | 0.039 | 0.005 | 0.007 | 0.018 | 0.07 |
| 23:24653216:G:A | *PCYT1B* | -0.028 | -0.006 | 0.005 | 0.014 | 0.13 |
| 23:79455466:C:T | *ITM2A* | 0.051 | 0.079 | 0.009 | 0.021 | 0.23 |
| 23:82687578:T:A | Empty | -0.080 | -0.089 | 0.015 | 0.042 | 0.85 |
| 23:110640115:A:G | CHRDL1 | 0.035 | 0.053 | 0.005 | 0.014 | 0.24 |

**Table S5 Genome-wide significant ($p<5x10^{-8}$) lead variants were associated with LDH related surgical operation.** There are 7347 operated LDH cases and 270 964 controls in the analysis, consisting of data from FinnGen.

| CHR:POS | Candidate gene | rsid | EA | OA | OR 95% CI | EAF | pval | pval meta | Ref. |
|---|---|---|---|---|---|---|---|---|---|
| **1:102875067** | ***COL11A1*** | **rs1318756** | **C** | **T** | **1.10 (1.07-1.13)** | **0.53** | **2.41e-08** | **2.00e-15** | **Novel** |
| **2:70465425** | ***TGFA*** | **rs3732247** | **T** | **C** | **0.88 (0.85-0.92)** | **0.34** | **2.69e-11** | **1.35e-11** | **Novel** |
| **7:19508326** | ***TWIST1*** | **rs6944632** | **G** | **A** | **0.91 (0.88-0.94)** | **0.61** | **2.05e-09** | **1.74e-07** | **Novel** |
| 7:45988978 | *IGFBP3* | rs1723939 | T | C | 1.13 (1.10-1.16) | 0.49 | 1.15e-13 | 2.07e-15 | [4,6] |
| 8:129707472 | *GSDMC* | rs7816131 | T | A | 0.85 (0.81-0.89) | 0.18 | 9.61e-13 | 3.52e-32 | [4,6] |
| 10:71977366 | *CHST3* | rs4148926 | C | G | 1.13 (1.10-1.17) | 0.55 | 1.57e-12 | 1.73e-20 | [4,6] |
| **12:23823019** | ***SOX5*** | **rs11834104** | **T** | **G** | **1.14 (1.10-1.18)** | **0.16** | **5.82-09** | **8.81e-25** | **Novel** |
| **17:71514369** | ***SOX9*** | **rs7225015** | **C** | **A** | **0.89 (0.86-0.93)** | **0.31** | **8.67e-10** | **2.19e-05** | **Novel** |

CHR: POS, chromosome and position (genome build hg38); Candidate gene, a gene at a new locus whose biological function is likely to explain the LDH related surgical operation association; rsid; SNP markers identification number; EA, effect allele; OA, other allele; OR 95% CI, odds ratio and it's 95% confidence interval; EAF, effect allele frequency; pval, p-value, pval meta, variants p-value in LDH meta-analysis; REF, reference article in which a LDH related surgical operation association was observed +/- 1 Mb in the vicinity of the lead variant.

**Table S6** Cumulative patient morbidity and cumulative surgeries observed for every LDH associated variant. The analysis was done by extracting the genotypes corresponding to the variants associated with LDH in FinnGen's Sandbox environment and combining them with the health register data. Cumulative morbidities are specified with same ICD-codes as in Table S1. The prevalence of LDH cases was 12.2% and prevalence of surgical patients was 2.6%.

| Candidate gene | rsid | CHR:POS | EA | OA | Cumulative Morbidity EA | Cumulative Morbidity OA | Cumulative Surgeries EA | Cumulative Surgeries OA |
|---|---|---|---|---|---|---|---|---|
| ***ALPL*** | rs150211890 | 1:21559185 | **G** | T | **0.147** | 0.121 | **0.028** | 0.024 |
| ***COL11A1*** | rs3056624 | 1:102882172 | **G** | GTATT | **0.126** | 0.118 | **0.026** | 0.024 |
| ***NGF*** | rs4644491 | 1:115310363 | **A** | G | **0.119** | 0.122 | **0.024** | 0.026 |
| ***COLGALT2*** | rs3010043 | 1:183973041 | **G** | A | **0.118** | 0.125 | **0.024** | 0.026 |
| ***PTPRC*** | rs28599571 | 1:198768851 | **T** | G | **0.124** | 0.120 | **0.026** | 0.024 |
| ***TGFB2*** | rs779040 | 1:218924545 | **C** | G | **0.118** | 0.122 | **0.024** | 0.026 |
| ***HHIPL2*** | rs35455442 | 1:222541797 | **A** | C | **0.116** | 0.122 | **0.023** | 0.026 |
| ***GFPT1*** | rs12997836 | 2:69304791 | **C** | T | **0.126** | 0.120 | **0.026** | 0.024 |
| ***TGFA*** | rs3849386 | 2:70496764 | **T** | C | **0.114** | 0.122 | **0.021** | 0.026 |
| ***GPR1*** | rs78826721 | 2:206137590 | **G** | A | **0.107** | 0.121 | **0.020** | 0.026 |
| ***HYAL2*** | rs41308273 | 3:50380254 | **A** | T | **0.111** | 0.121 | **0.023** | 0.026 |
| ***PDZRN3*** | rs11914834 | 3:73200973 | **T** | C | **0.116** | 0.122 | **0.023** | 0.026 |
| ***ADCY5*** | rs1965290 | 3:123568959 | **C** | T | **0.122** | 0.119 | **0.026** | 0.024 |
| ***NCK1*** | rs13321721 | 3:136490708 | **G** | A | **0.125** | 0.121 | **0.026** | 0.024 |
| ***SHOX2*** | rs5853827 | 3:158478549 | **ATCC** | A | **0.116** | 0.121 | **0.025** | 0.024 |
| ***FGFR3*** | rs35313041 | 4:1694376 | **T** | C | **0.125** | 0.119 | **0.026** | 0.024 |
| ***IBSP*** | rs10019020 | 4:87779677 | **A** | G | **0.126** | 0.120 | **0.026** | 0.024 |
| ***HDAC3*** | rs5871786 | 5:141735121 | **G** | GT | **0.117** | 0.122 | **0.023** | 0.026 |
| ***FGF18*** | rs4302608 | 5:171413500 | **G** | A | **0.118** | 0.122 | **0.024** | 0.026 |
| ***TRIM38*** | rs9393692 | 6:26276422 | **G** | A | **0.120** | 0.121 | **0.024** | 0.026 |
| ***HLA*** | rs1611653 | 6:29873925 | **C** | G | **0.124** | 0.120 | **0.025** | 0.024 |
| ***HLA*** | rs2844608 | 6:31279637 | **T** | C | **0.116** | 0.122 | **0.023** | 0.026 |
| ***HLA*** | rs9273873 | 6:32661873 | **C** | T | **0.131** | 0.121 | **0.026** | 0.024 |
| ***ILRUN*** | rs2744939 | 6:34580429 | **A** | T | **0.132** | 0.120 | **0.029** | 0.024 |
| ***CDC5L*** | rs6929734 | 6:44478351 | **T** | G | **0.122** | 0.120 | **0.026** | 0.024 |
| ***TBX18*** | rs2224214 | 6:84938145 | **T** | C | **0.127** | 0.122 | **0.026** | 0.024 |
| ***TWIST1*** | rs34895285 | 7:19442778 | **T** | TA | **0.116** | 0.122 | **0.022** | 0.026 |
| ***IGFBP3*** | rs788747 | 7:45987132 | **G** | T | **0.126** | 0.120 | **0.028** | 0.023 |
| ***ELN*** | rs10227463 | 7:73714641 | **T** | C | **0.116** | 0.122 | **0.023** | 0.026 |
| ***ZC3HC1*** | rs11556924 | 7:130023656 | **T** | C | **0.126** | 0.120 | **0.026** | 0.024 |
| ***STMP1*** | rs2551776 | 7:135418700 | **T** | C | **0.120** | 0.122 | **0.024** | 0.026 |
| ***VEST1*** | rs2164198 | 8:68665207 | **G** | A | **0.126** | 0.121 | **0.023** | 0.026 |
| ***GSDMC*** | rs7814941 | 8:129706613 | **G** | A | **0.101** | 0.122 | **0.017** | 0.024 |
| ***TUSC1*** | rs7019841 | 9:25398495 | **G** | A | **0.118** | 0.122 | **0.024** | 0.026 |
| ***PHF2*** | rs58723578 | 9:93911476 | **T** | C | **0.147** | 0.121 | **0.029** | 0.024 |
| ***LPAR1*** | rs10980637 | 9:110930238 | **T** | C | **0.133** | 0.120 | **0.026** | 0.024 |
| ***DNM1*** | rs9644952 | 9:128236873 | **A** | C | **0.127** | 0.121 | **0.027** | 0.024 |
| ***AKR1C1*** | rs536435747 | 10:4989436 | **AC** | A | **0.113** | 0.121 | **0.019** | 0.026 |
| ***MKX*** | rs2808290 | 10:27611953 | **T** | C | **0.123** | 0.120 | **0.025** | 0.024 |
| ***CHST3*** | rs4284332 | 10:71974194 | **C** | T | **0.127** | 0.118 | **0.027** | 0.024 |
| ***HTRA1*** | rs2672590 | 10:122475088 | **C** | A | **0.116** | 0.121 | **0.024** | 0.026 |
| ***ARNTL*** | rs12295734 | 11:13270734 | **C** | G | **0.123** | 0.119 | **0.025** | 0.024 |
| ***SOX6*** | rs9787942 | 11:15673785 | **C** | T | **0.116** | 0.125 | **0.023** | 0.026 |
| ***MYEOV*** | rs144549742 | 11:69208032 | **A** | T | **0.095** | 0.121 | **0.016** | 0.024 |
| ***SIK2*** | rs77651758 | 11:111459420 | **T** | C | **0.119** | 0.121 | **0.026** | 0.024 |
| ***SOX5*** | rs11831278 | 12:23807795 | **T** | C | **0.134** | 0.120 | **0.029** | 0.024 |
| ***GLI1*** | rs871871 | 12:57825898 | **A** | G | **0.118** | 0.121 | **0.023** | 0.026 |
| ***KMT5A*** | rs1626703 | 12:123226288 | **C** | A | **0.122** | 0.119 | **0.025** | 0.024 |
| ***DIAPH3*** | rs340208 | 13:59904471 | **T** | A | **0.124** | 0.118 | **0.025** | 0.024 |
| ***PAX9*** | rs11848465 | 14:37002513 | **T** | C | **0.117** | 0.121 | **0.024** | 0.026 |
| ***SERPINA1*** | rs28929474 | 14:94378610 | **T** | C | **0.067** | 0.121 | **0.023** | 0.024 |
| ***SMAD3*** | rs4776880 | 15:67072653 | **A** | G | **0.116** | 0.122 | **0.024** | 0.026 |
| ***CA10*** | rs59704663 | 17:52164544 | **A** | G | **-** | 0.121 | **-** | 0.024 |
| ***NPC1*** | rs1788760 | 18:23557478 | **G** | A | **0.119** | 0.122 | **0.024** | 0.026 |
| ***SETBP1*** | rs8088824 | 18:44571296 | **T** | C | **0.119** | 0.124 | **0.024** | 0.026 |
| ***DCC*** | rs17487130 | 18:53189047 | **T** | C | **0.127** | 0.120 | **0.026** | 0.024 |
| ***TECR*** | rs11671111 | 19:14533044 | **T** | C | **0.117** | 0.121 | **0.023** | 0.026 |
| ***FOXA3*** | rs10409222 | 19:45876389 | **T** | C | **0.128** | 0.121 | **0.026** | 0.024 |
| ***LENG8*** | rs2287822 | 19:54471384 | **A** | G | **0.128** | 0.120 | **0.027** | 0.024 |
| ***SLC5A3*** | rs3827180 | 21:33662166 | **A** | G | **0.127** | 0.120 | **0.025** | 0.024 |
| ***PCYT1B*** | rs5944665 | 23:24653216 | **A** | G | **-** | - | **-** | - |
| ***ITM2A*** | rs191015078 | 23:79455466 | **T** | C | **-** | - | **-** | - |
| ***Empty*** | rs111872003 | 23:82687578 | **A** | T | **-** | - | **-** | - |
| ***CHRDL1*** | rs7884700 | 23:110640115 | **G** | A | **-** | - | **-** | - |

**Table S7** Genetic correlations for all 438 phenotypes. Genetic correlations were calculated using LDSC-software[86]. All traits were extracted from the GWAS database provided by the MRC Integrative Epidemiology Unit (IEU). RG, genetic correlation coefficient value; pFDR, false discovery rate-corrected p-value.

| Trait | RG | se | p | pFDR |
|---|---|---|---|---|
| **Concentration of small VLDL particles** | 0.1533 | 0.0595 | 0.010 | 0.017 |
| **Knee pain for 3+ months** | 0.4914 | 0.0831 | 3.38e-09 | 1.00e-08 |
| **Time spent driving** | 0.2175 | 0.0336 | 1.00e-10 | 3.48e-10 |
| **Smoking status: Current** | 0.3477 | 0.0283 | 1.21e-34 | 1.77e-33 |
| **Cholesterol in small VLDL** | 0.0852 | 0.036 | 0.018 | 0.029 |
| **Vascular/heart problems diagnosed by doctor: Stroke** | 0.3332 | 0.0742 | 7.08e-06 | 1.67e-05 |
| **Vitamin and mineral supplements: Multivitamins +/- minerals** | 0.0673 | 0.0325 | 0.039 | 0.056 |
| **Arm fat mass (right)** | 0.2853 | 0.0185 | 8.24e-54 | 3.63e-52 |
| **Forced expiratory volume in 1-second (FEV1)** | -0.0303 | 0.0231 | 0.190 | 0.237 |
| **Mouth/teeth dental problems: Toothache** | 0.211 | 0.0575 | 2,00e-04 | 0.0004 |
| **Ratio of docosahexaenoic acid to total fatty acids** | -0.2177 | 0.0301 | 4.79e-13 | 1.87e-12 |
| **Phospholipids to total lipids ratio in small HDL** | 0.044 | 0.0375 | 0.240 | 0.296 |
| **Vitamin and mineral supplements: None of the above** | -0.0975 | 0.0316 | 0.002 | 0.004 |
| **HDL cholesterol** | -0.196 | 0.0269 | 3.27e-13 | 1.34e-12 |
| **Potassium in urine** | 0.0519 | 0.0277 | 0.061 | 0.086 |
| **Total lipids in very large VLDL** | 0.1825 | 0.0292 | 4.28e-10 | 1.38e-09 |
| **Pulse wave peak to peak time** | -0.2639 | 0.0432 | 9.88e-10 | 3.09e-09 |
| **Amyotrophic lateral sclerosis** | 0.0347 | 0.0595 | 0.561 | 0.604 |
| **LDL cholesterol** | -0.0631 | 0.0385 | 0.102 | 0.137 |
| **Types of transport used (excluding work): Cycle** | -0.2673 | 0.0309 | 5.09e-18 | 2.80e-17 |
| **Caudate volume** | -0.0089 | 0.0537 | 0.868 | 0.882 |
| **Type 2 diabetes** | 0.2798 | 0.0272 | 7.37e-25 | 6.37e-24 |
| **Exposure to tobacco smoke at home** | 0.2887 | 0.0393 | 2.02e-13 | 8.40e-13 |
| **Age at menopause (last menstrual period)** | -0.1986 | 0.0273 | 3.51e-13 | 1.42e-12 |
| **Intelligence** | -0.2842 | 0.0225 | 1.82e-36 | 2.86e-35 |
| **Alcohol intake versus 10 years previously** | 0.2551 | 0.0303 | 3.98e-17 | 2.12e-16 |
| **Triglycerides in HDL** | 0.1666 | 0.0359 | 3.56e-06 | 8.44e-06 |
| **Frequency of depressed mood in last 2 weeks** | 0.4376 | 0.0306 | 2.87e-46 | 7.04e-45 |
| **Total cholesterol** | 0.0404 | 0.0351 | 0.250 | 0.304 |
| **Putamen volume** | 0.0217 | 0.0536 | 0.686 | 0.717 |
| **Phospholipids in small HDL** | 0.1038 | 0.0324 | 0.001 | 0.002 |
| **Ratio of polyunsaturated fatty acids to monounsaturated fatty acids** | -0.2552 | 0.0292 | 2.23e-18 | 1.24e-17 |
| **Mouth/teeth dental problems: Dentures** | 0.2876 | 0.0257 | 3.63e-29 | 3.72e-28 |
| **Glycoprotein acetyls** | 0.2151 | 0.032 | 1.80e-11 | 6.56e-11 |
| **Neuroticism score** | 0.2814 | 0.0274 | 1.01e-24 | 8.56e-24 |
| **ICD10: H25 Senile cataract** | 0.1413 | 0.0939 | 0.132 | 0.173 |
| **Phospholipids in medium VLDL** | 0.1862 | 0.0647 | 0.004 | 0.007 |
| **Ever had stillbirth, spontaneous miscarriage or termination** | 0.2722 | 0.0465 | 4.77e-09 | 1.37e-08 |
| **Concentration of very large HDL particles** | -0.231 | 0.0305 | 3.53e-14 | 1.57e-13 |
| **Illness, injury, bereavement, stress in last 2 years: Serious illness, injury or assault of a close relative** | 0.0995 | 0.0454 | 0.028 | 0.043 |
| **Medication for pain relief, constipation, heartburn: Paracetamol** | 0.4428 | 0.0261 | 1.03e-64 | 6.48e-63 |
| **Hearing difficulty/problems with background noise** | 0.1528 | 0.0249 | 8.80e-10 | 2.78e-09 |
| **Mineral and other dietary supplements: Glucosamine** | 0.1585 | 0.0347 | 5.01e-06 | 1.18e-05 |
| **Triglycerides to total lipids ratio in very small VLDL** | 0.2221 | 0.0357 | 5.25e-10 | 1.68e-09 |
| **Nervous feelings** | 0.0636 | 0.0276 | 0.021 | 0.033 |
| **Concentration of IDL particles** | 0.0974 | 0.087 | 0.263 | 0.317 |
| **Ever unenthusiastic/disinterested for a whole week** | 0.3233 | 0.0402 | 8.46e-16 | 4.19e-15 |
| **ICD10: M24 Other specific joint derangements** | 0.2972 | 0.1113 | 0.008 | 0.013 |
| **Bring up phlegm/sputum/mucus on most days** | 0.2986 | 0.0634 | 2.46e-06 | 5.94e-06 |
| **Fractured bone site(s): Ankle** | 0.2058 | 0.0658 | 0.002 | 0.003 |
| **Arm fat percentage (right)** | 0.2371 | 0.0196 | 1.26e-33 | 1.73e-32 |
| **Alcohol intake frequency.** | 0.252 | 0.0217 | 4.51e-31 | 5.10e-30 |
| **Femoral neck bone mineral density** | 0.0675 | 0.0414 | 0.103 | 0.138 |
| **Coronary artery disease** | 0.3524 | 0.0275 | 1.22e-37 | 2.08e-36 |
| **Uric acid** | 0.0297 | 0.0285 | 0.297 | 0.349 |
| **Total cholesterol in HDL** | -0.1639 | 0.0787 | 0.037 | 0.055 |
| **Tinnitus: No, never** | -0.2225 | 0.0428 | 1.96e-07 | 5.15e-07 |
| **Triglycerides in very small VLDL** | 0.15 | 0.0645 | 0.020 | 0.032 |
| **Total lipids in medium HDL** | -0.0803 | 0.0318 | 0.012 | 0.019 |
| **Serum total cholesterol** | 0.0381 | 0.0841 | 0.650 | 0.688 |
| **ICD10: N81 Female genital prolapse** | 0.2338 | 0.0674 | 5,00e-04 | 0.001 |
| **Exposure to tobacco smoke outside home** | 0.2756 | 0.0309 | 5.41e-19 | 3.18e-18 |
| **Qualifications: CSEs or equivalent** | 0.3432 | 0.038 | 1.57e-19 | 9.61e-19 |
| **Concentration of very small VLDL particles** | 0.0502 | 0.0367 | 0.172 | 0.218 |
| **Age at menarche** | -0.1157 | 0.0265 | 1.26e-05 | 2.85e-05 |
| **Free cholesterol to total lipids ratio in medium LDL** | -0.2212 | 0.0321 | 5.92e-12 | 2.18e-11 |

| | | | | |
|---|---|---|---|---|
| **Time spent watching television (TV)** | 0.3004 | 0.0221 | 3.34e-42 | 6.69e-41 |
| **Used an inhaler for chest within last hour** | 0.1389 | 0.0759 | 0.067 | 0.093 |
| **ICD10: Z80 Family history of malignant neoplasm** | -0.0155 | 0.151 | 0.918 | 0.927 |
| **Depressive symptoms** | 0.4246 | 0.0399 | 1.95e-26 | 1.72e-25 |
| **Current tobacco smoking** | 0.3303 | 0.026 | 5.87e-37 | 9.59e-36 |
| **Medication for cholesterol, blood pressure, diabetes, or take exogenous hormones: Insulin** | 0.1103 | 0.0747 | 0.140 | 0.182 |
| **Childhood intelligence** | -0.2505 | 0.0591 | 2.24e-05 | 4.97e-05 |
| **Phospholipids in very small VLDL** | 0.1015 | 0.0779 | 0.193 | 0.240 |
| **VLDL cholesterol** | 0.0973 | 0.0337 | 0.004 | 0.007 |
| **College completion** | -0.421 | 0.0351 | 3.38e-33 | 4.26e-32 |
| **Phospholipids to total lipids ratio in large HDL** | 0.2666 | 0.0303 | 1.30e-18 | 7.53e-18 |
| **Concentration of chylomicrons and extremely large VLDL particles** | 0.1937 | 0.0308 | 3.02e-10 | 9.97e-10 |
| **Total lipids in small VLDL** | 0.1292 | 0.0294 | 1.08e-05 | 2.50e-05 |
| **ICD10: N20 Calculus of kidney and ureter** | 0.1657 | 0.0616 | 0.007 | 0.012 |
| **Cerebral aneurysm** | 0.175 | 0.0611 | 0.004 | 0.007 |
| **Lung cancer** | 0.0073 | 0.1042 | 0.944 | 0.946 |
| **ICD10: D25 Leiomyoma of uterus** | 0.1256 | 0.0546 | 0.021 | 0.034 |
| **Type of tobacco previously smoked: Cigars or pipes** | -0.0848 | 0.0939 | 0.367 | 0.420 |
| **Number of children fathered** | 0.331 | 0.0342 | 3.78e-22 | 2.73e-21 |
| **ICD10: I20 Angina pectoris** | 0.4789 | 0.0739 | 9.14e-11 | 3.20e-10 |
| **Average weekly intake of other alcoholic drinks** | 0.4461 | 0.4033 | 0.269 | 0.321 |
| **Major depressive disorder (ICD-10 coded)** | 0.4871 | 0.0487 | 1.66e-23 | 1.33e-22 |
| **Total lipids in small HDL** | 0.102 | 0.0318 | 0.001 | 0.002 |
| **Diagnoses - main ICD10: I10 Essential (primary) hypertension** | 0.2647 | 0.119 | 0.026 | 0.040 |
| **Triglycerides to total lipids ratio in small VLDL** | 0.1891 | 0.0432 | 1.21e-05 | 2.77e-05 |
| **ICD10: M67 Other disorders of synovium and tendon** | 0.4785 | 0.1888 | 0.011 | 0.018 |
| **Neuroticism** | 0.2685 | 0.028 | 9.15e-22 | 6.40e-21 |
| **Concentration of small HDL particles** | 0.2154 | 0.1074 | 0.045 | 0.064 |
| **Total phospholipids in lipoprotein particles** | -0.0381 | 0.0293 | 0.194 | 0.240 |
| **Medication for pain relief, constipation, heartburn: Aspirin** | 0.3256 | 0.0326 | 1.63e-23 | 1.33e-22 |
| **Cholesterol in very small VLDL** | -0.0563 | 0.0364 | 0.122 | 0.161 |
| **Back pain for 3+ months** | 0.593 | 0.0537 | 2.56e-28 | 2.51e-27 |
| **ICD10: R14 Flatulence and related conditions** | 0.1918 | 0.1 | 0.055 | 0.0781 |
| **Mineral and other dietary supplements: Iron** | 0.0112 | 0.0708 | 0.875 | 0.887 |
| **Phospholipids to total lipids ratio in small LDL** | -0.1303 | 0.0416 | 0.002 | 0.003 |
| **Cholesteryl esters to total lipids ratio in chylomicrons and extremely large VLDL** | -0.0812 | 0.0435 | 0.062 | 0.087 |
| **Forced vital capacity (FVC), Best measure** | -0.0263 | 0.0234 | 0.260 | 0.315 |
| **Cholesterol in large HDL** | -0.229 | 0.0268 | 1.34e-17 | 7.29e-17 |
| **Reason for glasses/contact lenses: Other eye condition** | 0.0854 | 0.1408 | 0.544 | 0.594 |
| **Fractured bone site(s): Arm** | 0.0839 | 0.0964 | 0.384 | 0.436 |
| **Impedance of arm (left)** | -0.2512 | 0.0187 | 5.43e-41 | 1.04e-39 |
| **Neo-conscientiousness** | 0.1257 | 0.091 | 0.167 | 0.213 |
| **Triglycerides in medium HDL** | 0.1776 | 0.0364 | 1.09e-06 | 2.71e-06 |
| **Extreme waist-to-hip ratio** | 0.2462 | 0.069 | 4,00e-04 | 0.001 |
| **Remnant cholesterol (non-HDL, non-LDL -cholesterol)** | -0.0112 | 0.0358 | 0.755 | 0.783 |
| **Duration of vigorous activity** | 0.2167 | 0.0358 | 1.40e-09 | 4.36e-09 |
| **Why reduced smoking: None of the above** | 0.101 | 0.1868 | 0.589 | 0.633 |
| **Types of physical activity in last 4 weeks: None of the above** | 0.3574 | 0.0316 | 1.05e-29 | 1.10e-28 |
| **Free cholesterol** | 0.0277 | 0.1009 | 0.78 | 0.810 |
| **Qualifications: O levels/GCSEs or equivalent** | -0.3493 | 0.029 | 2.58e-33 | 3.45e-32 |
| **Concentration of VLDL particles** | 0.1177 | 0.0312 | 2,00e-04 | 0.0004 |
| **Mineral and other dietary supplements: Selenium** | -0.0319 | 0.0641 | 0.618 | 0.662 |
| **Total lipids in IDL** | 0.0884 | 0.0868 | 0.309 | 0.362 |
| **ICD10: D12 Benign neoplasm of colon, rectum, anus and anal canal** | 0.1557 | 0.0509 | 0.002 | 0.004 |
| **Albumin** | -0.0416 | 0.088 | 0.637 | 0.678 |
| **Degree of unsaturation** | -0.234 | 0.0316 | 1.26e-13 | 5.28e-13 |
| **Total cholesterol in IDL** | 0.0621 | 0.0853 | 0.467 | 0.518 |
| **Eczema** | 0.0677 | 0.0588 | 0.250 | 0.304 |
| **HOMA-B** | 0.1435 | 0.0553 | 0.009 | 0.016 |
| **Free cholesterol in large LDL** | -0.1561 | 0.0391 | 6.65e-05 | 0.0001 |
| **Triglycerides to total lipids ratio in IDL** | 0.2455 | 0.0316 | 8.39e-15 | 3.89e-14 |
| **Arm fat-free mass (left)** | 0.2766 | 0.0179 | 1.02e-53 | 4.08e-52 |
| **ICD10: I30 Acute pericarditis** | 0.1413 | 0.1068 | 0.186 | 0.234 |
| **Cholesteryl esters in HDL** | -0.1997 | 0.0268 | 9.94e-14 | 4.21e-13 |
| **Cholesterol to total lipids ratio in medium HDL** | -0.235 | 0.029 | 5.98e-16 | 3.00e-15 |
| **Average diameter for LDL particles** | -0.1567 | 0.0391 | 6.20e-05 | 0.0001 |
| **Fractured bone site(s): Leg** | 0.2695 | 0.131 | 0.040 | 0.058 |
| **Number of days/week of moderate physical activity 10+ minutes** | 0.0744 | 0.0278 | 0.008 | 0.013 |
| **Worry too long after embarrassment** | 0.0172 | 0.0287 | 0.548 | 0.597 |
| **ICD10: K21 Gastro-oesophageal reflux disease** | 0.5161 | 0.0737 | 2.50e-12 | 9.35e-12 |

| | | | | |
|---|---|---|---|---|
| **Creatinine (enzymatic) in urine** | 0.188 | 0.0242 | 8.85e-15 | 4.07e-14 |
| **Phospholipids in large HDL** | -0.2048 | 0.0274 | 8.01e-14 | 3.46e-13 |
| **Distance between home and job workplace** | 0.0751 | 0.0588 | 0.202 | 0.250 |
| **Total lipids in very small VLDL** | 0.0551 | 0.0343 | 0.108 | 0.144 |
| **Schizophrenia** | -0.0257 | 0.0234 | 0.272 | 0.324 |
| **Vitamin and mineral supplements: Vitamin C** | 0.0946 | 0.0444 | 0.033 | 0.049 |
| **Cholesteryl esters to total lipids ratio in large HDL** | -0.2611 | 0.0291 | 3.13e-19 | 1.87e-18 |
| **Frequency of stair climbing in last 4 weeks** | -0.1768 | 0.0281 | 3.03e-10 | 9.97e-10 |
| **Body fat percentage** | 0.2643 | 0.0205 | 3.44e-38 | 6.08e-37 |
| **Trunk fat-free mass** | 0.2183 | 0.0182 | 4.54e-33 | 5.56e-32 |
| **Pallidum volume** | 0.0427 | 0.0682 | 0.532 | 0.583 |
| **3-hydroxybutyrate** | -0.3182 | 0.1401 | 0.023 | 0.0362 |
| **Townsend deprivation index at recruitment** | 0.1887 | 0.029 | 7.63e-11 | 2.7e-10 |
| **Diagnoses - main ICD10: H40 Glaucoma** | 0.0333 | 0.0855 | 0.697 | 0.726 |
| **Cholesteryl esters in large LDL** | -0.0819 | 0.0377 | 0.030 | 0.045 |
| **Birth weight** | -0.04 | 0.0224 | 0.074 | 0.102 |
| **Concentration of small LDL particles** | 0.0191 | 0.0373 | 0.608 | 0.652 |
| **Description of average fatty acid chain length, not actual carbon number** | -0.3657 | 0.0922 | 7.26e-05 | 0.0002 |
| **Cigarettes smoked per day** | 0.2673 | 0.0287 | 1.15e-20 | 7.50e-20 |
| **Total lipids in very small VLDL** | 0.1195 | 0.0721 | 0.097 | 0.131 |
| **Number of full brothers** | 0.2143 | 0.041 | 1.73e-07 | 4.59e-07 |
| **Age started oral contraceptive pill** | -0.4247 | 0.0388 | 7.70e-28 | 7.38e-27 |
| **Infant head circumference** | -0.0769 | 0.0586 | 0.189 | 0.237 |
| **Trunk fat percentage** | 0.2403 | 0.0203 | 3.39e-32 | 3.93e-31 |
| **Cigarettes per Day** | 0.2673 | 0.0287 | 1.28e-20 | 8.19e-20 |
| **Total cholesterol in large LDL** | 0.0474 | 0.0799 | 0.553 | 0.599 |
| **Vascular/heart problems diagnosed by doctor: Heart attack** | 0.3308 | 0.0389 | 1.83e-17 | 9.82e-17 |
| **Mean time to correctly identify matches** | -0.0933 | 0.0223 | 2.85e-05 | 6.28e-05 |
| **Pain type(s) experienced in last month: Neck or shoulder pain** | 0.5836 | 0.0279 | 3.90e-97 | 8.59e-95 |
| **Qualifications: College or University degree** | -0.4122 | 0.0204 | 1.90e-90 | 2.10e-88 |
| **Pain type(s) experienced in last month: Facial pain** | 0.4303 | 0.0688 | 3.90e-10 | 1.27e-09 |
| **Ischemic stroke** | 0.1525 | 0.0386 | 7.86e-05 | 0.0002 |
| **Fractured bone site(s): Spine** | 0.1102 | 0.1684 | 0.513 | 0.566 |
| **Phospholipids to total lipids ratio in chylomicrons and extremely large VLDL** | 0.1985 | 0.0464 | 1.92e-05 | 4.33e-05 |
| **Total lipids in large HDL** | -0.1731 | 0.0673 | 0.010 | 0.002 |
| **Free cholesterol in HDL** | -0.1787 | 0.0278 | 1.36e-10 | 4.64e-10 |
| **Cholesteryl esters to total lipids ratio in medium HDL** | -0.2211 | 0.0297 | 9.50e-14 | 4.07e-13 |
| **Transport type for commuting to job workplace: Cycle** | -0.3292 | 0.0331 | 2.69e-23 | 2.08e-22 |
| **Other polyunsaturated fatty acids than 18:2** | -0.0161 | 0.0676 | 0.812 | 0.83 |
| **Medication for cholesterol, blood pressure, diabetes, or take exogenous hormones: None of the above** | -0.3183 | 0.0316 | 8.20e-24 | 6.82e-23 |
| **Hypermetropia** | 0.0903 | 0.0568 | 0.112 | 0.148 |
| **Triglycerides to total lipids ratio in small HDL** | 0.2086 | 0.0286 | 2.90e-13 | 1.195e-12 |
| **Lung adenocarcinoma** | 0.1893 | 0.1289 | 0.142 | 0.184 |
| **Total lipids in medium VLDL** | 0.1018 | 0.0327 | 0.002 | 0.003 |
| **ICD10: I84 Haemorrhoids** | 0.2656 | 0.0608 | 1.25e-05 | 2.845e-05 |
| **Triglycerides in large VLDL** | 0.2253 | 0.0623 | 3,00e-04 | 0.0006 |
| **Cholesterol in very large VLDL** | 0.1624 | 0.0295 | 3.63e-08 | 9.99e-08 |
| **ICD10: K62 Other diseases of anus and rectum** | 0.4799 | 0.095 | 4.37e-07 | 1.12e-06 |
| **Total cholesterol in medium LDL** | 0.0483 | 0.0769 | 0.530 | 0.582 |
| **Cholesteryl esters to total lipids ratio in large LDL** | -0.0734 | 0.0418 | 0.079 | 0.109 |
| **Concentration of very large VLDL particles** | 0.1841 | 0.029 | 2.18e-10 | 7.40e-10 |
| **Phospholipids in VLDL** | 0.1425 | 0.0298 | 1.74e-06 | 4.28e-06 |
| **Falls in the last year** | 0.4066 | 0.0276 | 4.68e-49 | 1.38e-47 |
| **Creatinine** | 0.024 | 0.0572 | 0.675 | 0.708 |
| **ICD10: M17 Gonarthrosis [arthrosis of knee]** | 0.3664 | 0.0554 | 3.73e-11 | 1.34e-10 |
| **Transport type for commuting to job workplace: Car/motor vehicle** | 0.4238 | 0.0447 | 2.58e-21 | 1.78e-20 |
| **Phospholipids in large HDL** | -0.1595 | 0.0686 | 0.020 | 0.032 |
| **Leg fat mass (right)** | 0.3255 | 0.0191 | 6.60e-65 | 4.85e-63 |
| **Diastolic blood pressure** | 0.0182 | 0.0192 | 0.343 | 0.398 |
| **Cholesteryl esters to total lipids ratio in small VLDL** | -0.1084 | 0.0503 | 0.031 | 0.047 |
| **Free cholesterol in medium LDL** | -0.0999 | 0.0414 | 0.016 | 0.025 |
| **Why reduced smoking: Illness or ill health** | 0.4215 | 0.1912 | 0.028 | 0.042 |
| **Headaches for 3+ months** | 0.1647 | 0.0444 | 2,00e-04 | 0.0004 |
| **Alcohol drinker status: Never** | 0.0977 | 0.0484 | 0.044 | 0.063 |
| **Total fatty acids** | 0.1124 | 0.0316 | 4,00e-04 | 0.001 |
| **Cholesteryl esters in large VLDL** | 0.1323 | 0.0301 | 1.13e-05 | 2.59e-05 |
| **Happiness** | 0.0322 | 0.0341 | 0.346 | 0.400 |
| **Total triglycerides** | 0.1758 | 0.0298 | 3.90e-09 | 1.15e-08 |

| | | | | |
|---|---|---|---|---|
| **Number of treatments/medications taken** | 0.5169 | 0.024 | 5.15e-103 | 2.27e-100 |
| **Total lipids in small VLDL** | 0.1449 | 0.0611 | 0.018 | 0.029 |
| **ICD10: R35 Polyuria** | 0.28 | 0.1117 | 0.012 | 0.020 |
| **Miserableness** | 0.3272 | 0.0275 | 1.15e-32 | 1.37e-31 |
| **Average diameter for HDL particles** | -0.2183 | 0.028 | 6.32e-15 | 3.00e-14 |
| **Length of working week for main job** | 0.1907 | 0.0399 | 1.77e-06 | 4.33e-06 |
| **Target heart rate achieved** | -0.1415 | 0.0661 | 0.032 | 0.048 |
| **Concentration of medium LDL particles** | 0.0873 | 0.0804 | 0.277 | 0.328 |
| **Anorexia Nervosa** | -0.0923 | 0.0495 | 0.062 | 0.087 |
| **ICD10: K35 Acute appendicitis** | 0.054 | 0.1178 | 0.647 | 0.687 |
| **Job involves mainly walking or standing** | 0.3033 | 0.0248 | 2.15e-34 | 3.07e-33 |
| **Vitamin and mineral supplements: Vitamin A** | 0.1519 | 0.0723 | 0.036 | 0.053 |
| **Sleeplessness / insomnia** | 0.3537 | 0.023 | 1.88e-53 | 6.93e-52 |
| **ICD10: K44 Diaphragmatic hernia** | 0.2471 | 0.07 | 4,00e-04 | 0.001 |
| **Citrate** | -0.2618 | 0.0817 | 0.001 | 0.002 |
| **Glutamine** | -0.0563 | 0.0275 | 0.040 | 0.058 |
| **Tense / highly strung** | 0.2372 | 0.0293 | 5.33e-16 | 2.70e-15 |
| **ICD10: N40 Hyperplasia of prostate** | 0.1276 | 0.0587 | 0.030 | 0.045 |
| **Cholesteryl esters in very large HDL** | -0.2401 | 0.03 | 1.15e-15 | 5.57e-15 |
| **Wheeze or whistling in the chest in last year** | 0.3898 | 0.0276 | 3.72e-45 | 7.81e-44 |
| **Free cholesterol in very small VLDL** | 0.027 | 0.0359 | 0.453 | 0.507 |
| **Alcohol usually taken with meals** | -0.2563 | 0.0261 | 1.06e-22 | 8.06e-22 |
| **Triglycerides in small VLDL** | 0.1636 | 0.0303 | 6.36e-08 | 1.73e-07 |
| **ICD10: S52 Fracture of forearm** | -0.053 | 0.0597 | 0.375 | 0.427 |
| **Ever stopped smoking for 6+ months** | 0.3508 | 0.0745 | 2.47e-06 | 5.94e-06 |
| **Handedness (chirality/laterality): Use both right and left hands equally** | 0.267 | 0.0818 | 0.001 | 0.002 |
| **Cataract** | 0.028 | 0.0723 | 0.670 | 0.727 |
| **ICD10: J33 Nasal polyp** | 0.104 | 0.0924 | 0.260 | 0.315 |
| **Daytime dozing / sleeping (narcolepsy)** | 0.1069 | 0.025 | 1.97e-05 | 4.34e-05 |
| **Adopted as a child** | 0.3046 | 0.0625 | 1.08e-06 | 2.70e-06 |
| **Ibuprofen (e.g. Nurofen)** | 0.3702 | 0.0325 | 4.17e-30 | 4.48e-29 |
| **ICD10: K40 Inguinal hernia** | 0.0183 | 0.0437 | 0.676 | 0.708 |
| **Doctor diagnosed hayfever or allergic rhinitis** | 0.1023 | 0.0338 | 0.003 | 0.004 |
| **Omega-6 fatty acids** | 0.0842 | 0.1083 | 0.437 | 0.493 |
| **Number of depression episodes** | 0.0902 | 0.0739 | 0.222 | 0.274 |
| **Waist-to-hip ratio** | 0.2173 | 0.0299 | 3.73e-13 | 1.49e-12 |
| **Leg fat-free mass (right)** | 0.2175 | 0.0177 | 9.05e-35 | 1.38e-33 |
| **Omega-3 fatty acids** | -0.0272 | 0.0277 | 0.325 | 0.379 |
| **Illness, injury, bereavement, stress in last 2 years: Death of a close relative** | 0.3867 | 0.0543 | 1.07e-12 | 4.09e-12 |
| **Illness, injury, bereavement, stress in last 2 years: Marital separation/divorce** | 0.2884 | 0.0843 | 6,00e-04 | 0.001 |
| **HbA1c** | 0.0666 | 0.0396 | 0.093 | 0.126 |
| **Chronic kidney disease** | 0.083 | 0.07 | 0.235 | 0.290 |
| **Hand grip strength (right)** | -0.0148 | 0.0217 | 0.497 | 0.550 |
| **Vitamin and mineral supplements: Folic acid or Folate (Vit B9)** | 0.0813 | 0.0903 | 0.368 | 0.420 |
| **Cholesterol lowering medication** | 0.3597 | 0.0382 | 5.23e-21 | 3.55e-20 |
| **Free cholesterol to total lipids ratio in small LDL** | -0.2124 | 0.0362 | 4.28e-09 | 1.26e-08 |
| **Mean diameter for VLDL particles** | 0.1708 | 0.0602 | 0.005 | 0.008 |
| **Handedness (chirality/laterality): Left-handed** | -0.1362 | 0.0513 | 0.008 | 0.014 |
| **Free cholesterol to total lipids ratio in medium HDL** | -0.219 | 0.029 | 4.53e-14 | 2.00e-13 |
| **Reason for reducing amount of alcohol drunk: Illness or ill health** | 0.5039 | 0.108 | 3.06e-06 | 7.338e-06 |
| **ICD10: J84.1 Other interstitial pulmonary diseases with fibrosis** | -0.2923 | 0.1612 | 0.070 | 0.097 |
| **Total cholesterol in very large HDL** | -0.2394 | 0.1379 | 0.083 | 0.113 |
| **Primary biliary cirrhosis** | 0.0066 | 0.084 | 0.937 | 0.941 |
| **Smoking behaviors : Smoking cessation** | -0.1275 | 0.0497 | 0.010 | 0.017 |
| **Phospholipids to total lipids ratio in very large HDL** | -0.1958 | 0.0324 | 1.44e-09 | 4.43e-09 |
| **Number of older siblings** | 0.2647 | 0.0719 | 2,00e-04 | 0.0004 |
| **ICD10: I48 Atrial fibrillation and flutter** | 0.118 | 0.0441 | 0.007 | 0.013 |
| **Pain type(s) experienced in last month: Knee pain** | 0.4649 | 0.0304 | 6.10e-53 | 2.07e-51 |
| **Oral contraceptive pill or minipill** | -0.0202 | 0.0888 | 0.821 | 0.840 |
| **Apolipoprotein A-I** | -0.0981 | 0.0881 | 0.266 | 0.319 |
| **Total cholesterol in medium HDL** | -0.0241 | 0.0947 | 0.799 | 0.823 |
| **Total lipids in medium LDL** | 0.0361 | 0.0387 | 0.351 | 0.404 |
| **Mean platelet volume** | -0.0185 | 0.0202 | 0.360 | 0.414 |
| **Concentration of large HDL particles** | -0.1643 | 0.0672 | 0.014 | 0.023 |
| **ICD10: R10 Abdominal and pelvic pain** | 0.5573 | 0.0503 | 1.76e-28 | 1.77e-27 |
| **Emphysema/chronic bronchitis** | 0.3978 | 0.0455 | 2.10e-18 | 1.19e-17 |
| **Impedance of whole body** | -0.2005 | 0.0188 | 1.81e-26 | 1.63e-25 |
| **Blood pressure medication** | 0.1593 | 0.0281 | 1.49e-08 | 4.18e-08 |
| **Sodium in urine** | 0.2126 | 0.0233 | 7.98e-20 | 4.96e-19 |
| **Free cholesterol to total lipids ratio in medium VLDL** | -0.1967 | 0.0379 | 2.11e-07 | 5.52e-07 |

| | | | | |
|---|---|---|---|---|
| **Phosphatidylcholines** | -0.0525 | 0.028 | 0.061 | 0.086 |
| **Alanine** | 0.113 | 0.0642 | 0.079 | 0.108 |
| **Acetoacetate** | -0.1376 | 0.0893 | 0.123 | 0.162 |
| **Ever had prostate specific antigen (PSA) test** | 0.0649 | 0.0405 | 0.109 | 0.145 |
| **Time from waking to first cigarette** | -0.2771 | 0.0559 | 7.32e-07 | 1.86e-06 |
| **Cholesteryl esters in small HDL** | 0.0452 | 0.0323 | 0.162 | 0.207 |
| **Concentration of very large HDL particles** | -0.191 | 0.0853 | 0.025 | 0.039 |
| **Ratio of triglycerides to phosphoglycerides** | 0.2012 | 0.0278 | 4.37e-13 | 1.725e-12 |
| **Concentration of very large VLDL particles** | 0.2131 | 0.0605 | 4,00e-04 | 0.0008 |
| **Total lipids in very large HDL** | -0.2292 | 0.0305 | 5.96e-14 | 2.60e-13 |
| **Forced expiratory volume in 1-second (FEV1), predicted** | 0.0889 | 0.0247 | 3,00e-04 | 0.0006 |
| **circulating leptin levels** | 0.1201 | 0.0541 | 0.026 | 0.041 |
| **Mean diameter for HDL particles** | -0.1843 | 0.0658 | 0.005 | 0.009 |
| **Concentration of very small VLDL particles** | 0.1241 | 0.0717 | 0.084 | 0.114 |
| **ICD10: F43 Reaction to severe stress and adjustment disorders** | -0.0352 | 0.1809 | 0.846 | 0.863 |
| **Total fatty acids** | 0.1692 | 0.1033 | 0.102 | 0.137 |
| **Morning/evening person (chronotype)** | -0.0531 | 0.0202 | 0.009 | 0.014 |
| **Triglycerides in very large HDL** | 0.0526 | 0.0709 | 0.458 | 0.510 |
| **Apolipoprotein B** | 0.1094 | 0.0741 | 0.140 | 0.182 |
| **Number of operations, self-reported** | 0.5592 | 0.027 | 2.11e-95 | 3.10e-93 |
| **Total lipids in very large VLDL** | 0.1908 | 0.0557 | 6,00e-04 | 0.001 |
| **Frequency of unenthusiasm / disinterest in last 2 weeks** | 0.43 | 0.0283 | 3.33e-52 | 1.05e-50 |
| **Fracture resulting from simple fall** | 0.0118 | 0.0633 | 0.852 | 0.867 |
| **Operation code: bilateral oophorectomy** | 0.3928 | 0.0509 | 1.21e-14 | 5.50e-14 |
| **Total lipids in VLDL** | 0.1529 | 0.0294 | 1.92e-07 | 5.07e-07 |
| **apolipoprotein B** | 0.0312 | 0.0238 | 0.189 | 0.237 |
| **ICD10: N32 Other disorders of bladder** | 0.1215 | 0.0764 | 0.112 | 0.148 |
| **Acetoacetate** | 0.0696 | 0.048 | 0.148 | 0.190 |
| **Average weekly champagne plus white wine intake** | -0.2512 | 0.0356 | 1.77e-12 | 6.66e-12 |
| **Number of cigarettes currently smoked daily (current cigarette smokers)** | 0.2239 | 0.055 | 4.65e-05 | 0.0001 |
| **Mouth/teeth dental problems: Mouth ulcers** | 0.1181 | 0.0325 | 3,00e-04 | 0.0006 |
| **Weight change compared with 1 year ago** | 0.2642 | 0.0437 | 1.53e-09 | 4.66e-09 |
| **Total lipids in large VLDL** | 0.1654 | 0.0297 | 2.54e-08 | 7.06e-08 |
| **ICD10: O75 Other complications of labour and delivery not elsewhere classified** | -0.0669 | 0.0894 | 0.454 | 0.507 |
| **ICD10: R11 Nausea and vomiting** | 0.5524 | 0.2068 | 0.008 | 0.0129 |
| **ICD10: M54 Dorsalgia** | 0.8303 | 0.0576 | 4.75e-47 | 1.23e-45 |
| **Total lipids in large VLDL** | 0.1911 | 0.0573 | 9,00e-04 | 0.002 |
| **Blood clot in the leg (DVT)** | 0.3503 | 0.0564 | 5.31e-10 | 1.68e-09 |
| **Triglycerides in VLDL** | 0.1722 | 0.0299 | 8.25e-09 | 2.35e-08 |
| **Cholesteryl esters in small LDL** | 0.0379 | 0.0374 | 0.310 | 0.362 |
| **Cholesterol to total lipids ratio in very large HDL** | 0.0882 | 0.0428 | 0.039 | 0.057 |
| **Maximum workload during fitness test** | -0.2889 | 0.0543 | 1.06e-07 | 2.86e-07 |
| **Phospholipids to total lipids ratio in large LDL** | 0.0093 | 0.0391 | 0.812 | 0.833 |
| **Total lipids in lipoprotein particles** | 0.0156 | 0.0322 | 0.628 | 0.670 |
| **Guilty feelings** | 0.1544 | 0.0284 | 5.69e-08 | 1.56e-07 |
| **Free cholesterol to total lipids ratio in chylomicrons and extremely large VLDL** | -0.0965 | 0.0385 | 0.012 | 0.020 |
| **Mineral and other dietary supplements: Zinc** | 0.0615 | 0.0459 | 0.180 | 0.227 |
| **Average number of methylene groups in a fatty acid chain** | 0.0817 | 0.106 | 0.440 | 0.495 |
| **Free cholesterol in chylomicrons and extremely large VLDL** | 0.1941 | 0.0323 | 1.90e-09 | 5.72e-09 |
| **ICD10: H26.9 Cataract, unspecified** | 0.0424 | 0.0512 | 0.408 | 0.461 |
| **Had major operations** | 0.4984 | 0.0499 | 1.73e-23 | 1.36e-22 |
| **Cholesterol esters in large VLDL** | 0.057 | 0.0793 | 0.472 | 0.523 |
| **Cholesterol esters in medium HDL** | 0.0035 | 0.0994 | 0.972 | 0.972 |
| **Smoking status: Never** | -0.2845 | 0.0237 | 2.73e-33 | 3.54e-32 |
| **Phospholipids in medium HDL** | 0.0493 | 0.0826 | 0.551 | 0.598 |
| **Work/job satisfaction** | -0.0209 | 0.0459 | 0.649 | 0.688 |
| **Former alcohol drinker** | 0.3015 | 0.056 | 7.32e-08 | 1.98e-07 |
| **Triglycerides in medium VLDL** | 0.1485 | 0.0304 | 1.04e-06 | 2.62e-06 |
| **Fractured/broken bones in last 5 years** | 0.1601 | 0.0375 | 1.95e-05 | 4.34e-05 |
| **ICD10: Z09 Follow-up examination after treatment for conditions other than malignant neoplasms** | 0.2227 | 0.0967 | 0.021 | 0.034 |
| **Cholesteryl esters in medium VLDL** | -0.1228 | 0.0391 | 0.002 | 0.003 |
| **Pain type(s) experienced in last month: Headache** | 0.2681 | 0.0288 | 1.16e-20 | 7.50e-20 |
| **Concentration of HDL particles** | -0.093 | 0.0291 | 0.001 | 0.003 |
| **Mineral and other dietary supplements: Calcium** | -0.063 | 0.0451 | 0.162 | 0.207 |
| **Cholesterol esters in large HDL** | -0.1897 | 0.0673 | 0.005 | 0.008 |
| **Whole body fat mass** | 0.2957 | 0.0188 | 8.46e-56 | 4.15e-54 |
| **Noisy workplace** | 0.3987 | 0.0374 | 1.79e-26 | 1.63e-25 |
| **Forced vital capacity (FVC)** | -0.0319 | 0.0225 | 0.157 | 0.202 |
| **Wears glasses or contact lenses** | 0.0352 | 0.0459 | 0.443 | 0.497 |
| **Apolipoprotein A1** | -0.121 | 0.0296 | 4.30e-05 | 9.34e-05 |

| | | | | |
|---|---|---|---|---|
| **Concentration of large HDL particles** | -0.2251 | 0.0269 | 6.61e-17 | 3.47e-16 |
| **ICD10: J34 Other disorders of nose and nasal sinuses** | 0.7953 | 0.2469 | 0.001 | 0.002 |
| **ICD10: M25 Other joint disorders, not elsewhere classified** | 0.8166 | 0.1295 | 2.87e-10 | 9.60e-10 |
| **Cholesterol esters in medium VLDL** | 0.129 | 0.0583 | 0.027 | 0.041 |
| **Leucine** | 0.1341 | 0.0875 | 0.125 | 0.164 |
| **Ever taken oral contraceptive pill** | 0.0912 | 0.0441 | 0.039 | 0.056 |
| **Ever used hormone-replacement therapy (HRT)** | 0.3886 | 0.0337 | 7.84e-31 | 8.65e-30 |
| **Eye problems/disorders: Injury or trauma resulting in loss of vision** | 0.2413 | 0.1145 | 0.035 | 0.052 |
| **Ever depressed for a whole week** | 0.2286 | 0.0354 | 1.01e-10 | 3.49e-10 |
| **Offspring birth weight** | 0.0456 | 0.0389 | 0.241 | 0.296 |
| **Sensitivity / hurt feelings** | 0.2142 | 0.0273 | 3.86e-15 | 1.852e-14 |
| **ICD10: S76 Injury of muscle and tendon at hip and thigh level** | 0.1343 | 0.1324 | 0.310 | 0.362 |
| **Pulse rate, automated reading** | 0.0992 | 0.0204 | 1.11e-06 | 2.76e-06 |
| **Cholesterol to total lipids ratio in IDL** | -0.2285 | 0.032 | 9.85e-13 | 3.81e-12 |
| **Ratio of linoleic acid to total fatty acids** | -0.2759 | 0.0306 | 1.81e-19 | 1.09e-18 |
| **Phospholipids in medium LDL** | 0.0898 | 0.0798 | 0.260 | 0.315 |
| **Phospholipids in IDL** | 0.0975 | 0.0898 | 0.278 | 0.328 |
| **Myopia** | -0.2752 | 0.0313 | 1.50e-18 | 8.59e-18 |
| **Squamous cell lung cancer** | 0.2079 | 0.0899 | 0.021 | 0.033 |
| **Free cholesterol in large VLDL** | 0.1706 | 0.0291 | 4.42e-09 | 1.28e-08 |
| **Lactate** | 0.0879 | 0.0447 | 0.049 | 0.070 |
| **Phospholipids in medium VLDL** | 0.0741 | 0.0355 | 0.037 | 0.055 |
| **Ever had hysterectomy (womb removed)** | 0.427 | 0.052 | 2.07e-16 | 1.07e-15 |
| **Cholesterol to total lipids ratio in very large VLDL** | -0.2007 | 0.0356 | 1.72e-08 | 4.81e-08 |
| **Alcohol drinker status: Current** | -0.3199 | 0.0417 | 1.70e-14 | 7.66e-14 |
| **Ferritin** | -0.0357 | 0.0795 | 0.654 | 0.690 |
| **Glucose** | 0.1328 | 0.0599 | 0.026 | 0.041 |
| **Blood clot in the lung** | 0.2998 | 0.0677 | 9.36e-06 | 2.17e-05 |
| **Childhood asthma (age<16)** | 0.3331 | 0.0863 | 1,00e-04 | 0.0002 |
| **Triglycerides in large HDL** | 0.02 | 0.0341 | 0.557 | 0.602 |
| **ICD10: E04 Other non-toxic goitre** | 0.1449 | 0.0743 | 0.051 | 0.073 |
| **Frequency of tiredness / lethargy in last 2 weeks** | 0.4347 | 0.0234 | 6.74e-77 | 5.95e-75 |
| **Phospholipids in medium HDL** | -0.0465 | 0.0327 | 0.155 | 0.200 |
| **Trunk fat mass** | 0.2766 | 0.019 | 6.29e-48 | 1.73e-46 |
| **Types of physical activity in last 4 weeks: Other exercises** | -0.2679 | 0.0246 | 1.55e-27 | 1.45e-26 |
| **Total lipids in HDL** | -0.1518 | 0.0289 | 1.45e-07 | 3.88e-07 |
| **Cholesteryl esters to total lipids ratio in IDL** | -0.2228 | 0.0343 | 8.30e-11 | 2.93e-10 |
| **ICD10: I25 Chronic ischaemic heart disease** | 0.3727 | 0.0383 | 2.53e-22 | 1.86e-21 |
| **Ratio of bisallylic groups to total fatty acids** | -0.2104 | 0.0654 | 0.001 | 0.002 |
| **Impedance of arm (right)** | -0.248 | 0.0188 | 6.38e-40 | 1.17e-38 |
| **Concentration of large VLDL particles** | 0.1986 | 0.06 | 9,00e-04 | 0.002 |
| **Phospholipids to total lipids ratio in very large VLDL** | 0.04 | 0.0425 | 0.347 | 0.400 |
| **Ever smoked** | 0.1663 | 0.0241 | 5.51e-12 | 2.04e-11 |
| **ICD10: K20 Oesophagitis** | 0.3954 | 0.0952 | 3.30e-05 | 7.26e-05 |
| **Length of menstrual cycle** | -0.154 | 0.043 | 3,00e-04 | 0.0006 |
| **Triglycerides in small HDL** | 0.2052 | 0.0306 | 2.02e-11 | 7.30e-11 |
| **Polyunsaturated fatty acids** | -0.0122 | 0.0284 | 0.667 | 0.702 |
| **Snoring** | -0.1586 | 0.0224 | 1.41e-12 | 5.37e-12 |
| **Loud music exposure frequency** | 0.1588 | 0.0405 | 8.76e-05 | 0.0002 |
| **Duration of moderate activity** | 0.2584 | 0.0332 | 7.17e-15 | 3.37e-14 |
| **Weight** | 0.2938 | 0.0181 | 5.84e-59 | 3.22e-57 |
| **Phospholipids in very large VLDL** | 0.1815 | 0.0291 | 4.27e-10 | 1.38e-09 |
| **Number of days/week of vigorous physical activity 10+ minutes** | 0.0083 | 0.0279 | 0.767 | 0.794 |
| **Pain type(s) experienced in last month: Stomach or abdominal pain** | 0.427 | 0.0438 | 1.79e-22 | 1.34e-21 |
| **Hippocampus volume** | 0.0587 | 0.0682 | 0.389 | 0.441 |
| **multiple sclerosis** | 0.0729 | 0.0327 | 0.026 | 0.040 |
| **Total cholesterol minus HDL-C** | -0.0407 | 0.0366 | 0.267 | 0.319 |
| **Total lipids in small HDL** | 0.1772 | 0.0952 | 0.063 | 0.087 |
| **Non-accidental death in close genetic family** | 0.0666 | 0.0635 | 0.295 | 0.347 |
| **Heel bone mineral density (BMD) T-score, automated** | 0.0792 | 0.02 | 7.53e-05 | 0.0002 |
| **Diastolic blood pressure, automated reading** | 0.099 | 0.0208 | 2.05e-06 | 5.00e-06 |
| **Triglycerides in small LDL** | 0.1675 | 0.0289 | 6.91e-09 | 1.98e-08 |
| **ICD10: M06.99 Rheumatoid arthritis, unspecified** | 0.2886 | 0.0746 | 1,00e-04 | 0.0002 |
| **Number of pregnancy terminations** | 0.0637 | 0.0471 | 0.176 | 0.223 |
| **Phospholipids to total lipids ratio in small VLDL** | -0.2152 | 0.0374 | 8.72e-09 | 2.47e-08 |
| **Basal metabolic rate** | 0.2523 | 0.0177 | 4.60e-46 | 1.07e-44 |
| **Total lipids in large HDL** | -0.2168 | 0.027 | 1.02e-15 | 4.98e-15 |
| **3-Hydroxybutyrate** | -0.0042 | 0.0459 | 0.927 | 0.934 |
| **Triglycerides in chylomicrons and extremely large VLDL** | 0.1858 | 0.0317 | 4.43e-09 | 1.28e-08 |
| **Mono-unsaturated fatty acids** | 0.2212 | 0.0967 | 0.022 | 0.0346 |

| | | | | |
|---|---|---|---|---|
| **Time spent using computer** | -0.1203 | 0.0237 | 3.84e-07 | 9.91e-07 |
| **Free cholesterol to total lipids ratio in large LDL** | -0.2207 | 0.0304 | 4.02e-13 | 1.60e-12 |
| **Free cholesterol in medium HDL** | -0.1279 | 0.0288 | 9.10e-06 | 2.12e-05 |
| **Ratio of saturated fatty acids to total fatty acids** | 0.1867 | 0.0377 | 7.12e-07 | 1.81e-06 |
| **Subjective well being** | -0.1382 | 0.0416 | 9,00e-04 | 0.002 |
| **Mouth/teeth dental problems: None of the above** | -0.275 | 0.0293 | 6.80e-21 | 4.54e-20 |
| **Average weekly fortified wine intake** | -0.2594 | 0.0506 | 2.89e-07 | 7.50e-07 |
| **Free cholesterol in very large HDL** | -0.2222 | 0.035 | 2.22e-10 | 7.49e-10 |
| **Qualifications: NVQ or HND or HNC or equivalent** | 0.3186 | 0.0388 | 2.17e-16 | 1.11e-15 |
| **Forced expiratory volume in 1-second (FEV1), Best measure** | -0.0261 | 0.0237 | 0.270 | 0.322 |
| **Triglycerides in large LDL** | 0.1767 | 0.0297 | 2.85e-09 | 8.54e-09 |
| **Cholesteryl esters to total lipids ratio in medium VLDL** | -0.2134 | 0.0353 | 1.45e-09 | 4.43e-09 |
| **Difference in height between adolescence and adulthood** | 0.0092 | 0.0597 | 0.877 | 0.887 |
| **Leucine** | 0.1617 | 0.0393 | 3.82e-05 | 8.34e-05 |
| **Valine** | 0.1482 | 0.0363 | 4.51e-05 | 9.74e-05 |
| **Phospholipids in very large VLDL** | 0.2033 | 0.0614 | 9,00e-04 | 0.002 |
| **Concentration of medium HDL particles** | -0.1105 | 0.031 | 4,00e-04 | 0.0008 |
| **Tinnitus: Yes, but not now, but have in the past** | 0.2102 | 0.0678 | 0.002 | 0.003 |
| **Chest pain or discomfort walking normally** | 0.5054 | 0.0523 | 3.98e-22 | 2.83e-21 |
| **Job involves heavy manual or physical work** | 0.355 | 0.025 | 8.41e-46 | 1.85e-44 |
| **Phosphoglycerides** | -0.0175 | 0.0284 | 0.536 | 0.587 |
| **Diagnoses - main ICD10: K30 Dyspepsia** | 0.6359 | 0.1371 | 3.52e-06 | 8.40e-06 |
| **Cholesterol in large LDL** | -0.104 | 0.0371 | 0.005 | 0.001 |

**Table S8** Potentially causal exposures for LDH. Analysis was performed by using TwoSampleMR-database and the phenotype IDs used in the analysis were extracted from the MRC-IEU database. For the analysis, we extracted genetic instruments from FinnGen based GWAS results, to avoid possible bias from overlapping samples. Because MRC-IEU's GWAS data is mainly based on data from UKBB. The results are based on the Inverse variance weighted-model and were statistically significant (P< 0.05). As a sensitivity analysis we also performed analysis by using MR Egger. Nsnp, number of SNPs; OR, odds ratio, pFDR, false discovery rate-corrected p-value; pHET, p-value for heterogeneity; pPLE, p-value for pleiotropy

| Direction | Trait | Method | nsnp | Causal estimate scale | Causal estimate | pFDR | pHET | Egger intercept | pPLE |
|---|---|---|---|---|---|---|---|---|---|
| **Trait-> LDH** | Overweight || id:ieu-a-93<br>Overweight || id:ieu-a-93 | IVW<br>MR Egger | 13<br>13 | OR<br>OR | 1.15 (1.05–1.25)<br>1.14 (0.86–1.52) | 1.72e-03<br>0.45 | 0.035<br>0.023 | 0.0003 | 0.9777 |
| **Trait-> LDH** | Lumbar spine bone mineral density || id:ieu-a-982<br>Lumbar spine bone mineral density || id:ieu-a-982 | IVW<br>MR Egger | 21<br>21 | OR<br>OR | 1.15 (1.08–1.23)<br>1.24 (0.98–1.57) | 2.29e-05<br>0.23 | 0.243<br>0.216 | -0.0054 | 0.5326 |
| **Trait-> LDH** | Higher level of education || id:ukb-b-16489<br>Higher level of education || id:ukb-b-16489 | IVW<br>MR Egger | 227<br>227 | OR<br>OR | 0.34 (0.28–0.42)<br>0.28 (0.12–0.64) | 4.35e-23<br>0.01 | 4.58e-11<br>3.80e-11 | 0.0015 | 0.6122 |

**Table S9** Outcomes that LDH is potentially causal. Analysis was performed by using TwoSampleMR-database and the phenotype IDs used in the analysis were extracted from the MRC-IEU database. For the analysis, we extracted genetic instruments from FinnGen based GWAS results, to avoid possible bias from overlapping samples. Because MRC-IEU's GWAS data is mainly based on data from UKBB. The results are based on the Inverse variance weighted-model and were statistically significant (P< 0.05). As a sensitivity analysis we also performed analysis by using MR Egger. Nsnp, number of SNPs; pFDR, false discovery rate-corrected p-value; pHET, p-value for heterogeneity; pPLE, p-value for pleiotropy

| Direction | Trait | Method | nsnp | Causal estimate scale | Causal estimate | pFDR | pHET | Egger intercept | pPLE |
|---|---|---|---|---|---|---|---|---|---|
| **LDH-> Trait** | Frequency of tiredness in last 2 weeks \|\| id:ukb-b-929 | IVW | 31 | Beta | 0.02 (0.00–0.05) | 4.61e-02 | 8.22e-08 | | |
| | Frequency of tiredness in last 2 weeks \|\| id:ukb-b-929 | MR Egger | 31 | Beta | 0.07 (-0.05–0.19) | 0.40 | 8.34e-08 | -0.0026 | 0.4579 |
| **LDH-> Trait** | Back pain \|\| id:ukb-b-983 | IVW | 31 | Beta | 0.05 (0.04–0.06) | 6.20e-13 | 2.91e-11 | | |
| | Back pain \|\| id:ukb-b-983 | MR Egger | 31 | Beta | 0.08 (0.01–0.15) | 0.09 | 5.39e-11 | -0.019 | 0.3432 |

**Table S10** List of risk factors that were used in bi-directional Mendelian randomization. We used the Two-Sample MR R library to conduct a bi-directional Mendelian randomization to examine the causal relationships between LDH and its associated risk factors. Risk factors were extracted from the GWAS database provided by the MRC Integrative Epidemiology Unit (IEU) (https://gwas.mrcieu.ac.uk/)

| GWAS-ID | Year | Trait | Consortium | Sample size | Number of SNP's |
|---|---|---|---|---|---|
| **ukb-d-20544_11** | 2018 | Mental health problems ever diagnosed by a professional: Depression | NA | 117 782 | 13 571 547 |
| **ukb-a-525** | 2017 | ICD10: F31 Bipolar affective disorder | Neale Lab | 337 199 | 10 894 596 |
| **ukb-b-16489** | 2018 | Qualifications: College or University degree | MRC-IEU | 458 079 | 9 851 867 |
| **ukb-b-9547** | 2018 | Medication or take exogenous hormones: Oral contraceptive pill or minipill | MRC-IEU | 249 710 | 9 851 867 |
| **ukb-b-3656** | 2018 | Number of treatments/medications taken | MRC-IEU | 462 933 | 9 851 867 |
| **ukb-b-9838** | 2018 | Pain type(s) experienced in last month: Back pain | MRC-IEU | 461 857 | 9 851 867 |
| **ukb-b-929** | 2018 | Frequency of tiredness/lethargy in last 2 weeks | MRC-IEU | 449 019 | 9 851 867 |
| **ieu-a-982** | 2015 | Lumbar spine bone mineral density | GEFOS | 28 498 | 10 582 867 |
| **ukb-b-14177** | 2018 | Vascular/heart problems diagnosed by doctor: High blood pressure | MRC-IEU | 461 880 | 9 851 867 |
| **Ieu-a-93** | 2013 | Overweight | GIANT | 158 855 | 2 435 045 |
| **ukb-b-2002** | 2018 | Job involves heavy manual or physical work | MRC_IEU | 263 615 | 9 851 867 |

**Table S11** A list of FinnGen authors and their affiliations.

| Full Name | Affiliation | Role 1 | Role 2 |
|---|---|---|---|
| **Aarno Palotie** | Institute for Molecular Medicine Finland (FIMM), HiLIFE, University of Helsinki, Helsinki, Finland; Broad Institute of MIT and Harvard; Massachusetts General Hospital | **Steering Committee** | **Steering Committee** |
| **Mark Daly** | Institute for Molecular Medicine Finland (FIMM), HiLIFE, University of Helsinki, Helsinki, Finland; Broad Institute of MIT and Harvard; Massachusetts General Hospital | **Steering Committee** | **Steering Committee** |
| **Bridget Riley-Gills** | Abbvie, Chicago, IL, United States | **Steering Committee** | **Pharmaceutical companies** |
| **Howard Jacob** | Abbvie, Chicago, IL, United States | **Steering Committee** | **Pharmaceutical companies** |
| **Dirk Paul** | Astra Zeneca, Cambridge, United Kingdom | **Steering Committee** | **Pharmaceutical companies** |
| **Slavé Petrovski** | Astra Zeneca, Cambridge, United Kingdom | **Steering Committee** | **Pharmaceutical companies** |
| **Heiko Runz** | Biogen, Cambridge, MA, United States | **Steering Committee** | **Pharmaceutical companies** |
| **Sally John** | Biogen, Cambridge, MA, United States | **Steering Committee** | **Pharmaceutical companies** |
| **George Okafo** | Boehringer Ingelheim, Ingelheim am Rhein, Germany | **Steering Committee** | **Pharmaceutical companies** |
| **Nathan Lawless** | Boehringer Ingelheim, Ingelheim am Rhein, Germany | **Steering Committee** | **Pharmaceutical companies** |
| **Heli Salminen-Mankonen** | Boehringer Ingelheim, Ingelheim am Rhein, Germany | **Steering Committee** | **Pharmaceutical companies** |
| **Robert Plenge** | Bristol Myers Squibb, New York, NY, United States | **Steering Committee** | **Pharmaceutical companies** |
| **Joseph Maranville** | Bristol Myers Squibb, New York, NY, United States | **Steering Committee** | **Pharmaceutical companies** |
| **Mark McCarthy** | Genentech, San Francisco, CA, United States | **Steering Committee** | **Pharmaceutical companies** |
| **Margaret G. Ehm** | GlaxoSmithKline, Collegeville, PA, United States | **Steering Committee** | **Pharmaceutical companies** |
| **Kirsi Auro** | GlaxoSmithKline, Espoo, Finland | **Steering Committee** | **Pharmaceutical companies** |
| **Simonne Longerich** | Merck, Kenilworth, NJ, United States | **Steering Committee** | **Pharmaceutical companies** |
| **Anders Mälarstig** | Pfizer, New York, NY, United States | **Steering Committee** | **Pharmaceutical companies** |
| **Katherine Klinger** | Translational Sciences, Sanofi R&D, Framingham, MA, USA | **Steering Committee** | **Pharmaceutical companies** |
| **Clement Chatelain** | Translational Sciences, Sanofi R&D, Framingham, MA, USA | **Steering Committee** | **Pharmaceutical companies** |
| **Matthias Gossel** | Translational Sciences, Sanofi R&D, Framingham, MA, USA | **Steering Committee** | **Pharmaceutical companies** |
| **Karol Estrada** | Maze Therapeutics, San Francisco, CA, United States | **Steering Committee** | **Pharmaceutical companies** |
| **Robert Graham** | Maze Therapeutics, San Francisco, CA, United States | **Steering Committee** | **Pharmaceutical companies** |
| **Robert Yang** | Janssen Biotech, Beerse, Belgium | **Steering Committee** | **Pharmaceutical companies** |
| **Chris O´Donnell** | Novartis Institutes for BioMedical Research, Cambridge, MA, United States | **Steering Committee** | **Pharmaceutical companies** |
| **Tomi P. Mäkelä** | HiLIFE, University of Helsinki, Finland, Finland | **Steering Committee** | **University of Helsinki & Biobanks** |
| **Jaakko Kaprio** | Institute for Molecular Medicine Finland (FIMM), HiLIFE, University of Helsinki, Helsinki, Finland | **Steering Committee** | **University of Helsinki & Biobanks** |
| **Petri Virolainen** | Auria Biobank / University of Turku / Hospital District of Southwest Finland, Turku, Finland | **Steering Committee** | **University of Helsinki & Biobanks** |
| **Antti Hakanen** | Auria Biobank / University of Turku / Hospital District of Southwest Finland, Turku, Finland | **Steering Committee** | **University of Helsinki & Biobanks** |
| **Terhi Kilpi** | THL Biobank / Finnish Institute for Health and Welfare (THL), Helsinki, Finland | **Steering Committee** | **University of Helsinki & Biobanks** |
| **Markus Perola** | THL Biobank / Finnish Institute for Health and Welfare (THL), Helsinki, Finland | **Steering Committee** | **University of Helsinki & Biobanks** |

| | | | |
|---|---|---|---|
| **Jukka Partanen** | Finnish Red Cross Blood Service / Finnish Hematology Registry and Clinical Biobank, Helsinki, Finland | Steering Committee | **University of Helsinki & Biobanks** |
| **Anne Pitkäranta** | Helsinki Biobank / Helsinki University and Hospital District of Helsinki and Uusimaa, Helsinki | Steering Committee | **University of Helsinki & Biobanks** |
| **Taneli Raivio** | Helsinki Biobank / Helsinki University and Hospital District of Helsinki and Uusimaa, Helsinki | Steering Committee | **University of Helsinki & Biobanks** |
| **Jani Tikkanen** | Northern Finland Biobank Borealis / University of Oulu / Northern Ostrobothnia Hospital District, Oulu, Finland | Steering Committee | **University of Helsinki & Biobanks** |
| **Raisa Serpi** | Northern Finland Biobank Borealis / University of Oulu / Northern Ostrobothnia Hospital District, Oulu, Finland | Steering Committee | **University of Helsinki & Biobanks** |
| **Tarja Laitinen** | Finnish Clinical Biobank Tampere / University of Tampere / Pirkanmaa Hospital District, Tampere, Finland | Steering Committee | **University of Helsinki & Biobanks** |
| **Veli-Matti Kosma** | Biobank of Eastern Finland / University of Eastern Finland / Northern Savo Hospital District, Kuopio, Finland | Steering Committee | **University of Helsinki & Biobanks** |
| **Jari Laukkanen** | Central Finland Biobank / University of Jyväskylä / Central Finland Health Care District, Jyväskylä, Finland | Steering Committee | **University of Helsinki & Biobanks** |
| **Marco Hautalahti** | FINBB - Finnish biobank cooperative | Steering Committee | **University of Helsinki & Biobanks** |
| **Outi Tuovila** | Business Finland, Helsinki, Finland | Steering Committee | **Other Experts/ Non-Voting Members** |
| **Raimo Pakkanen** | Business Finland, Helsinki, Finland | Steering Committee | **Other Experts/ Non-Voting Members** |
| **Jeffrey Waring** | Abbvie, Chicago, IL, United States | Scientific Committee | **Pharmaceutical companies** |
| **Bridget Riley-Gillis** | Abbvie, Chicago, IL, United States | Scientific Committee | **Pharmaceutical companies** |
| **Fedik Rahimov** | Abbvie, Chicago, IL, United States | Scientific Committee | **Pharmaceutical companies** |
| **Ioanna Tachmazidou** | Astra Zeneca, Cambridge, United Kingdom | Scientific Committee | **Pharmaceutical companies** |
| **Chia-Yen Chen** | Biogen, Cambridge, MA, United States | Scientific Committee | **Pharmaceutical companies** |
| **Heiko Runz** | Biogen, Cambridge, MA, United States | Scientific Committee | **Pharmaceutical companies** |
| **Zhihao Ding** | Boehringer Ingelheim, Ingelheim am Rhein, Germany | Scientific Committee | **Pharmaceutical companies** |
| **Marc Jung** | Boehringer Ingelheim, Ingelheim am Rhein, Germany | Scientific Committee | **Pharmaceutical companies** |
| **Shameek Biswas** | Bristol Myers Squibb, New York, NY, United States | Scientific Committee | **Pharmaceutical companies** |
| **Rion Pendergrass** | Genentech, San Francisco, CA, United States | Scientific Committee | **Pharmaceutical companies** |
| **Margaret G. Ehm** | GlaxoSmithKline, Collegeville, PA, United States | Scientific Committee | **Pharmaceutical companies** |
| **David Pulford** | GlaxoSmithKline, Stevenage, United Kingdom | Scientific Committee | **Pharmaceutical companies** |
| **Neha Raghavan** | Merck, Kenilworth, NJ, United States | Scientific Committee | **Pharmaceutical companies** |
| **Adriana Huertas-Vazquez** | Merck, Kenilworth, NJ, United States | Scientific Committee | **Pharmaceutical companies** |
| **Jae-Hoon Sul** | Merck, Kenilworth, NJ, United States | Scientific Committee | **Pharmaceutical companies** |
| **Anders Mälarstig** | Pfizer, New York, NY, United States | Scientific Committee | **Pharmaceutical companies** |
| **Xinli Hu** | Pfizer, New York, NY, United States | Scientific Committee | **Pharmaceutical companies** |
| **Åsa Hedman** | Pfizer, New York, NY, United States | Scientific Committee | **Pharmaceutical companies** |
| **Katherine Klinger** | Translational Sciences, Sanofi R&D, Framingham, MA, USA | Scientific Committee | **Pharmaceutical companies** |
| **Robert Graham** | Maze Therapeutics, San Francisco, CA, United States | Scientific Committee | **Pharmaceutical companies** |

| | | | |
|---|---|---|---|
| **Manuel Rivas** | Maze Therapeutics, San Francisco, CA, United States | Scientific Committee | **Pharmaceutical companies** |
| **Dawn Waterworth** | Janssen Research & Development, LLC, Spring House, PA, United States | Scientific Committee | **Pharmaceutical companies** |
| **Nicole Renaud** | Novartis Institutes for BioMedical Research, Cambridge, MA, United States | Scientific Committee | **Pharmaceutical companies** |
| **Ma´en Obeidat** | Novartis Institutes for BioMedical Research, Cambridge, MA, United States | Scientific Committee | **Pharmaceutical companies** |
| **Samuli Ripatti** | Institute for Molecular Medicine Finland (FIMM), HiLIFE, University of Helsinki, Helsinki, Finland | Scientific Committee | **University of Helsinki & Biobanks** |
| **Johanna Schleutker** | Auria Biobank / Univ. of Turku / Hospital District of Southwest Finland, Turku, Finland | Scientific Committee | **University of Helsinki & Biobanks** |
| **Markus Perola** | THL Biobank / Finnish Institute for Health and Welfare (THL), Helsinki, Finland | Scientific Committee | **University of Helsinki & Biobanks** |
| **Mikko Arvas** | Finnish Red Cross Blood Service / Finnish Hematology Registry and Clinical Biobank, Helsinki, Finland | Scientific Committee | **University of Helsinki & Biobanks** |
| **Olli Carpén** | Helsinki Biobank / Helsinki University and Hospital District of Helsinki and Uusimaa, Helsinki | Scientific Committee | **University of Helsinki & Biobanks** |
| **Reetta Hinttala** | Northern Finland Biobank Borealis / University of Oulu / Northern Ostrobothnia Hospital District, Oulu, Finland | Scientific Committee | **University of Helsinki & Biobanks** |
| **Johannes Kettunen** | Northern Finland Biobank Borealis / University of Oulu / Northern Ostrobothnia Hospital District, Oulu, Finland | Scientific Committee | **University of Helsinki & Biobanks** |
| **Arto Mannermaa** | Biobank of Eastern Finland / University of Eastern Finland / Northern Savo Hospital District, Kuopio, Finland | Scientific Committee | **University of Helsinki & Biobanks** |
| **Katriina Aalto-Setälä** | Faculty of Medicine and Health Technology, Tampere University, Tampere, Finland | Scientific Committee | **University of Helsinki & Biobanks** |
| **Mika Kähönen** | Finnish Clinical Biobank Tampere / University of Tampere / Pirkanmaa Hospital District, Tampere, Finland | Scientific Committee | **University of Helsinki & Biobanks** |
| **Jari Laukkanen** | Central Finland Biobank / University of Jyväskylä / Central Finland Health Care District, Jyväskylä, Finland | Scientific Committee | **University of Helsinki & Biobanks** |
| **Johanna Mäkelä** | FINBB - Finnish biobank cooperative | Scientific Committee | **University of Helsinki & Biobanks** |
| **Reetta Kälviäinen** | Northern Savo Hospital District, Kuopio, Finland | Clinical Groups | **Neurology Group** |
| **Valtteri Julkunen** | Northern Savo Hospital District, Kuopio, Finland | Clinical Groups | **Neurology Group** |
| **Hilkka Soininen** | Northern Savo Hospital District, Kuopio, Finland | Clinical Groups | **Neurology Group** |
| **Anne Remes** | Northern Ostrobothnia Hospital District, Oulu, Finland | Clinical Groups | **Neurology Group** |
| **Mikko Hiltunen** | University of Eastern Finland, Kuopio, Finland | Clinical Groups | **Neurology Group** |
| **Jukka Peltola** | Pirkanmaa Hospital District, Tampere, Finland | Clinical Groups | **Neurology Group** |
| **Minna Raivio** | Hospital District of Helsinki and Uusimaa, Helsinki, Finland | Clinical Groups | **Neurology Group** |
| **Pentti Tienari** | Hospital District of Helsinki and Uusimaa, Helsinki, Finland | Clinical Groups | **Neurology Group** |
| **Juha Rinne** | Hospital District of Southwest Finland, Turku, Finland | Clinical Groups | **Neurology Group** |
| **Roosa Kallionpää** | Hospital District of Southwest Finland, Turku, Finland | Clinical Groups | **Neurology Group** |
| **Juulia Partanen** | Institute for Molecular Medicine Finland, HiLIFE, University of Helsinki, Finland | Clinical Groups | **Neurology Group** |
| **Ali Abbasi** | Abbvie, Chicago, IL, United States | Clinical Groups | **Neurology Group** |
| **Adam Ziemann** | Abbvie, Chicago, IL, United States | Clinical Groups | **Neurology Group** |
| **Nizar Smaoui** | Abbvie, Chicago, IL, United States | Clinical Groups | **Neurology Group** |
| **Anne Lehtonen** | Abbvie, Chicago, IL, United States | Clinical Groups | **Neurology Group** |

| | | | |
|---|---|---|---|
| **Susan Eaton** | Biogen, Cambridge, MA, United States | Clinical Groups | **Neurology Group** |
| **Heiko Runz** | Biogen, Cambridge, MA, United States | Clinical Groups | **Neurology Group** |
| **Sanni Lahdenperä** | Biogen, Cambridge, MA, United States | Clinical Groups | **Neurology Group** |
| **Shameek Biswas** | Bristol Myers Squibb, New York, NY, United States | Clinical Groups | **Neurology Group** |
| **Natalie Bowers** | Genentech, San Francisco, CA, United States | Clinical Groups | **Neurology Group** |
| **Edmond Teng** | Genentech, San Francisco, CA, United States | Clinical Groups | **Neurology Group** |
| **Rion Pendergrass** | Genentech, San Francisco, CA, United States | Clinical Groups | **Neurology Group** |
| **Fanli Xu** | GlaxoSmithKline, Brentford, United Kingdom | Clinical Groups | **Neurology Group** |
| **David Pulford** | GlaxoSmithKline, Stevenage, United Kingdom | Clinical Groups | **Neurology Group** |
| **Kirsi Auro** | GlaxoSmithKline, Espoo, Finland | Clinical Groups | **Neurology Group** |
| **Laura Addis** | GlaxoSmithKline, Brentford, United Kingdom | Clinical Groups | **Neurology Group** |
| **John Eicher** | GlaxoSmithKline, Brentford, United Kingdom | Clinical Groups | **Neurology Group** |
| **Qingqin S Li** | Janssen Research & Development, LLC, Titusville, NJ 08560, United States | Clinical Groups | **Neurology Group** |
| **Karen He** | Janssen Research & Development, LLC, Spring House, PA, United States | Clinical Groups | **Neurology Group** |
| **Ekaterina Khramtsova** | Janssen Research & Development, LLC, Spring House, PA, United States | Clinical Groups | **Neurology Group** |
| **Neha Raghavan** | Merck, Kenilworth, NJ, United States | Clinical Groups | **Neurology Group** |
| **Martti Färkkilä** | Hospital District of Helsinki and Uusimaa, Helsinki, Finland | Clinical Groups | **Gastroenterology Group** |
| **Jukka Koskela** | Hospital District of Helsinki and Uusimaa, Helsinki, Finland | Clinical Groups | **Gastroenterology Group** |
| **Sampsa Pikkarainen** | Hospital District of Helsinki and Uusimaa, Helsinki, Finland | Clinical Groups | **Gastroenterology Group** |
| **Airi Jussila** | Pirkanmaa Hospital District, Tampere, Finland | Clinical Groups | **Gastroenterology Group** |
| **Katri Kaukinen** | Pirkanmaa Hospital District, Tampere, Finland | Clinical Groups | **Gastroenterology Group** |
| **Timo Blomster** | Northern Ostrobothnia Hospital District, Oulu, Finland | Clinical Groups | **Gastroenterology Group** |
| **Mikko Kiviniemi** | Northern Savo Hospital District, Kuopio, Finland | Clinical Groups | **Gastroenterology Group** |
| **Markku Voutilainen** | Hospital District of Southwest Finland, Turku, Finland | Clinical Groups | **Gastroenterology Group** |
| **Mark Daly** | Institute for Molecular Medicine, Finland (FIMM), HiLIFE, University of Helsinki, Helsinki, Finland; Broad Institute of MIT and Harvard; Massachusetts General Hospital | Clinical Groups | **Gastroenterology Group** |
| **Ali Abbasi** | Abbvie, Chicago, IL, United States | Clinical Groups | **Gastroenterology Group** |
| **Jeffrey Waring** | Abbvie, Chicago, IL, United States | Clinical Groups | **Gastroenterology Group** |
| **Nizar Smaoui** | Abbvie, Chicago, IL, United States | Clinical Groups | **Gastroenterology Group** |
| **Fedik Rahimov** | Abbvie, Chicago, IL, United States | Clinical Groups | **Gastroenterology Group** |
| **Anne Lehtonen** | Abbvie, Chicago, IL, United States | Clinical Groups | **Gastroenterology Group** |
| **Tim Lu** | Genentech, San Francisco, CA, United States | Clinical Groups | **Gastroenterology Group** |
| **Natalie Bowers** | Genentech, San Francisco, CA, United States | Clinical Groups | **Gastroenterology Group** |
| **Rion Pendergrass** | Genentech, San Francisco, CA, United States | Clinical Groups | **Gastroenterology Group** |
| **Linda McCarthy** | GlaxoSmithKline, Brentford, United Kingdom | Clinical Groups | **Gastroenterology Group** |
| **Amy Hart** | Janssen Research & Development, LLC, Spring House, PA, United States | Clinical Groups | **Gastroenterology Group** |
| **Meijian Guan** | Janssen Research & Development, LLC, Spring House, PA, United States | Clinical Groups | **Gastroenterology Group** |

| | | | |
|---|---|---|---|
| **Jason Miller** | Merck, Kenilworth, NJ, United States | Clinical Groups | **Gastroenterology Group** |
| **Kirsi Kalpala** | Pfizer, New York, NY, United States | Clinical Groups | **Gastroenterology Group** |
| **Melissa Miller** | Pfizer, New York, NY, United States | Clinical Groups | **Gastroenterology Group** |
| **Xinli Hu** | Pfizer, New York, NY, United States | Clinical Groups | **Gastroenterology Group** |
| **Kari Eklund** | Hospital District of Helsinki and Uusimaa, Helsinki, Finland | Clinical Groups | **Rheumatology Group** |
| **Antti Palomäki** | Hospital District of Southwest Finland, Turku, Finland | Clinical Groups | **Rheumatology Group** |
| **Pia Isomäki** | Pirkanmaa Hospital District, Tampere, Finland | Clinical Groups | **Rheumatology Group** |
| **Laura Pirilä** | Hospital District of Southwest Finland, Turku, Finland | Clinical Groups | **Rheumatology Group** |
| **Oili Kaipiainen-Seppänen** | Northern Savo Hospital District, Kuopio, Finland | Clinical Groups | **Rheumatology Group** |
| **Johanna Huhtakangas** | Northern Ostrobothnia Hospital District, Oulu, Finland | Clinical Groups | **Rheumatology Group** |
| **Nina Mars** | Institute for Molecular Medicine Finland (FIMM), HiLIFE, University of Helsinki, Helsinki, Finland | Clinical Groups | **Rheumatology Group** |
| **Ali Abbasi** | Abbvie, Chicago, IL, United States | Clinical Groups | **Rheumatology Group** |
| **Jeffrey Waring** | Abbvie, Chicago, IL, United States | Clinical Groups | **Rheumatology Group** |
| **Fedik Rahimov** | Abbvie, Chicago, IL, United States | Clinical Groups | **Rheumatology Group** |
| **Apinya Lertratanakul** | Abbvie, Chicago, IL, United States | Clinical Groups | **Rheumatology Group** |
| **Nizar Smaoui** | Abbvie, Chicago, IL, United States | Clinical Groups | **Rheumatology Group** |
| **Anne Lehtonen** | Abbvie, Chicago, IL, United States | Clinical Groups | **Rheumatology Group** |
| **Coralie Viollet** | AstraZeneca, Cambridge, United Kingdom | Clinical Groups | **Rheumatology Group** |
| **Marla Hochfeld** | Bristol Myers Squibb, New York, NY, United States | Clinical Groups | **Rheumatology Group** |
| **Natalie Bowers** | Genentech, San Francisco, CA, United States | Clinical Groups | **Rheumatology Group** |
| **Rion Pendergrass** | Genentech, San Francisco, CA, United States | Clinical Groups | **Rheumatology Group** |
| **Jorge Esparza Gordillo** | GlaxoSmithKline, Brentford, United Kingdom | Clinical Groups | **Rheumatology Group** |
| **Kirsi Auro** | GlaxoSmithKline, Espoo, Finland | Clinical Groups | **Rheumatology Group** |
| **Dawn Waterworth** | Janssen Research & Development, LLC, Spring House, PA, United States | Clinical Groups | **Rheumatology Group** |
| **Fabiana Farias** | Merck, Kenilworth, NJ, United States | Clinical Groups | **Rheumatology Group** |
| **Kirsi Kalpala** | Pfizer, New York, NY, United States | Clinical Groups | **Rheumatology Group** |
| **Nan Bing** | Pfizer, New York, NY, United States | Clinical Groups | **Rheumatology Group** |
| **Xinli Hu** | Pfizer, New York, NY, United States | Clinical Groups | **Rheumatology Group** |
| **Tarja Laitinen** | Pirkanmaa Hospital District, Tampere, Finland | Clinical Groups | **Pulmonology Group** |
| **Margit Pelkonen** | Northern Savo Hospital District, Kuopio, Finland | Clinical Groups | **Pulmonology Group** |
| **Paula Kauppi** | Hospital District of Helsinki and Uusimaa, Helsinki, Finland | Clinical Groups | **Pulmonology Group** |
| **Hannu Kankaanranta** | University of Gothenburg, Gothenburg, Sweden/ Seinäjoki Central Hospital, Seinäjoki, Finland/ Tampere University, Tampere, Finland | Clinical Groups | **Pulmonology Group** |
| **Terttu Harju** | Northern Ostrobothnia Hospital District, Oulu, Finland | Clinical Groups | **Pulmonology Group** |
| **Riitta Lahesmaa** | Hospital District of Southwest Finland, Turku, Finland | Clinical Groups | **Pulmonology Group** |
| **Nizar Smaoui** | Abbvie, Chicago, IL, United States | Clinical Groups | **Pulmonology Group** |
| **Coralie Viollet** | AstraZeneca, Cambridge, United Kingdom | Clinical Groups | **Pulmonology Group** |

| | | | |
|---|---|---|---|
| **Susan Eaton** | Biogen, Cambridge, MA, United States | Clinical Groups | **Pulmonology Group** |
| **Hubert Chen** | Genentech, San Francisco, CA, United States | Clinical Groups | **Pulmonology Group** |
| **Rion Pendergrass** | Genentech, San Francisco, CA, United States | Clinical Groups | **Pulmonology Group** |
| **Natalie Bowers** | Genentech, San Francisco, CA, United States | Clinical Groups | **Pulmonology Group** |
| **Joanna Betts** | GlaxoSmithKline, Brentford, United Kingdom | Clinical Groups | **Pulmonology Group** |
| **Kirsi Auro** | GlaxoSmithKline, Espoo, Finland | Clinical Groups | **Pulmonology Group** |
| **Rajashree Mishra** | GlaxoSmithKline, Brentford, United Kingdom | Clinical Groups | **Pulmonology Group** |
| **Majd Mouded** | Novartis, Basel, Switzerland | Clinical Groups | **Pulmonology Group** |
| **Debby Ngo** | Novartis, Basel, Switzerland | Clinical Groups | **Pulmonology Group** |
| **Teemu Niiranen** | Finnish Institute for Health and Welfare (THL), Helsinki, Finland | Clinical Groups | **Cardiometabolic Diseases Group** |
| **Felix Vaura** | Finnish Institute for Health and Welfare (THL), Helsinki, Finland | Clinical Groups | **Cardiometabolic Diseases Group** |
| **Veikko Salomaa** | Finnish Institute for Health and Welfare (THL), Helsinki, Finland | Clinical Groups | **Cardiometabolic Diseases Group** |
| **Kaj Metsärinne** | Hospital District of Southwest Finland, Turku, Finland | Clinical Groups | **Cardiometabolic Diseases Group** |
| **Jenni Aittokallio** | Hospital District of Southwest Finland, Turku, Finland | Clinical Groups | **Cardiometabolic Diseases Group** |
| **Mika Kähönen** | Pirkanmaa Hospital District, Tampere, Finland | Clinical Groups | **Cardiometabolic Diseases Group** |
| **Jussi Hernesniemi** | Pirkanmaa Hospital District, Tampere, Finland | Clinical Groups | **Cardiometabolic Diseases Group** |
| **Daniel Gordin** | Hospital District of Helsinki and Uusimaa, Helsinki, Finland | Clinical Groups | **Cardiometabolic Diseases Group** |
| **Juha Sinisalo** | Hospital District of Helsinki and Uusimaa, Helsinki, Finland | Clinical Groups | **Cardiometabolic Diseases Group** |
| **Marja-Riitta Taskinen** | Hospital District of Helsinki and Uusimaa, Helsinki, Finland | Clinical Groups | **Cardiometabolic Diseases Group** |
| **Tiinamaija Tuomi** | Hospital District of Helsinki and Uusimaa, Helsinki, Finland | Clinical Groups | **Cardiometabolic Diseases Group** |
| **Timo Hiltunen** | Hospital District of Helsinki and Uusimaa, Helsinki, Finland | Clinical Groups | **Cardiometabolic Diseases Group** |
| **Jari Laukkanen** | Central Finland Health Care District, Jyväskylä, Finland | Clinical Groups | **Cardiometabolic Diseases Group** |
| **Amanda Elliott** | Institute for Molecular Medicine Finland (FIMM), HiLIFE, University of Helsinki, Helsinki, Finland; Broad Institute, Cambridge, MA, USA and Massachusetts General Hospital, Boston, MA, USA | Clinical Groups | **Cardiometabolic Diseases Group** |
| **Mary Pat Reeve** | Institute for Molecular Medicine Finland (FIMM), HiLIFE, University of Helsinki, Helsinki, Finland | Clinical Groups | **Cardiometabolic Diseases Group** |
| **Sanni Ruotsalainen** | Institute for Molecular Medicine Finland (FIMM), HiLIFE, University of Helsinki, Helsinki, Finland | Clinical Groups | **Cardiometabolic Diseases Group** |
| **Dirk Paul** | Astra Zeneca, Cambridge, United Kingdom | Clinical Groups | **Cardiometabolic Diseases Group** |
| **Natalie Bowers** | Genentech, San Francisco, CA, United States | Clinical Groups | **Cardiometabolic Diseases Group** |
| **Rion Pendergrass** | Genentech, San Francisco, CA, United States | Clinical Groups | **Cardiometabolic Diseases Group** |
| **Audrey Chu** | GlaxoSmithKline, Brentford, United Kingdom | Clinical Groups | **Cardiometabolic Diseases Group** |
| **Kirsi Auro** | GlaxoSmithKline, Espoo, Finland | Clinical Groups | **Cardiometabolic Diseases Group** |
| **Dermot Reilly** | Janssen Research & Development, LLC, Boston, MA, United States | Clinical Groups | **Cardiometabolic Diseases Group** |
| **Mike Mendelson** | Novartis, Boston, MA, United States | Clinical Groups | **Cardiometabolic Diseases Group** |
| **Jaakko Parkkinen** | Pfizer, New York, NY, United States | Clinical Groups | **Cardiometabolic Diseases Group** |
| **Melissa Miller** | Pfizer, New York, NY, United States | Clinical Groups | **Cardiometabolic Diseases Group** |
| **Tuomo Meretoja** | Hospital District of Helsinki and Uusimaa, Helsinki, Finland | Clinical Groups | **Oncology Group** |

| | | | |
|---|---|---|---|
| **Heikki Joensuu** | Hospital District of Helsinki and Uusimaa, Helsinki, Finland | Clinical Groups | **Oncology Group** |
| **Olli Carpén** | Hospital District of Helsinki and Uusimaa, Helsinki, Finland | Clinical Groups | **Oncology Group** |
| **Johanna Mattson** | Hospital District of Helsinki and Uusimaa, Helsinki, Finland | Clinical Groups | **Oncology Group** |
| **Eveliina Salminen** | Hospital District of Helsinki and Uusimaa, Helsinki, Finland | Clinical Groups | **Oncology Group** |
| **Annika Auranen** | Pirkanmaa Hospital District , Tampere, Finland | Clinical Groups | **Oncology Group** |
| **Peeter Karihtala** | Northern Ostrobothnia Hospital District, Oulu, Finland | Clinical Groups | **Oncology Group** |
| **Päivi Auvinen** | Northern Savo Hospital District, Kuopio, Finland | Clinical Groups | **Oncology Group** |
| **Klaus Elenius** | Hospital District of Southwest Finland, Turku, Finland | Clinical Groups | **Oncology Group** |
| **Johanna Schleutker** | Hospital District of Southwest Finland, Turku, Finland | Clinical Groups | **Oncology Group** |
| **Esa Pitkänen** | Institute for Molecular Medicine Finland (FIMM), HiLIFE, University of Helsinki, Helsinki, Finland | Clinical Groups | **Oncology Group** |
| **Nina Mars** | Institute for Molecular Medicine Finland (FIMM), HiLIFE, University of Helsinki, Helsinki, Finland | Clinical Groups | **Oncology Group** |
| **Mark Daly** | Institute for Molecular Medicine Finland (FIMM), HiLIFE, University of Helsinki, Helsinki, Finland; Broad Institute of MIT and Harvard; Massachusetts General Hospital | Clinical Groups | **Oncology Group** |
| **Relja Popovic** | Abbvie, Chicago, IL, United States | Clinical Groups | **Oncology Group** |
| **Jeffrey Waring** | Abbvie, Chicago, IL, United States | Clinical Groups | **Oncology Group** |
| **Bridget Riley-Gillis** | Abbvie, Chicago, IL, United States | Clinical Groups | **Oncology Group** |
| **Anne Lehtonen** | Abbvie, Chicago, IL, United States | Clinical Groups | **Oncology Group** |
| **Margarete Fabre** | AstraZeneca, Cambridge, United Kingdom | Clinical Groups | **Oncology Group** |
| **Jennifer Schutzman** | Genentech, San Francisco, CA, United States | Clinical Groups | **Oncology Group** |
| **Natalie Bowers** | Genentech, San Francisco, CA, United States | Clinical Groups | **Oncology Group** |
| **Rion Pendergrass** | Genentech, San Francisco, CA, United States | Clinical Groups | **Oncology Group** |
| **Diptee Kulkarni** | GlaxoSmithKline, Brentford, United Kingdom | Clinical Groups | **Oncology Group** |
| **Kirsi Auro** | GlaxoSmithKline, Espoo, Finland | Clinical Groups | **Oncology Group** |
| **Alessandro Porello** | Janssen Research & Development, LLC, Spring House, PA, United States | Clinical Groups | **Oncology Group** |
| **Andrey Loboda** | Merck, Kenilworth, NJ, United States | Clinical Groups | **Oncology Group** |
| **Heli Lehtonen** | Pfizer, New York, NY, United States | Clinical Groups | **Oncology Group** |
| **Stefan McDonough** | Pfizer, New York, NY, United States | Clinical Groups | **Oncology Group** |
| **Sauli Vuoti** | Janssen-Cilag Oy, Espoo, Finland | Clinical Groups | **Oncology Group** |
| **Kai Kaarniranta** | Northern Savo Hospital District, Kuopio, Finland; Department of Molecular Genetics, University of Lodz, Lodz, Poland | Clinical Groups | **Opthalmology Group** |
| **Joni A Turunen** | Helsinki University Hospital and University of Helsinki, Helsinki, Finland; Eye Genetics Group, Folkhälsan Research Center, Helsinki, Finland | Clinical Groups | **Opthalmology Group** |
| **Terhi Ollila** | Hospital District of Helsinki and Uusimaa, Helsinki, Finland | Clinical Groups | **Opthalmology Group** |
| **Hannu Uusitalo** | Pirkanmaa Hospital District, Tampere, Finland | Clinical Groups | **Opthalmology Group** |
| **Juha Karjalainen** | Institute for Molecular Medicine Finland (FIMM), HiLIFE, University of Helsinki, Helsinki, Finland | Clinical Groups | **Opthalmology Group** |
| **Esa Pitkänen** | Institute for Molecular Medicine Finland (FIMM), HiLIFE, University of Helsinki, Helsinki, Finland | Clinical Groups | **Opthalmology Group** |
| **Mengzhen Liu** | Abbvie, Chicago, IL, United States | Clinical Groups | **Opthalmology Group** |
| **Heiko Runz** | Biogen, Cambridge, MA, United States | Clinical Groups | **Opthalmology Group** |

| | | | |
|---|---|---|---|
| **Stephanie Loomis** | Biogen, Cambridge, MA, United States | Clinical Groups | **Opthalmology Group** |
| **Erich Strauss** | Genentech, San Francisco, CA, United States | Clinical Groups | **Opthalmology Group** |
| **Natalie Bowers** | Genentech, San Francisco, CA, United States | Clinical Groups | **Opthalmology Group** |
| **Hao Chen** | Genentech, San Francisco, CA, United States | Clinical Groups | **Opthalmology Group** |
| **Rion Pendergrass** | Genentech, San Francisco, CA, United States | Clinical Groups | **Opthalmology Group** |
| **Kaisa Tasanen** | Northern Ostrobothnia Hospital District, Oulu, Finland | Clinical Groups | **Dermatology Group** |
| **Laura Huilaja** | Northern Ostrobothnia Hospital District, Oulu, Finland | Clinical Groups | **Dermatology Group** |
| **Katariina Hannula-Jouppi** | Hospital District of Helsinki and Uusimaa, Helsinki, Finland | Clinical Groups | **Dermatology Group** |
| **Teea Salmi** | Pirkanmaa Hospital District, Tampere, Finland | Clinical Groups | **Dermatology Group** |
| **Sirkku Peltonen** | Hospital District of Southwest Finland, Turku, Finland | Clinical Groups | **Dermatology Group** |
| **Leena Koulu** | Hospital District of Southwest Finland, Turku, Finland | Clinical Groups | **Dermatology Group** |
| **Nizar Smaoui** | Abbvie, Chicago, IL, United States | Clinical Groups | **Dermatology Group** |
| **Fedik Rahimov** | Abbvie, Chicago, IL, United States | Clinical Groups | **Dermatology Group** |
| **Anne Lehtonen** | Abbvie, Chicago, IL, United States | Clinical Groups | **Dermatology Group** |
| **David Choy** | Genentech, San Francisco, CA, United States | Clinical Groups | **Dermatology Group** |
| **Rion Pendergrass** | Genentech, San Francisco, CA, United States | Clinical Groups | **Dermatology Group** |
| **Dawn Waterworth** | Janssen Research & Development, LLC, Spring House, PA, United States | Clinical Groups | **Dermatology Group** |
| **Kirsi Kalpala** | Pfizer, New York, NY, United States | Clinical Groups | **Dermatology Group** |
| **Ying Wu** | Pfizer, New York, NY, United States | Clinical Groups | **Dermatology Group** |
| **Pirkko Pussinen** | Hospital District of Helsinki and Uusimaa, Helsinki, Finland | Clinical Groups | **Odontology Group** |
| **Aino Salminen** | Hospital District of Helsinki and Uusimaa, Helsinki, Finland | Clinical Groups | **Odontology Group** |
| **Tuula Salo** | Hospital District of Helsinki and Uusimaa, Helsinki, Finland | Clinical Groups | **Odontology Group** |
| **David Rice** | Hospital District of Helsinki and Uusimaa, Helsinki, Finland | Clinical Groups | **Odontology Group** |
| **Pekka Nieminen** | Hospital District of Helsinki and Uusimaa, Helsinki, Finland | Clinical Groups | **Odontology Group** |
| **Ulla Palotie** | Hospital District of Helsinki and Uusimaa, Helsinki, Finland | Clinical Groups | **Odontology Group** |
| **Maria Siponen** | Northern Savo Hospital District, Kuopio, Finland | Clinical Groups | **Odontology Group** |
| **Liisa Suominen** | Northern Savo Hospital District, Kuopio, Finland | Clinical Groups | **Odontology Group** |
| **Päivi Mäntylä** | Northern Savo Hospital District, Kuopio, Finland | Clinical Groups | **Odontology Group** |
| **Ulvi Gursoy** | Hospital District of Southwest Finland, Turku, Finland | Clinical Groups | **Odontology Group** |
| **Vuokko Anttonen** | Northern Ostrobothnia Hospital District, Oulu, Finland | Clinical Groups | **Odontology Group** |
| **Kirsi Sipilä** | Research Unit of Oral Health Sciences Faculty of Medicine, University of Oulu, Oulu, Finland; Medical Research Center, Oulu, Oulu University Hospital and University of Oulu, Oulu, Finland | Clinical Groups | **Odontology Group** |
| **Rion Pendergrass** | Genentech, San Francisco, CA, United States | Clinical Groups | **Odontology Group** |
| **Hannele Laivuori** | Institute for Molecular Medicine Finland (FIMM), HiLIFE, University of Helsinki, Helsinki, Finland | Clinical Groups | **Women's Health and Reproduction Group** |
| **Venla Kurra** | Pirkanmaa Hospital District, Tampere, Finland | Clinical Groups | **Women's Health and** |

| | | | |
|---|---|---|---|
| | | | Reproduction Group |
| **Laura Kotaniemi-Talonen** | Pirkanmaa Hospital District, Tampere, Finland | Clinical Groups | **Women's Health and Reproduction Group** |
| **Oskari Heikinheimo** | Hospital District of Helsinki and Uusimaa, Helsinki, Finland | Clinical Groups | **Women's Health and Reproduction Group** |
| **Ilkka Kalliala** | Hospital District of Helsinki and Uusimaa, Helsinki, Finland | Clinical Groups | **Women's Health and Reproduction Group** |
| **Lauri Aaltonen** | Hospital District of Helsinki and Uusimaa, Helsinki, Finland | Clinical Groups | **Women's Health and Reproduction Group** |
| **Varpu Jokimaa** | Hospital District of Southwest Finland, Turku, Finland | Clinical Groups | **Women's Health and Reproduction Group** |
| **Johannes Kettunen** | Northern Ostrobothnia Hospital District, Oulu, Finland | Clinical Groups | **Women's Health and Reproduction Group** |
| **Marja Vääräsmäki** | Northern Ostrobothnia Hospital District, Oulu, Finland | Clinical Groups | **Women's Health and Reproduction Group** |
| **Outi Uimari** | Northern Ostrobothnia Hospital District, Oulu, Finland | Clinical Groups | **Women's Health and Reproduction Group** |
| **Laure Morin-Papunen** | Northern Ostrobothnia Hospital District, Oulu, Finland | Clinical Groups | **Women's Health and Reproduction Group** |
| **Maarit Niinimäki** | Northern Ostrobothnia Hospital District, Oulu, Finland | Clinical Groups | **Women's Health and Reproduction Group** |
| **Terhi Piltonen** | Northern Ostrobothnia Hospital District, Oulu, Finland | Clinical Groups | **Women's Health and Reproduction Group** |
| **Katja Kivinen** | Institute for Molecular Medicine Finland (FIMM), HiLIFE, University of Helsinki, Helsinki, Finland | Clinical Groups | **Women's Health and Reproduction Group** |
| **Elisabeth Widen** | Institute for Molecular Medicine Finland (FIMM), HiLIFE, University of Helsinki, Helsinki, Finland | Clinical Groups | **Women's Health and Reproduction Group** |
| **Taru Tukiainen** | Institute for Molecular Medicine Finland (FIMM), HiLIFE, University of Helsinki, Helsinki, Finland | Clinical Groups | **Women's Health and Reproduction Group** |
| **Mary Pat Reeve** | Institute for Molecular Medicine Finland (FIMM), HiLIFE, University of Helsinki, Helsinki, Finland | Clinical Groups | **Women's Health and Reproduction Group** |
| **Mark Daly** | Institute for Molecular Medicine Finland (FIMM), HiLIFE, University of Helsinki, Helsinki, Finland; Broad Institute of MIT and Harvard; Massachusetts General Hospital | Clinical Groups | **Women's Health and Reproduction Group** |
| **Niko Välimäki** | University of Helsinki, Helsinki, Finland | Clinical Groups | **Women's Health and Reproduction Group** |
| **Eija Laakkonen** | University of Jyväskylä, Jyväskylä, Finland | Clinical Groups | **Women's Health and Reproduction Group** |

| | | | |
|---|---|---|---|
| **Jaakko Tyrmi** | University of Oulu, Oulu, Finland / University of Tampere, Tampere, Finland | Clinical Groups | **Women's Health and Reproduction Group** |
| **Heidi Silven** | University of Oulu, Oulu, Finland | Clinical Groups | **Women's Health and Reproduction Group** |
| **Eeva Sliz** | University of Oulu, Oulu, Finland | Clinical Groups | **Women's Health and Reproduction Group** |
| **Riikka Arffman** | University of Oulu, Oulu, Finland | Clinical Groups | **Women's Health and Reproduction Group** |
| **Susanna Savukoski** | University of Oulu, Oulu, Finland | Clinical Groups | **Women's Health and Reproduction Group** |
| **Triin Laisk** | Estonian biobank, Tartu, Estonia | Clinical Groups | **Women's Health and Reproduction Group** |
| **Natalia Pujol** | Estonian biobank, Tartu, Estonia | Clinical Groups | **Women's Health and Reproduction Group** |
| **Mengzhen Liu** | Abbvie, Chicago, IL, United States | Clinical Groups | **Women's Health and Reproduction Group** |
| **Bridget Riley-Gillis** | Abbvie, Chicago, IL, United States | Clinical Groups | **Women's Health and Reproduction Group** |
| **Rion Pendergrass** | Genentech, San Francisco, CA, United States | Clinical Groups | **Women's Health and Reproduction Group** |
| **Janet Kumar** | GlaxoSmithKline, Collegeville, PA, United States | Clinical Groups | **Women's Health and Reproduction Group** |
| **Kirsi Auro** | GlaxoSmithKline, Espoo, Finland | Clinical Groups | **Women's Health and Reproduction Group** |
| **Iiris Hovatta** | University of Helsinki, Finland | Clinical Groups | **Depression group** |
| **Chia-Yen Chen** | Biogen, Cambridge, MA, United States | Clinical Groups | **Depression group** |
| **Erkki Isometsä** | Hospital District of Helsinki and Uusimaa, Helsinki, Finland | Clinical Groups | **Depression group** |
| **Hanna Ollila** | Institute for Molecular Medicine Finland (FIMM), HiLIFE, University of Helsinki, Helsinki, Finland | Clinical Groups | **Depression group** |
| **Jaana Suvisaari** | Finnish Institute for Health and Welfare (THL), Helsinki, Finland | Clinical Groups | **Depression group** |
| **Thomas Damm Als** | Aarhus University, Denmark | Clinical Groups | **Depression group** |
| **Antti Mäkitie** | Department of Otorhinolaryngology - Head and Neck Surgery, University of Helsinki and Helsinki University Hospital, Helsinki, Finland | Clinical Groups | **ENT (ear, nose and throath) Group** |
| **Argyro Bizaki-Vallaskangas** | Pirkanmaa Hospital District, Tampere, Finland | Clinical Groups | **ENT (ear, nose and throath) Group** |
| **Sanna Toppila-Salmi** | University of Eastern Finland and Kuopio University Hospital, Department of Otorhinolaryngology, Kuopio, Finland and Department of Allergy, Helsinki University Hospital and University of Helsinki, Finland | Clinical Groups | **ENT (ear, nose and throath) Group** |
| **Tytti Willberg** | Hospital District of Southwest Finland, Turku, Finland | Clinical Groups | **ENT (ear, nose and throath) Group** |

| | | | |
|---|---|---|---|
| **Elmo Saarentaus** | Institute for Molecular Medicine Finland (FIMM), HiLIFE, University of Helsinki, Helsinki, Finland | Clinical Groups | **ENT (ear, nose and throath) Group** |
| **Antti Aarnisalo** | Hospital District of Helsinki and Uusimaa, Helsinki, Finland | Clinical Groups | **ENT (ear, nose and throath) Group** |
| **Eveliina Salminen** | Hospital District of Helsinki and Uusimaa, Helsinki, Finland | Clinical Groups | **ENT (ear, nose and throath) Group** |
| **Elisa Rahikkala** | Northern Ostrobothnia Hospital District, Oulu, Finland | Clinical Groups | **ENT (ear, nose and throath) Group** |
| **Johannes Kettunen** | Northern Ostrobothnia Hospital District, Oulu, Finland | Clinical Groups | **ENT (ear, nose and throath) Group** |
| **Kristiina Aittomäki** | Department of Medical Genetics, Helsinki University Central Hospital, Helsinki, Finland | Clinical Groups | **POI (premature ovarian failure) Group** |
| **Fredrik Åberg** | Transplantation and Liver Surgery Clinic, Helsinki University Hospital, Helsinki University, Helsinki, Finland | Clinical Groups | **LiverScore Group** |
| **Mitja Kurki** | Institute for Molecular Medicine Finland (FIMM), HiLIFE, University of Helsinki, Helsinki, Finland; Broad Institute, Cambridge, MA, United States | FinnGen Analysis working group | FinnGen Analysis working group |
| **Samuli Ripatti** | Institute for Molecular Medicine Finland (FIMM), HiLIFE, University of Helsinki, Helsinki, Finland | FinnGen Analysis working group | FinnGen Analysis working group |
| **Mark Daly** | Institute for Molecular Medicine, Finland (FIMM), HiLIFE, University of Helsinki, Helsinki, Finland; Broad Institute of MIT and Harvard; Massachusetts General Hospital | FinnGen Analysis working group | FinnGen Analysis working group |
| **Juha Karjalainen** | Institute for Molecular Medicine Finland (FIMM), HiLIFE, University of Helsinki, Helsinki, Finland | FinnGen Analysis working group | FinnGen Analysis working group |
| **Aki Havulinna** | Institute for Molecular Medicine Finland (FIMM), HiLIFE, University of Helsinki, Helsinki, Finland; Finnish Institute for Health and Welfare (THL), Helsinki, Finland | FinnGen Analysis working group | FinnGen Analysis working group |
| **Juha Mehtonen** | Institute for Molecular Medicine Finland (FIMM), HiLIFE, University of Helsinki, Helsinki, Finland | FinnGen Analysis working group | FinnGen Analysis working group |
| **Priit Palta** | Institute for Molecular Medicine Finland (FIMM), HiLIFE, University of Helsinki, Helsinki, Finland | FinnGen Analysis working group | FinnGen Analysis working group |
| **Shabbeer Hassan** | Institute for Molecular Medicine Finland (FIMM), HiLIFE, University of Helsinki, Helsinki, Finland | FinnGen Analysis working group | FinnGen Analysis working group |
| **Pietro Della Briotta Parolo** | Institute for Molecular Medicine Finland (FIMM), HiLIFE, University of Helsinki, Helsinki, Finland | FinnGen Analysis working group | FinnGen Analysis working group |
| **Wei Zhou** | Broad Institute, Cambridge, MA, United States | FinnGen Analysis working group | FinnGen Analysis working group |
| **Mutaamba Maasha** | Broad Institute, Cambridge, MA, United States | FinnGen Analysis working group | FinnGen Analysis working group |
| **Shabbeer Hassan** | Institute for Molecular Medicine Finland (FIMM), HiLIFE, University of Helsinki, Helsinki, Finland | FinnGen Analysis working group | FinnGen Analysis working group |
| **Susanna Lemmelä** | Institute for Molecular Medicine Finland (FIMM), HiLIFE, University of Helsinki, Helsinki, Finland | FinnGen Analysis working group | FinnGen Analysis working group |

| | | | |
|---|---|---|---|
| **Manuel Rivas** | University of Stanford, Stanford, CA, United States | **FinnGen Analysis working group** | **FinnGen Analysis working group** |
| **Aarno Palotie** | Institute for Molecular Medicine Finland (FIMM), HiLIFE, University of Helsinki, Helsinki, Finland | **FinnGen Analysis working group** | **FinnGen Analysis working group** |
| **Aoxing Liu** | Institute for Molecular Medicine Finland (FIMM), HiLIFE, University of Helsinki, Helsinki, Finland | **FinnGen Analysis working group** | **FinnGen Analysis working group** |
| **Arto Lehisto** | Institute for Molecular Medicine Finland (FIMM), HiLIFE, University of Helsinki, Helsinki, Finland | **FinnGen Analysis working group** | **FinnGen Analysis working group** |
| **Andrea Ganna** | Institute for Molecular Medicine Finland (FIMM), HiLIFE, University of Helsinki, Helsinki, Finland | **FinnGen Analysis working group** | **FinnGen Analysis working group** |
| **Vincent Llorens** | Institute for Molecular Medicine Finland (FIMM), HiLIFE, University of Helsinki, Helsinki, Finland | **FinnGen Analysis working group** | **FinnGen Analysis working group** |
| **Hannele Laivuori** | Institute for Molecular Medicine Finland (FIMM), HiLIFE, University of Helsinki, Helsinki, Finland | **FinnGen Analysis working group** | **FinnGen Analysis working group** |
| **Taru Tukiainen** | Institute for Molecular Medicine Finland (FIMM), HiLIFE, University of Helsinki, Helsinki, Finland | **FinnGen Analysis working group** | **FinnGen Analysis working group** |
| **Mary Pat Reeve** | Institute for Molecular Medicine Finland (FIMM), HiLIFE, University of Helsinki, Helsinki, Finland | **FinnGen Analysis working group** | **FinnGen Analysis working group** |
| **Henrike Heyne** | Institute for Molecular Medicine Finland (FIMM), HiLIFE, University of Helsinki, Helsinki, Finland | **FinnGen Analysis working group** | **FinnGen Analysis working group** |
| **Nina Mars** | Institute for Molecular Medicine Finland (FIMM), HiLIFE, University of Helsinki, Helsinki, Finland | **FinnGen Analysis working group** | **FinnGen Analysis working group** |
| **Joel Rämö** | Institute for Molecular Medicine Finland (FIMM), HiLIFE, University of Helsinki, Helsinki, Finland | **FinnGen Analysis working group** | **FinnGen Analysis working group** |
| **Elmo Saarentaus** | Institute for Molecular Medicine Finland (FIMM), HiLIFE, University of Helsinki, Helsinki, Finland | **FinnGen Analysis working group** | **FinnGen Analysis working group** |
| **Hanna Ollila** | Institute for Molecular Medicine Finland (FIMM), HiLIFE, University of Helsinki, Helsinki, Finland | **FinnGen Analysis working group** | **FinnGen Analysis working group** |
| **Rodos Rodosthenous** | Institute for Molecular Medicine Finland (FIMM), HiLIFE, University of Helsinki, Helsinki, Finland | **FinnGen Analysis working group** | **FinnGen Analysis working group** |
| **Satu Strausz** | Institute for Molecular Medicine Finland (FIMM), HiLIFE, University of Helsinki, Helsinki, Finland | **FinnGen Analysis working group** | **FinnGen Analysis working group** |
| **Tuula Palotie** | University of Helsinki and Hospital District of Helsinki and Uusimaa, Helsinki, Finland | **FinnGen Analysis working group** | **FinnGen Analysis working group** |
| **Kimmo Palin** | University of Helsinki, Helsinki, Finland | **FinnGen Analysis working group** | **FinnGen Analysis working group** |

| | | | |
|---|---|---|---|
| **Javier Garcia-Tabuenca** | University of Tampere, Tampere, Finland | **FinnGen Analysis working group** | **FinnGen Analysis working group** |
| **Harri Siirtola** | University of Tampere, Tampere, Finland | **FinnGen Analysis working group** | **FinnGen Analysis working group** |
| **Tuomo Kiiskinen** | Institute for Molecular Medicine Finland (FIMM), HiLIFE, University of Helsinki, Helsinki, Finland | **FinnGen Analysis working group** | **FinnGen Analysis working group** |
| **Jiwoo Lee** | Institute for Molecular Medicine Finland (FIMM), HiLIFE, University of Helsinki, Helsinki, Finland; Broad Institute, Cambridge, MA, United States | **FinnGen Analysis working group** | **FinnGen Analysis working group** |
| **Kristin Tsuo** | Institute for Molecular Medicine Finland (FIMM), HiLIFE, University of Helsinki, Helsinki, Finland; Broad Institute, Cambridge, MA, United States | **FinnGen Analysis working group** | **FinnGen Analysis working group** |
| **Amanda Elliott** | Institute for Molecular Medicine Finland (FIMM), HiLIFE, University of Helsinki, Helsinki, Finland; Broad Institute, Cambridge, MA, USA and Massachusetts General Hospital, Boston, MA, USA | **FinnGen Analysis working group** | **FinnGen Analysis working group** |
| **Kati Kristiansson** | THL Biobank / Finnish Institute for Health and Welfare (THL), Helsinki, Finland | **FinnGen Analysis working group** | **FinnGen Analysis working group** |
| **Mikko Arvas** | Finnish Red Cross Blood Service / Finnish Hematology Registry and Clinical Biobank, Helsinki, Finland | **FinnGen Analysis working group** | **FinnGen Analysis working group** |
| **Kati Hyvärinen** | Finnish Red Cross Blood Service, Helsinki, Finland | **FinnGen Analysis working group** | **FinnGen Analysis working group** |
| **Jarmo Ritari** | Finnish Red Cross Blood Service, Helsinki, Finland | **FinnGen Analysis working group** | **FinnGen Analysis working group** |
| **Olli Carpén** | Helsinki Biobank / Helsinki University and Hospital District of Helsinki and Uusimaa, Helsinki | **FinnGen Analysis working group** | **FinnGen Analysis working group** |
| **Johannes Kettunen** | Northern Finland Biobank Borealis / University of Oulu / Northern Ostrobothnia Hospital District, Oulu, Finland | **FinnGen Analysis working group** | **FinnGen Analysis working group** |
| **Katri Pylkäs** | University of Oulu, Oulu, Finland | **FinnGen Analysis working group** | **FinnGen Analysis working group** |
| **Eeva Sliz** | University of Oulu, Oulu, Finland | **FinnGen Analysis working group** | **FinnGen Analysis working group** |
| **Minna Karjalainen** | University of Oulu, Oulu, Finland | **FinnGen Analysis working group** | **FinnGen Analysis working group** |
| **Tuomo Mantere** | Northern Finland Biobank Borealis / University of Oulu / Northern Ostrobothnia Hospital District, Oulu, Finland | **FinnGen Analysis working group** | **FinnGen Analysis working group** |
| **Eeva Kangasniemi** | Finnish Clinical Biobank Tampere / University of Tampere / Pirkanmaa Hospital District, Tampere, Finland | **FinnGen Analysis working group** | **FinnGen Analysis working group** |
| **Sami Heikkinen** | University of Eastern Finland, Kuopio, Finland | **FinnGen Analysis working group** | **FinnGen Analysis working group** |

| | | | |
|---|---|---|---|
| **Arto Mannermaa** | Biobank of Eastern Finland / University of Eastern Finland / Northern Savo Hospital District, Kuopio, Finland | **FinnGen Analysis working group** | **FinnGen Analysis working group** |
| **Eija Laakkonen** | University of Jyväskylä, Jyväskylä, Finland | **FinnGen Analysis working group** | **FinnGen Analysis working group** |
| **Nina Pitkänen** | Auria Biobank / University of Turku / Hospital District of Southwest Finland, Turku, Finland | **FinnGen Analysis working group** | **FinnGen Analysis working group** |
| **Samuel Lessard** | Translational Sciences, Sanofi R&D, Framingham, MA, USA | **FinnGen Analysis working group** | **FinnGen Analysis working group** |
| **Clément Chatelain** | Translational Sciences, Sanofi R&D, Framingham, MA, USA | **FinnGen Analysis working group** | **FinnGen Analysis working group** |
| **Lila Kallio** | Auria Biobank / University of Turku / Hospital District of Southwest Finland, Turku, Finland | **Biobank directors** | **Biobank directors** |
| **Tiina Wahlfors** | THL Biobank / Finnish Institute for Health and Welfare (THL), Helsinki, Finland | **Biobank directors** | **Biobank directors** |
| **Jukka Partanen** | Finnish Red Cross Blood Service / Finnish Hematology Registry and Clinical Biobank, Helsinki, Finland | **Biobank directors** | **Biobank directors** |
| **Eero Punkka** | Helsinki Biobank / Helsinki University and Hospital District of Helsinki and Uusimaa, Helsinki | **Biobank directors** | **Biobank directors** |
| **Raisa Serpi** | Northern Finland Biobank Borealis / University of Oulu / Northern Ostrobothnia Hospital District, Oulu, Finland | **Biobank directors** | **Biobank directors** |
| **Sanna Siltanen** | Finnish Clinical Biobank Tampere / University of Tampere / Pirkanmaa Hospital District, Tampere, Finland | **Biobank directors** | **Biobank directors** |
| **Veli-Matti Kosma** | Biobank of Eastern Finland / University of Eastern Finland / Northern Savo Hospital District, Kuopio, Finland | **Biobank directors** | **Biobank directors** |
| **Teijo Kuopio** | Central Finland Biobank / University of Jyväskylä / Central Finland Health Care District, Jyväskylä, Finland | **Biobank directors** | **Biobank directors** |
| **Anu Jalanko** | Institute for Molecular Medicine Finland (FIMM), HiLIFE, University of Helsinki, Helsinki, Finland | **FinnGen Teams** | **Administration** |
| **Huei-Yi Shen** | Institute for Molecular Medicine Finland (FIMM), HiLIFE, University of Helsinki, Helsinki, Finland | **FinnGen Teams** | **Administration** |
| **Risto Kajanne** | Institute for Molecular Medicine Finland (FIMM), HiLIFE, University of Helsinki, Helsinki, Finland | **FinnGen Teams** | **Administration** |
| **Mervi Aavikko** | Institute for Molecular Medicine Finland (FIMM), HiLIFE, University of Helsinki, Helsinki, Finland | **FinnGen Teams** | **Administration** |
| **Helen Cooper** | Institute for Molecular Medicine Finland (FIMM), HiLIFE, University of Helsinki, Helsinki, Finland | **FinnGen Teams** | **Administration** |
| **Denise Öller** | Institute for Molecular Medicine Finland (FIMM), HiLIFE, University of Helsinki, Helsinki, Finland | **FinnGen Teams** | **Administration** |
| **Rasko Leinonen** | Institute for Molecular Medicine Finland (FIMM), HiLIFE, University of Helsinki, Helsinki, Finland; European Molecular Biology Laboratory, European Bioinformatics Institute, Cambridge, UK | **FinnGen Teams** | **Administration** |
| **Henna Palin** | Finnish Clinical Biobank Tampere / University of Tampere / Pirkanmaa Hospital District, Tampere, Finland | **FinnGen Teams** | **Administration** |
| **Malla-Maria Linna** | Helsinki Biobank / Helsinki University and Hospital District of Helsinki and Uusimaa, Helsinki | **FinnGen Teams** | **Administration** |
| **Mitja Kurki** | Institute for Molecular Medicine Finland (FIMM), HiLIFE, University of Helsinki, Helsinki, Finland; Broad Institute, Cambridge, MA, United States | **FinnGen Teams** | **Analysis** |
| **Juha Karjalainen** | Institute for Molecular Medicine Finland (FIMM), HiLIFE, University of Helsinki, Helsinki, Finland | **FinnGen Teams** | **Analysis** |
| **Pietro Della Briotta Parolo** | Institute for Molecular Medicine Finland (FIMM), HiLIFE, University of Helsinki, Helsinki, Finland | **FinnGen Teams** | **Analysis** |
| **Arto Lehisto** | Institute for Molecular Medicine Finland (FIMM), HiLIFE, University of Helsinki, Helsinki, Finland | **FinnGen Teams** | **Analysis** |
| **Juha Mehtonen** | Institute for Molecular Medicine Finland (FIMM), HiLIFE, University of Helsinki, Helsinki, Finland | **FinnGen Teams** | **Analysis** |
| **Wei Zhou** | Broad Institute, Cambridge, MA, United States | **FinnGen Teams** | **Analysis** |
| **Masahiro Kanai** | Broad Institute, Cambridge, MA, United States | **FinnGen Teams** | **Analysis** |
| **Mutaamba Maasha** | Broad Institute, Cambridge, MA, United States | **FinnGen Teams** | **Analysis** |

| | | | |
|---|---|---|---|
| **Zhili Zheng** | Broad Institute, Cambridge, MA, United States | **FinnGen Teams** | **Analysis** |
| **Hannele Laivuori** | Institute for Molecular Medicine Finland (FIMM), HiLIFE, University of Helsinki, Helsinki, Finland | **FinnGen Teams** | **Clinical Endpoint Development** |
| **Aki Havulinna** | Institute for Molecular Medicine Finland (FIMM), HiLIFE, University of Helsinki, Helsinki, Finland; Finnish Institute for Health and Welfare (THL), Helsinki, Finland | **FinnGen Teams** | **Clinical Endpoint Development** |
| **Susanna Lemmelä** | Institute for Molecular Medicine Finland (FIMM), HiLIFE, University of Helsinki, Helsinki, Finland | **FinnGen Teams** | **Clinical Endpoint Development** |
| **Tuomo Kiiskinen** | Institute for Molecular Medicine Finland (FIMM), HiLIFE, University of Helsinki, Helsinki, Finland | **FinnGen Teams** | **Clinical Endpoint Development** |
| **L. Elisa Lahtela** | Institute for Molecular Medicine Finland (FIMM), HiLIFE, University of Helsinki, Helsinki, Finland | **FinnGen Teams** | **Clinical Endpoint Development** |
| **Mari Kaunisto** | Institute for Molecular Medicine Finland (FIMM), HiLIFE, University of Helsinki, Helsinki, Finland | **FinnGen Teams** | **Communication** |
| **Elina Kilpeläinen** | Institute for Molecular Medicine Finland (FIMM), HiLIFE, University of Helsinki, Helsinki, Finland | **FinnGen Teams** | **E-Science** |
| **Timo P. Sipilä** | Institute for Molecular Medicine Finland (FIMM), HiLIFE, University of Helsinki, Helsinki, Finland | **FinnGen Teams** | **E-Science** |
| **Oluwaseun Alexander Dada** | Institute for Molecular Medicine Finland (FIMM), HiLIFE, University of Helsinki, Helsinki, Finland | **FinnGen Teams** | **E-Science** |
| **Awaisa Ghazal** | Institute for Molecular Medicine Finland (FIMM), HiLIFE, University of Helsinki, Helsinki, Finland | **FinnGen Teams** | **E-Science** |
| **Anastasia Kytölä** | Institute for Molecular Medicine Finland (FIMM), HiLIFE, University of Helsinki, Helsinki, Finland | **FinnGen Teams** | **E-Science** |
| **Rigbe Weldatsadik** | Institute for Molecular Medicine Finland (FIMM), HiLIFE, University of Helsinki, Helsinki, Finland | **FinnGen Teams** | **E-Science** |
| **Sanni Ruotsalainen** | Institute for Molecular Medicine Finland (FIMM), HiLIFE, University of Helsinki, Helsinki, Finland | **FinnGen Teams** | **E-Science** |
| **Kati Donner** | Institute for Molecular Medicine Finland (FIMM), HiLIFE, University of Helsinki, Helsinki, Finland | **FinnGen Teams** | **Genotyping** |
| **Timo P. Sipilä** | Institute for Molecular Medicine Finland (FIMM), HiLIFE, University of Helsinki, Helsinki, Finland | **FinnGen Teams** | **Genotyping** |
| **Anu Loukola** | Helsinki Biobank / Helsinki University and Hospital District of Helsinki and Uusimaa, Helsinki | **FinnGen Teams** | **Sample Collection Coordination** |
| **Päivi Laiho** | THL Biobank / Finnish Institute for Health and Welfare (THL), Helsinki, Finland | **FinnGen Teams** | **Sample Logistics** |
| **Tuuli Sistonen** | THL Biobank / Finnish Institute for Health and Welfare (THL), Helsinki, Finland | **FinnGen Teams** | **Sample Logistics** |
| **Essi Kaiharju** | THL Biobank / Finnish Institute for Health and Welfare (THL), Helsinki, Finland | **FinnGen Teams** | **Sample Logistics** |
| **Markku Laukkanen** | THL Biobank / Finnish Institute for Health and Welfare (THL), Helsinki, Finland | **FinnGen Teams** | **Sample Logistics** |
| **Elina Järvensivu** | THL Biobank / Finnish Institute for Health and Welfare (THL), Helsinki, Finland | **FinnGen Teams** | **Sample Logistics** |
| **Sini Lähteenmäki** | THL Biobank / Finnish Institute for Health and Welfare (THL), Helsinki, Finland | **FinnGen Teams** | **Sample Logistics** |
| **Lotta Männikkö** | THL Biobank / Finnish Institute for Health and Welfare (THL), Helsinki, Finland | **FinnGen Teams** | **Sample Logistics** |
| **Regis Wong** | THL Biobank / Finnish Institute for Health and Welfare (THL), Helsinki, Finland | **FinnGen Teams** | **Sample Logistics** |
| **Auli Toivola** | THL Biobank / Finnish Institute for Health and Welfare (THL), Helsinki, Finland | **FinnGen Teams** | **Sample Logistics** |
| **Minna Brunfeldt** | THL Biobank / Finnish Institute for Health and Welfare (THL), Helsinki, Finland | **FinnGen Teams** | **Registry Data Operations** |
| **Hannele Mattsson** | THL Biobank / Finnish Institute for Health and Welfare (THL), Helsinki, Finland | **FinnGen Teams** | **Registry Data Operations** |
| **Kati Kristiansson** | THL Biobank / Finnish Institute for Health and Welfare (THL), Helsinki, Finland | **FinnGen Teams** | **Registry Data Operations** |
| **Susanna Lemmelä** | Institute for Molecular Medicine Finland (FIMM), HiLIFE, University of Helsinki, Helsinki, Finland | **FinnGen Teams** | **Registry Data Operations** |
| **Sami Koskelainen** | THL Biobank / Finnish Institute for Health and Welfare (THL), Helsinki, Finland | **FinnGen Teams** | **Registry Data Operations** |
| **Tero Hiekkalinna** | THL Biobank / Finnish Institute for Health and Welfare (THL), Helsinki, Finland | **FinnGen Teams** | **Registry Data Operations** |
| **Teemu Paajanen** | THL Biobank / Finnish Institute for Health and Welfare (THL), Helsinki, Finland | **FinnGen Teams** | **Registry Data Operations** |
| **Priit Palta** | Institute for Molecular Medicine Finland (FIMM), HiLIFE, University of Helsinki, Helsinki, Finland | **FinnGen Teams** | **Sequencing Informatics** |
| **Kalle Pärn** | Institute for Molecular Medicine Finland (FIMM), HiLIFE, University of Helsinki, Helsinki, Finland | **FinnGen Teams** | **Sequencing Informatics** |

| | | | |
|---|---|---|---|
| **Mart Kals** | Institute for Molecular Medicine Finland (FIMM), HiLIFE, University of Helsinki, Helsinki, Finland | FinnGen Teams | **Sequencing Informatics** |
| **Shuang Luo** | Institute for Molecular Medicine Finland (FIMM), HiLIFE, University of Helsinki, Helsinki, Finland | FinnGen Teams | **Sequencing Informatics** |
| **Tarja Laitinen** | Pirkanmaa Hospital District, Tampere, Finland | FinnGen Teams | **Trajectory** |
| **Mary Pat Reeve** | Institute for Molecular Medicine Finland (FIMM), HiLIFE, University of Helsinki, Helsinki, Finland | FinnGen Teams | **Trajectory** |
| **Shanmukha Sampath Padmanabhuni** | Institute for Molecular Medicine Finland (FIMM), HiLIFE, University of Helsinki, Helsinki, Finland | FinnGen Teams | **Trajectory** |
| **Marianna Niemi** | University of Tampere, Tampere, Finland | FinnGen Teams | **Trajectory** |
| **Harri Siirtola** | University of Tampere, Tampere, Finland | FinnGen Teams | **Trajectory** |
| **Javier Gracia-Tabuenca** | University of Tampere, Tampere, Finland | FinnGen Teams | **Trajectory** |
| **Mika Helminen** | University of Tampere, Tampere, Finland | FinnGen Teams | **Trajectory** |
| **Tiina Luukkaala** | University of Tampere, Tampere, Finland | FinnGen Teams | **Trajectory** |
| **Iida Vähätalo** | University of Tampere, Tampere, Finland | FinnGen Teams | **Trajectory** |
| **Jyrki Tammerluoto** | Institute for Molecular Medicine Finland (FIMM), HiLIFE, University of Helsinki, Helsinki, Finland | FinnGen Teams | **Data protection officer** |
| **Marco Hautalahti** | Finnish Biobank Cooperative - FINBB | FinnGen Teams | **FINBB - Finnish biobank cooperative** |
| **Johanna Mäkelä** | Finnish Biobank Cooperative - FINBB | FinnGen Teams | **FINBB - Finnish biobank cooperative** |
| **Sarah Smith** | Finnish Biobank Cooperative - FINBB | FinnGen Teams | **FINBB - Finnish biobank cooperative** |
| **Tom Southerington** | Finnish Biobank Cooperative - FINBB | FinnGen Teams | **FINBB - Finnish biobank cooperative** |
| **Petri Lehto** | Finnish Biobank Cooperative - FINBB | FinnGen Teams | **FINBB - Finnish biobank cooperative** |

**Table S12** A list of Estonian Biobank Research Team authors and their affiliations

| Full Name | Affiliation |
|---|---|
| **Andres Metspalu** | Estonian Genome Centre, Institute of Genomics, University of Tartu, Tartu, Estonia |
| **Mari Nelis** | Estonian Genome Centre, Institute of Genomics, University of Tartu, Tartu, Estonia |
| **Lili Milani** | Estonian Genome Centre, Institute of Genomics, University of Tartu, Tartu, Estonia |
| **Reedik Mägi** | Estonian Genome Centre, Institute of Genomics, University of Tartu, Tartu, Estonia |
| **Georgi Hudjashov** | Estonian Genome Centre, Institute of Genomics, University of Tartu, Tartu, Estonia |
| **Tõnu Esko** | Estonian Genome Centre, Institute of Genomics, University of Tartu, Tartu, Estonia |
